# Estimating the impact of implementation and timing of the COVID-19 vaccination programme in Brazil: a counterfactual analysis

**DOI:** 10.1101/2021.12.24.21268384

**Authors:** Leonardo Souto Ferreira, Flavia Maria Darcie Marquitti, Rafael Lopes Paixão da Silva, Marcelo Eduardo Borges, Marcelo Ferreira da Costa Gomes, Oswaldo Gonçalves Cruz, Roberto André Kraenkel, Renato Mendes Coutinho, Paulo Inácio Prado, Leonardo Soares Bastos

## Abstract

**Background:** Vaccines developed between 2020 - 2021 against the SARS-CoV-2 virus were designed to diminish the severity and prevent deaths due to COVID-19. However, estimates of the effectiveness of vaccination campaigns in achieving these goals remain a methodological challenge. In this work, we developed a Bayesian statistical model to estimate the number of deaths and hospitalisations averted by vaccination of older adults (above 60 years old) in Brazil.

**Methods:** We fit a linear model to predict the number of deaths and hospitalisations of older adults as a function of vaccination coverage in this group and casualties in younger adults. We used this model in a counterfactual analysis, simulating alternative scenarios without vaccination or with faster vaccination roll-out. We estimated the direct effects of COVID-19 vaccination by computing the difference between hypothetical and realised scenarios.

**Findings:** We estimated that more than 165,000 individuals above 60 years of age were not hospitalised due to COVID-19 in the first seven months of the vaccination campaign. An additional contingent of 104,000 hospitalisations could have been averted if vaccination had started earlier. We also estimated that more than 58 thousand lives were saved by vaccinations in the period analysed for the same age group and that an additional 47 thousand lives could have been saved had the Brazilian government started the vaccination programme earlier.

**Interpretation:** Our estimates provided a lower bound for vaccination impacts in Brazil, demonstrating the importance of preventing the suffering and loss of older Brazilian adults. Once vaccines were approved, an early vaccination roll-out could have saved many more lives, especially when facing a pandemic.

**Funding:** The Coordenação de Aperfeiçoamento de Pessoal de Nível Superior – Brazil (Finance Code 001 to FMDM and LSF), Conselho Nacional de Desenvolvimento Científico e Tecnológico – Brazil (grant number: 315854/2020-0 to MEB, 141698/2018-7 to RLPS, 313055/2020-3 to PIP, 311832/2017-2 to RAK), Fundação de Amparo à Pesquisa do Estado de São Paulo – Brazil (contract number: 2016/01343-7 to RAK), Fundação de Amparo à Pesquisa do Estado do Rio de Janeiro – Brazil (grant number: E-26/201.277/2021 to LSB) and Inova Fiocruz/Fundação Oswaldo Cruz – Brazil (grant number: 48401485034116) to LSB, OGC and MGFC. The funding agencies had no role in the conceptualization of the study.

## INTRODUCTION

Since March 15, 2020, SARS-CoV-2 has been declared in community transmission in Brazil. During the first year of the pandemic, the epidemic spread fast in Brazil but with different timings and burdens between regions due to regional differences in health assistance, income and local mitigation policies.^1,2^ On top of that, by January 2021, Brazil’s epidemic saw a strong increase in the number of notified cases and deaths due to SARS-CoV-2, especially in the northern region of Brazil.^2^ The new burst quickly spread to the rest of the country, synchronizing as waves in each region, ultimately reducing by the end of May. These waves were later associated with the appearance of the variant of concern (VOC) P.1, also known as Gamma, whose emergence was estimated as of November 2020 to be in Manaus.^3,4^ Brazil also had community transmission of Alpha VOC. However, Alpha VOC did not overcome Gamma, because the latter was found to be more transmissible and with a potential immunity escape.^5,6,4^ The Gamma variant was eventually replaced by the Delta variant in relative frequency, although the majority of Brazilian COVID-19 hospitalisations (≈82%) and deaths (≈81.5%) in 2021 occurred during the Gamma dominance.^7^ The country did not suffer another marked increase in cases and deaths during the rest of 2021 as other countries, and this difference is attributed to the vaccination campaign in Brazil.

Brazil has an outstanding history of successful government policies for mass vaccination, including coordinated vaccination campaigns at the country level, effective communication strategies, free availability of doses, and the capillarity of the Brazilian Unified Health System (SUS). For example, in 2010, SUS was able to quickly vaccinate 89 million individuals in response to the 2009 H1N1 influenza pandemic.^8,9^ However, due to several funding cuts and widespread misinformation, the subsequent vaccination campaigns could not surpass the coverage objectives.^10,11,12^ The COVID-19 vaccination campaign in Brazil suffered from poor coordination and logistics at the federal level,^13^ which delayed and slowed down the pace of vaccine roll-out. Vaccination eventually started on January 17, 2021, first covering institutionalised people, Indigenous peoples, and health professionals. After that, the vaccination roll-out was structured considering age groups, from older to younger individuals, on an at-risk basis.^14,15^ By December 22, 2021, Brazil had 88.9% and 66.7% of the total population vaccinated with one and two doses, respectively,^16^ with an ongoing campaign of booster inoculation. This coverage surpasses richer countries that had earlier availability of vaccines. The success may be attributed to some foundations Brazil had already built before the pandemic started: a national system of health (SUS) which provides care even in the most remote areas of the large country, and with a long tradition of mass vaccination campaigns; state-level government policies which could be more (and never less) rigorous than the federal policies during the pandemic; and strong beliefs within the population regarding vaccination reinforced through years of good results of immunisation campaigns, despite recent hesitancy movements and the high levels of misinformation.

However, information about the effectiveness of the current vaccination campaign in preventing hospitalisations and deaths countrywide, the main purpose of the developed vaccines, still lacks a proper estimation. The only estimates available are for the state of São Paulo, the most populous state with the highest GDP in Brazil.^17^ Around 24 thousand hospitalisations and 11 thousand deaths have been averted by vaccinations in São Paulo in the age group of >65 years between February 8 and May 28 of 2021, reducing hospitalisation costs by US$287 million.^17^ Thus, our objective was to expand these analyses to the whole country, also accounting for other possible scenarios of vaccination roll-outs. Previous research shows the deaths of individuals above 80 y.o. were constant at about 25 - 30% of all reported COVID-19 deaths at any age, decreasing to 13% in May 2021, after vaccination of this age group; the same trend was observed for the 70 - 79 y.o. age group after a time lag.^18^

We developed a statistical model to predict the number of deaths and hospitalisations by COVID-19 in the age group of older adults, separate from the time series of deaths and cases in younger age groups. The model considered the reduction in relative risk to older age groups as vaccine coverage progressed in this population over time. We then used the estimated effect of vaccine coverage on reducing relative risk in a counterfactual analysis to estimate the direct effect of vaccination in averting hospitalisations and deaths by COVID-19 in Brazil. Since the model directly accounted for vaccination, we also provided estimates for the potential number of hospitalisations and deaths averted if vaccines were made available earlier to the population. The analysis was conducted considering the age group of adults above 60 years old with a time series that runs until August 28, 2021.

## METHODS

### Data

The weekly count of hospitalisations and deaths due to COVID-19 indicated as a Severe Acute Respiratory Infection (SARI), in each age group, was obtained from the Influenza Epidemiological Surveillance Information System (SIVEP-Gripe),^19,20^ extracted on October 18, 2021. This database is publicly available by the Brazilian National Health System. No ethical approval is needed according to the National Ethical Commission (CONEP) of the National Health Council, Resolution Number 510 of April 7, 2016. Shortly after the pandemic onset in Brazil, the anonymised data was published with all the individual cases notified by healthcare units, and municipal and state health secretariats. We filtered the dataset to keep only the cases that were hospitalised or died due to SARI associated with COVID-19. The association with COVID-19 was filtered based on the case classification field (CLASSI_FIN), plus all cases with a positive RT-PCR test for SARS-CoV-2, regardless of classification. All cases were aggregated by the state of residence. For the weekly count of hospitalised cases, the date of reference is based on symptom onset. We also used the date of symptom onset as a reference for the weekly number of deaths. The vaccination data was extracted from the National Immunisation Plan Information System (SIPNI),^16^ on November 6, 2021, which included the location, date, type of vaccine, dose (1st, 2nd, 3rd) of vaccine, and age of every person vaccinated against COVID-19 in Brazil. For the SIPNI, we considered the state where the vaccine was applied instead of the state of residence of the individual, as the latter is not always available; although people can be vaccinated in cities other than the one they reside, the difference in numbers is negligible at the state level. Cases (hospitalisations and deaths) and vaccination status were aggregated by age group (20 - 29 y.o: reference group; 60 - 69, 70 - 79 and >80 y.o: older adults). For the results, we presented data for each older adult class (60 - 69, 70 - 79 and >80), aggregated older adults (>60), and by country level. In the ***Supplementary Material*** (hereafter ***SM***) we presented the Brazilian time series of hospitalisation (see **Fig. S1** in the ***SM***), deaths (**Fig. S2** in the ***SM***) and the different variants (**Fig. S3** in the ***SM***) since the beginning of the pandemic.

### Statistical model

We built a Bayesian statistical model to infer and predict the number of hospitalisations and deaths due to COVID-19 in age classes above 60 y.o. (>60) as a linear function of both the number of hospitalisations and deaths in the 20 - 29 age class and the vaccine coverage in each age class. We used the coverage of the second dose (counting from 14 days post-inoculation) of each vaccine *v* = {*AZD*1222, *CoronaVac, BNT*162*b*2}, produced by AstraZeneca/Oxford/Fiocruz, Sinovac/Butantan and Pfizer/BioNTech, respectively, from the SIPNI.^16^ Aside from these three vaccines, Ad26.COV2.S (Janssen) was also used in Brazilian territory, however, we considered that the number of inoculated doses was too low to have statistical significance in this analysis. The model is given by:

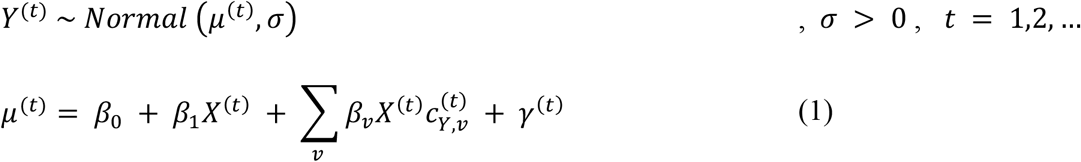

where *Y*^(*t*)^ denotes the number of hospitalisations or deaths at time *t* expected in the age group being studied, *X*^(*t*)^ is the number of hospitalisations or deaths at time *t* of the age group being used as reference, 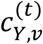 denotes the coverage of vaccine *v* in the studied age group at time *t*, and *γ*^(*t*)^ for *t* = 1,2, … represents temporal Gaussian random effects, modelled as a first-order autoregressive process of order 1, AR(1), as the following

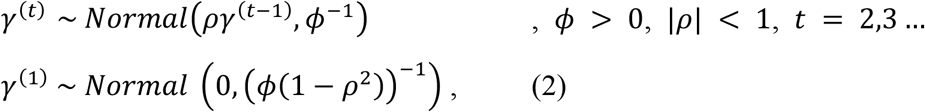

where *ρ* is a temporal correlation and *ϕ* is the random effects precision.

The model assumes *Y* is proportional to *X* in the absence of vaccination. The third term in Equation 1 expresses that, when vaccination is present, the difference between *X* and *Y* is linearly related to the coverage of each vaccine (note that a different β is fitted to each vaccine). Finally, the latter term of Eq. 1 accounts for temporal dependence among hospitalisations or deaths. We also provided a comparison between different random effect models and a model without random effects in the ***SM***. By using the Watanabe-Akaike Information Criteria (WAIC), the autoregressive process of the order 1 model had the highest relative likelihood in the majority of cases, thus was our choice for the random effect model (see **Fig. S4** in the ***SM***)

Finally, our prior distributions were given (in terms of precision) by:

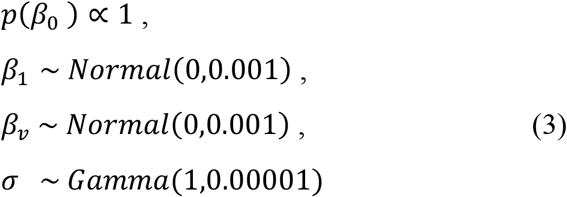

and the prior distributions of the AR(1) random effects (following the notation of Rue et al.^21^) were given by:

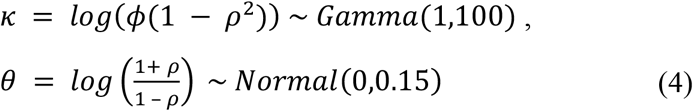

Cases related to hospitalisations and deaths, for each age class and for each state, were modelled independently and used the same priors, without sharing parameters.

The inference was made using the integrated nested Laplace approximation (INLA) approach,^21,22^ implemented in R.^23^ The models were fitted independently for each age group of older adults (categorised here as 60 - 69, 70 - 79 and >80) and Brazilian state using the 20 - 29 age group as reference one (*X*) between March 1, 2020, and August 29, 2021 – for sensitivity analysis regarding the reference groups, check **Tables S2** and **S3** in the ***SM***. The posterior trajectories of fitted and simulated time series were drawn from 1,000 samples for each simulation set (age group and state) and added to generate an aggregated posterior sample for the whole country. Therefore, the distribution of these sums of trajectories provided the 95% Credible Interval (hereafter, 95% CI) for the national trajectories for each age group.

To estimate the effect of vaccination, we set the coverage values to zero and predicted the number of hospitalisations and deaths expected in the absence of the explanatory variables of coverage. Then, we compared the cumulative number of hospitalisations and deaths of this hypothetical trajectory with their equivalent when observed vaccine coverage is considered from January 1, 2021 up to August 29, 2021. We also estimated the effect of vaccination on COVID-19 dynamics if vaccines had been made available earlier in 2021. To achieve this, we created two counterfactual scenarios of faster vaccination by keeping the same pace and shifting the time series of observed vaccine coverage to start moderately or highly accelerated. These time series were moved to four and eight weeks earlier, and vaccination cases recorded earlier than January 17, 2020 (date of first vaccination) were all set to this date. The counterfactual scenarios were repeated for each target age group, considering the same amount of vaccines were available in these faster starting scenarios. To keep the time series until August 29, 2021, in these hypothetical scenarios, we used the four and eight following weeks in the observed time series.

## RESULTS

In **Figure 1**, the observed time series (dark orange curve and dots) and the estimated series without vaccination (light orange curve) presented similar trajectories until May for 60 - 79 y.o, and until March - April for the age group of more than 80 y.o (>80). The curves are also presented for each state in the Results Section of the ***Supplementary Material***. We also presented the similarity between the fitted model and the observed data in **Fig. S5** in the ***SM***. The overlap of these curves showed that the risk of hospitalisations and deaths in the target groups relative to the reference group was little affected during the period when vaccine coverage was still low. Therefore, the model provided a realistic estimate of the reduction of the relative risk of casualties in the target groups as coverage increases (see **Fig. S6** and **S7** in the ***SM***).

**Figure 1:**
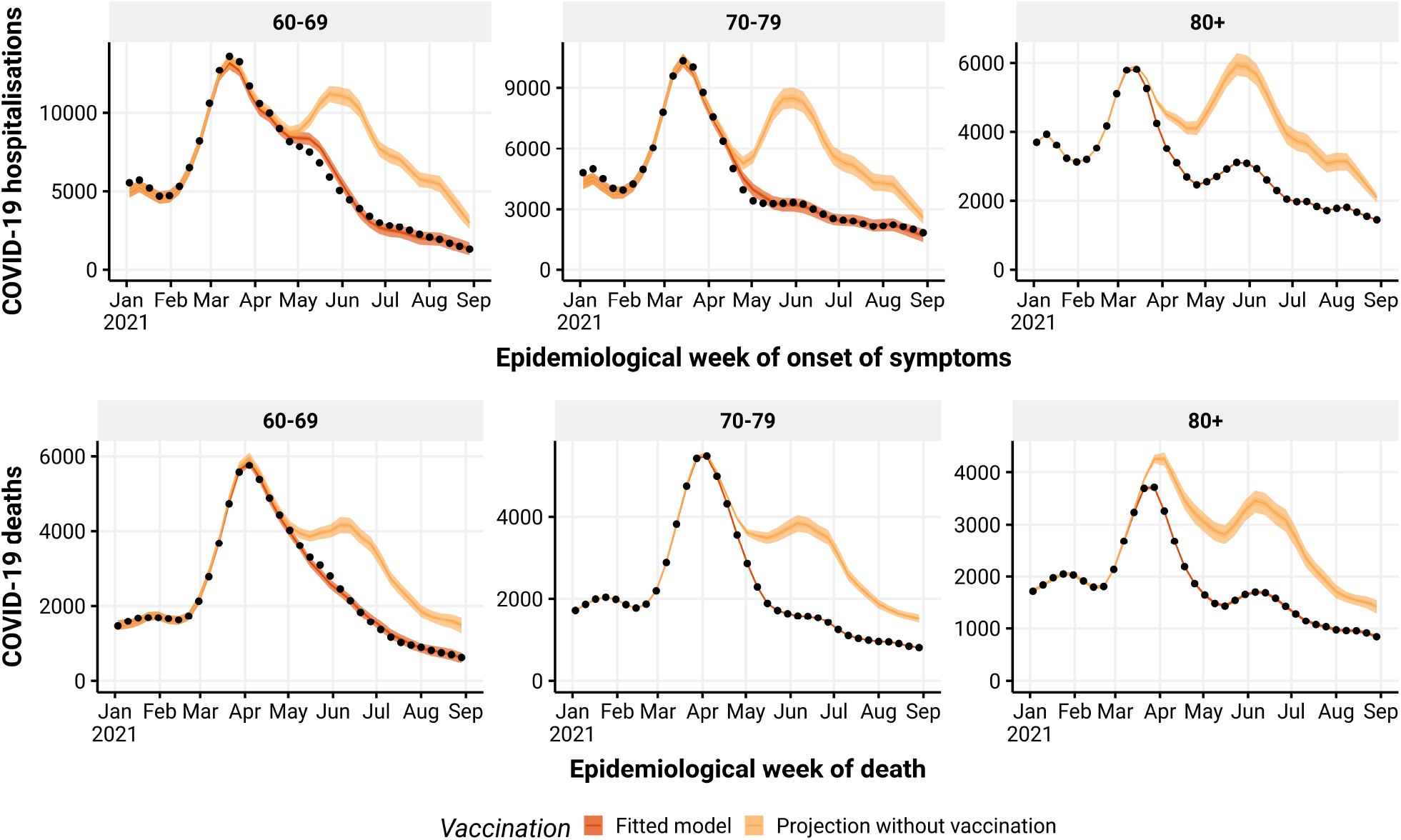
Estimated number of hospitalisations (top) and deaths (bottom) by epidemiological week with (dark orange) or without (light orange) vaccination roll-out, by age group (panels). The observed number of hospitalisations and deaths are given by the black dots. Line: median, shadow: 95% CI.

Vaccination was not able to suppress the wave of hospitalisation cases due to the Gamma variant, which occurred from late January 2021 to late March 2021 (**Fig. 1**). Although for 60 - 79 y.o., the estimates of deaths showed no difference when compared to the observed deaths during the Gamma wave, vaccination decreased the overall number of fatal cases for those who were vaccinated earlier (>80) and who likely had a more robust immunity when the Gamma VOC hit harder. After that, vaccination played a decisive role in precluding the next wave of death and hospitalisation cases when the Delta VOC was introduced in Brazil, between May 2021 and July 2021 (**Fig. 1**).

If the vaccination roll-out had started with moderate acceleration (i.e. 4 weeks faster), it would have reduced even more hospitalisations and deaths than it actually did in the >80 age group during the Gamma wave (**Fig. 2**). It would have also decreased the number of deaths in the 70 - 79 age group during the Gamma wave. When we considered a highly accelerated vaccine deployment (i.e. starting 8 weeks faster), the number of deaths would have been reduced by approximately 45%, 50%, and 40% of the observed number that occurred during the peak of the VOC Gamma wave in the 60 - 69, 70 - 79, and >80 age groups, respectively.

**Figure 2:**
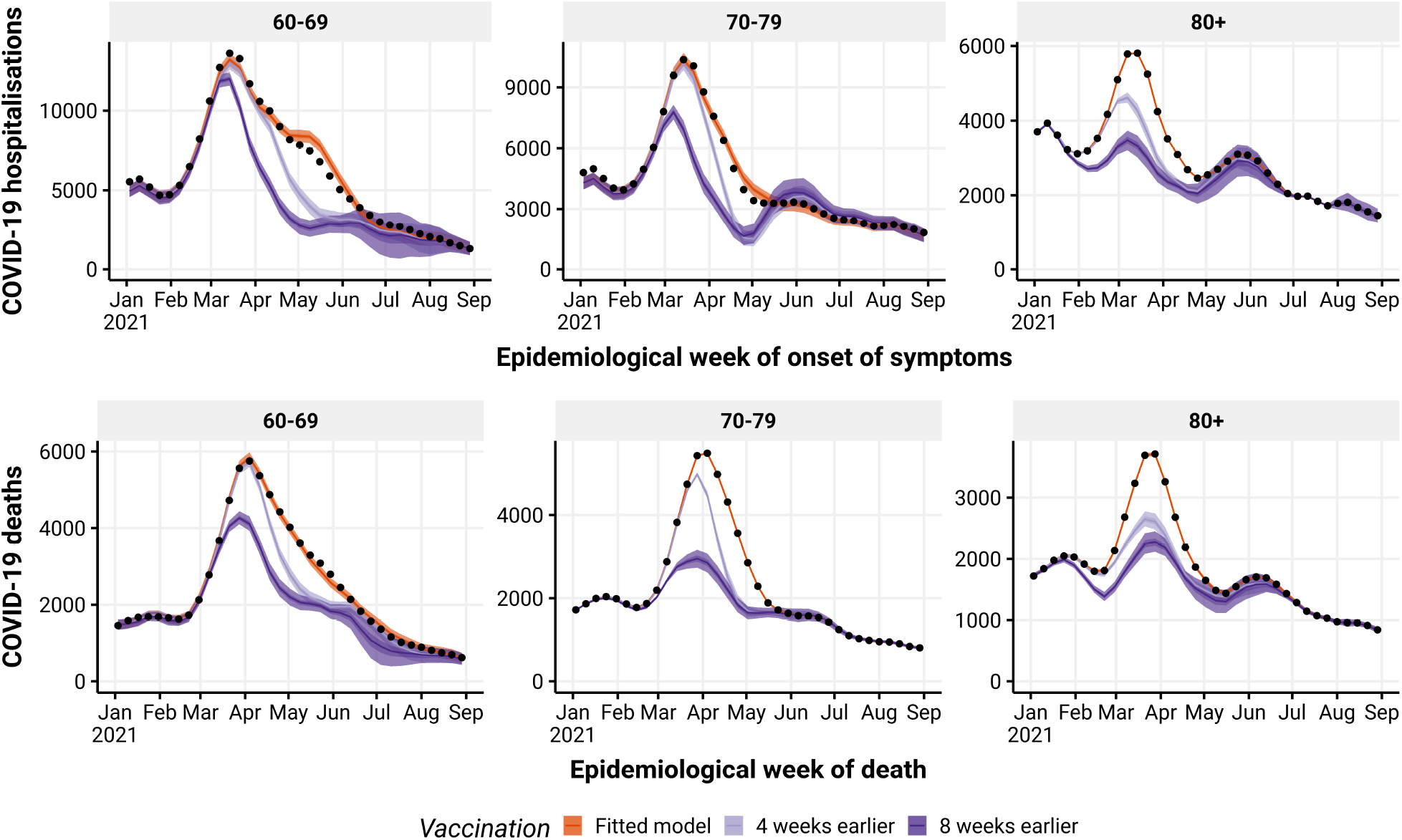
Estimated number of hospitalisations (top) and deaths (bottom) due to COVID-19 by epidemiological week with the realised (dark orange), 4 (light purple) and 8 (dark purple) weeks faster vaccination roll-out, by age group (panels). The observed number of hospitalisations and deaths are given by the black dots. Check legend for grey scale. Line: median, shadow: 95% CI.

If we compare the cumulative number of hospitalisations and deaths between January 1, 2021, and August 29, 2021, with the counterfactual scenario without vaccination, we estimated that vaccination against COVID-19 directly accounted for reducing at least 167,914 (95% CI: 158,206 - 178,298) hospitalisations, and 58,644 (95% CI: 53,785 - 63,952) deaths in the older adults age group. These figures increased to 220,676 (95% CI: 205,975 - 235,407) and to 81,569 (95% CI: 74,262 - 89,162), respectively, if we assume that the same vaccines had a moderate acceleration in the vaccination roll-out. Finally, if vaccination had a high acceleration in the beginning, the number of hospitalisations and deaths averted would increase to 272,421 (95% CI: 251,698 - 292,869) and 105,532 (95% CI: 94,728 - 116,809), respectively (see **Table 1**). The small overlap between the probability distribution curve of the hypothetical scenario of moderate acceleration and the real scenario curve illustrates how significant the aversion would have been in the number of deaths and hospitalisations by starting vaccination earlier, for all age groups studied (**Fig. 3**). The difference between the real and the hypothetical scenario with high acceleration is even more remarkable, evidenciated by the little overlap between the distributions for the different age groups (**Fig. 3**).

**Table 1:** Estimated reductions in hospitalisations/deaths by age group and vaccine roll-out. 60+ is the aggregate of all age groups of older adults.

| Outcome | Age Group | Vaccine roll-out | Estimated reduction (95% CI) |
| --- | --- | --- | --- |
| Hospitalisations | 60+ | Realised | 167,914 (158,206-178,298) |
|  |  | Moderate acceleration | 220,676 (205,975-235,407) |
|  |  | High acceleration | 272,421 (251,698-292,869) |
|  | 60-69 | Realised | 69,174 (62,857-75,217) |
|  |  | Moderate acceleration | 97,241 (87,806-107,003) |
|  |  | High acceleration | 125,279 (110,356-141,121) |
|  | 70-79 | Realised | 56,027 (49,299-62,501) |
|  |  | Moderate acceleration | 70,028 (60,884-78,786) |
|  |  | High acceleration | 85,223 (72,410-96,597) |
|  | 80+ | Realised | 42,916 (37,639-48,082) |
|  |  | Moderate acceleration | 53,597 (46,588-60,582) |
|  |  | High acceleration | 62,134 (53,572-70,724) |
| Deaths | 60+ | Realised | 58,644 (53,785-63,952) |
|  |  | Moderate acceleration | 81,569 (74,262-89,162) |
|  |  | High acceleration | 105,532 (94,728-116,809) |
|  | 60-69 | Realised | 19,686 (17,213-22,134) |
|  |  | Moderate acceleration | 29,512 (25,108-33,776) |
|  |  | High acceleration | 40,357 (31,979-48,155) |
|  | 70-79 | Realised | 19,183 (16,283-22,138) |
|  |  | Moderate acceleration | 25,957 (21,758-30,173) |
|  |  | High acceleration | 34,283 (28,488-39,713) |
|  | 80+ | Realised | 19,791 (16,202-23,051) |
|  |  | Moderate acceleration | 26,212 (21,554-30,837) |
|  |  | High acceleration | 30,917 (24,991-36,755) |

**Figure 3:**
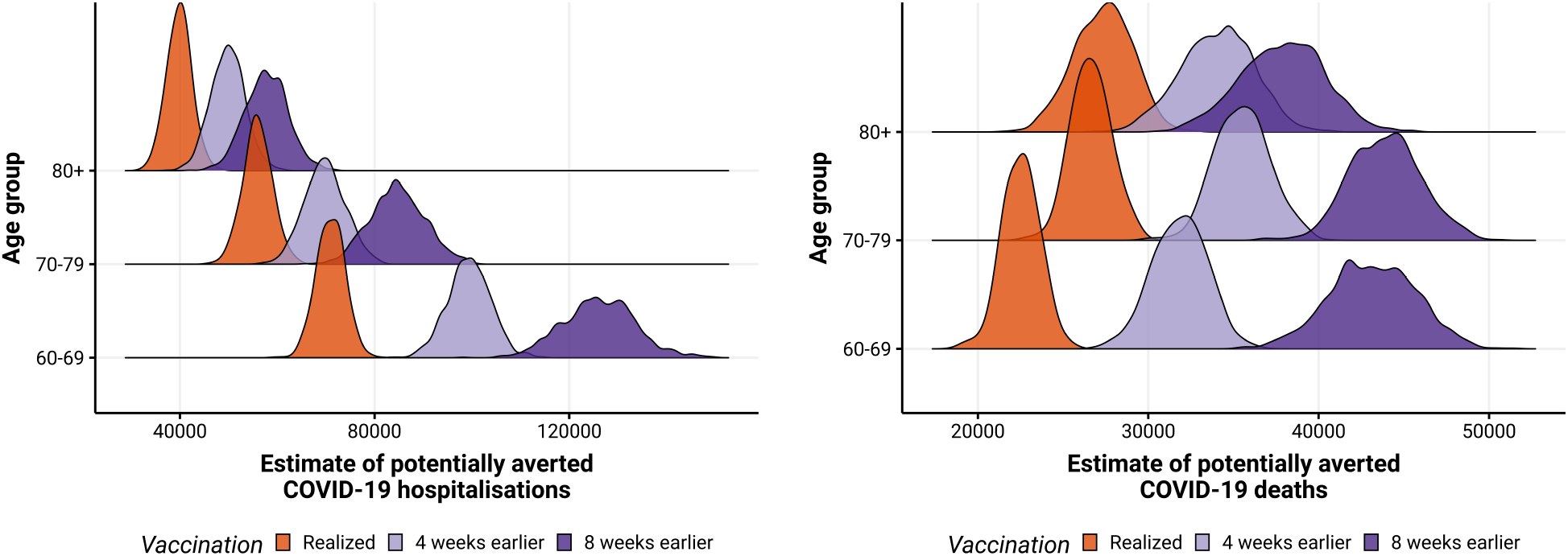
Posterior distribution of hospitalisations (left) and deaths (right) due to COVID-19 potentially averted by vaccination between January 1, 2021, and August 29, 2021, by age group, with the realised (dark orange), 4 (light purple) and 8 weeks faster (dark purple) vaccination roll-out. Check legend for grey scale.

## DISCUSSION

In this work, we used a Bayesian model approach to estimate the number of hospitalisations and deaths in the older adults population (above 60 y.o.) due to COVID-19 in Brazil, under three different scenarios of vaccination roll-outs: the real one; starting immunisations with moderate acceleration (4 weeks faster); and starting immunisations with high acceleration (8 weeks faster). These numbers were compared to a putative scenario of no vaccinations. By assuming that the risks in the target group (older adults) relative to a reference group (20 - 29 age group) decrease as vaccination coverage increases, we accurately fit the decrease in the number of hospitalisations and deaths in the target group. In our analysis, we assumed that exposure to infection was constant in each age group over the analysed period. Still, changes in the relative exposure risks among age groups may have occurred. For instance, young adults may have increased their mobility because of employment issues, which would overestimate averted hospitalisations and deaths under the assumptions of time homogeneity. On the other hand, if the older adults increased their mobility because they felt confident after they got the vaccine shots, our estimated figures would be underestimated. As we do not have access to information on how relative exposure risks among age groups differ over time, we kept homogeneous mobility over time as the most parsimonious assumption.

We estimated that vaccinations in Brazil averted more than 167,000 COVID-19 hospitalisations of individuals above 60 y.o. between January and August 2021, a decrease of 35% compared to the scenario with no vaccination. Furthermore, if we consider the mean cost of US$12,000.00 per hospital admission,^25^ Brazil saved about US$2 billion in health care as a direct effect of vaccination, which is equivalent to what the country spent on vaccination in the same period (US$2.2 billion).^26^ An additional 104,000 individuals above 60 y.o. might not have been hospitalised if the immunisation had started with high acceleration. We also estimated that more than 58,000 lives of older adults were saved in the period analysed, a 35% decrease in deaths that would occur between March and August in the scenario without vaccination. A further 47 thousand lives could have been saved if the Brazilian government started the immunisation with high acceleration, i.e., at least 20% of the actual deaths of >60 y.o. individuals during the period analysed could have been avoided. It is important to note that, although the Brazilian vaccination campaign officially commenced January 18, 2021,^14^ coverage by a second dose in the population above 80 y.o. only reached the level of about 50% nationwide by the end of March.^16^ For those 60 - 69 y.o., it only reached above 50% by the end of the first semester.^16^ Since those age groups were mostly vaccinated with CoronaVac,^27,16^ with an interval of 2 - 4 weeks between doses at that time, the moderate and high accelerated vaccination scenarios are not unrealistic ones, since population level impacts are only significant after at least a significant part of the target population is immunised.

In the next paragraphs, we discuss three points that allow us to state that our estimates are a lower bound for the saved lives in the most critical period of the COVID-19 epidemic in Brazil: (*i*) our model estimated only the direct effects of vaccination, therefore no herd immunity and no secondary morbidity or mortality effects were considered; (*ii*) we performed analysis considering only the most vulnerable age population (>60), which accounts for 46.4% of hospitalisations and 67.6% of deaths in Brazil during the period analysed; and (*iii*) we considered the exact same pace of vaccination in our hypothetical faster roll-out scenarios that were performed, which is very slow if compared to the speed capacity and organization Brazil had during previous mass vaccinations.^8,9^

Vaccination can reduce hospitalisations and deaths via three main (direct and indirect) effects: reducing the severity of the disease in infected individuals; reducing the susceptibility to infection of vaccinated individuals; and reducing the transmission potential of vaccinated individuals that do get infected, mostly by shortening the period which viral shedding is high.^28,29,30^ Our counterfactual scenarios assumed that vaccination affected hospitalisation and death only in the target age group (>60), but not in the reference age group (20 - 29 y.o.), since this age group remained unvaccinated in the period studied. This assumption means that we did not account for the curbing of infections caused by reducing susceptibility and transmission. Population effects, such as those affecting the transmission, are not included here, and, therefore, this model is always expected to underestimate averted hospitalisations and deaths. Additionally, with fewer hospitalisations, the healthcare system could provide better services and potentially increase the survival of individuals with severe COVID-19, however, this effect was not accounted for in our model. High healthcare burden substantially affected in-hospital mortality, especially during peaks and in regions with fragile health infrastructures.^2^ Overlooking this effect also leads to an underestimation of the vaccination effect on deaths.

Our estimates were restricted to age groups over 60 y.o., and we made this choice for two main reasons: the National Immunisation Plan prioritised an order of vaccination from older age groups towards younger age groups, so that for the period analysed, most of the vaccination effort had been directed towards these age groups. Vaccination of younger age groups by age criterion (excluding health workers, individuals with certain medical conditions, among others), in turn, only started after July in most states. The second reason is that age groups above 60 y.o. represent the highest risk of hospitalisation and mortality, accounting for 46.4% of hospitalisations and 67.6% of deaths in Brazil during the period analysed. Therefore, the choice to focus on these age groups reduced the estimates of the number of averted hospitalisations and casualties since it is also affected by the age pyramid distribution in each state.

Another hypothetical scenario could be to explore a different pace of vaccination, compatible with the Brazilian capacity to organise mass vaccinations. In the past, Brazil was capable of achieving the oral immunisation of nearly 20 million children against polio in a single day.^31,32^ In the 2010 mass vaccination against Influenza, Brazil vaccinated more than 80% of the target group, corresponding to 89 million people, during the seasonal campaign.^8,9^ Recent local and national experiences with yellow fever vaccination also indicate the country has the organisation and structure to make fast massive campaigns to control epidemics,^33^ which, for a variety of reasons, was not the case with the COVID-19 vaccination. Brazil completed 250,000 doses per day between February and March 2021, increasing to an average of 500,000 doses per day in the period between April and May 2021, and reaching a pace of above 1 million doses per day only in June 2021.^16^ If the Brazilian government had used all its capacity to organise the COVID-19 campaign, one could expect more significant reduction in deaths and hospitalisations than the figures estimated in this work.

On the other tail of age groups (children), Brazil faces a similar problem to the one analysed here. The BNT162b2 pediatric vaccine was approved by the Brazilian Health Regulatory Agency (ANVISA) in December 16, 2021,^34^ and the starting date of the pediatric vaccination was a month later – officially on January 14, 2022 in a slow rate of delivery.^35,16^ Contradictorily, Brazil has high pediatric hospitalisation and mortality from COVID-19. For the 5 - 11 age group, COVID-19 was the cause of 3,302 and 3,317 hospitalisations in 2020 and 2021 (until November 29, 2021), respectively,^18^ whereas 156 and 142 deaths occurred in 2020 and 2021 (until November 29, 2021), respectively, as a consequence of COVID-19.^18^ Additionally, deaths due to SARI consequences were 450 and 292 in 2020 and 2021 (until November 29, 2021), respectively.^18^ In order to compare the magnitude of these figures related to SARI, one can look at the leading mortality cause of children in the 5 - 9 age group in Brazil; disregarding external causes (such as violence), nervous system diseases and neoplasms, the greatest cause of mortality between 2015 and 2019 was the sum of all respiratory system diseases, which in average caused 283 deaths per year.^36^ These numbers evidence the significant role of COVID-19 alone in child mortality. Based on the results presented in this work, in which we observed the direct effect of lives saved, compared to a scenario with no vaccination, we hypothesise that postponing children’s vaccination led to avoidable suffering and deaths. Therefore, future studies similar to ours will be needed to estimate the number of children’s lives Brazil has sacrificed due to unexplained delays in vaccine deployment.

## Supporting information

Additional results

## Data Availability

All data and code used in this work is publicly available at https://github.com/covid19br/
bayes-vacina-paper.

https://github.com/covid19br/bayes-vacina-paper

## Contributors

Conceptualization – LSF, LSB, MEB and RLPS. Methodology – LSF, LSB and PIP. Software – LSF. Validation – PIP. Formal Analysis – LSF. Investigation – LSF. Data Curation – MEB and RLPS. Writing - Original Draft – LSF, FMDM and RLPS. Writing - Review & Editing – All authors. Visualization – MFCG and LSF.

## Data Availability Statement

All data and code used in this work is publicly available at https://github.com/covid19br/bayes-vacina-paper, we also provide a repository with raw data in.^37^

## Ethical Approval Statement

Ethics approval was not required for this study because data were obtained from the Influenza’s Epidemiological Surveillance Information System (SIVEP-Gripe), which is publicly available by the Brazilian National Health System. According to the National Ethical Commission (CONEP) of the National Health Council, Resolution Number 510 of April 7, 2016 (http://conselho.saude.gov.br/resolucoes/2016/Reso510.pdf), no ethical approval is needed.

## Acknowledgements

All authors thank the members of Observatório COVID-19 BR for their insightful discussion of the results of this work. Specially, we would like to thank Verônica Coelho, Maria Amélia Veras, Brigina Kemp, Maria Rita Donalísio, Lorena Barberia, Flávia Ferrari, José Cassio de Moraes, and Guilherme Werneck for their comments.

