## Additional results for "Estimating the impact of implementation and timing of the COVID-19 vaccination programme in Brazil: a counterfactual analysis"

This supplementary material contains the times series of the pandemic in Brazil, the model selection for data fitting, a graphical measurement of fitting quality, a figure showing the timing when vaccination reached perceptible coverage in each state and age group, a figure of vaccine coverage during the period analysed and the graphical results for individual states. The tabulated results for each state is available in our repository at https://github.com/covid19br/bayes-vacina-paper together with the code used to generate the results.

### 1 COVID-19 pandemic and variants in Brazil

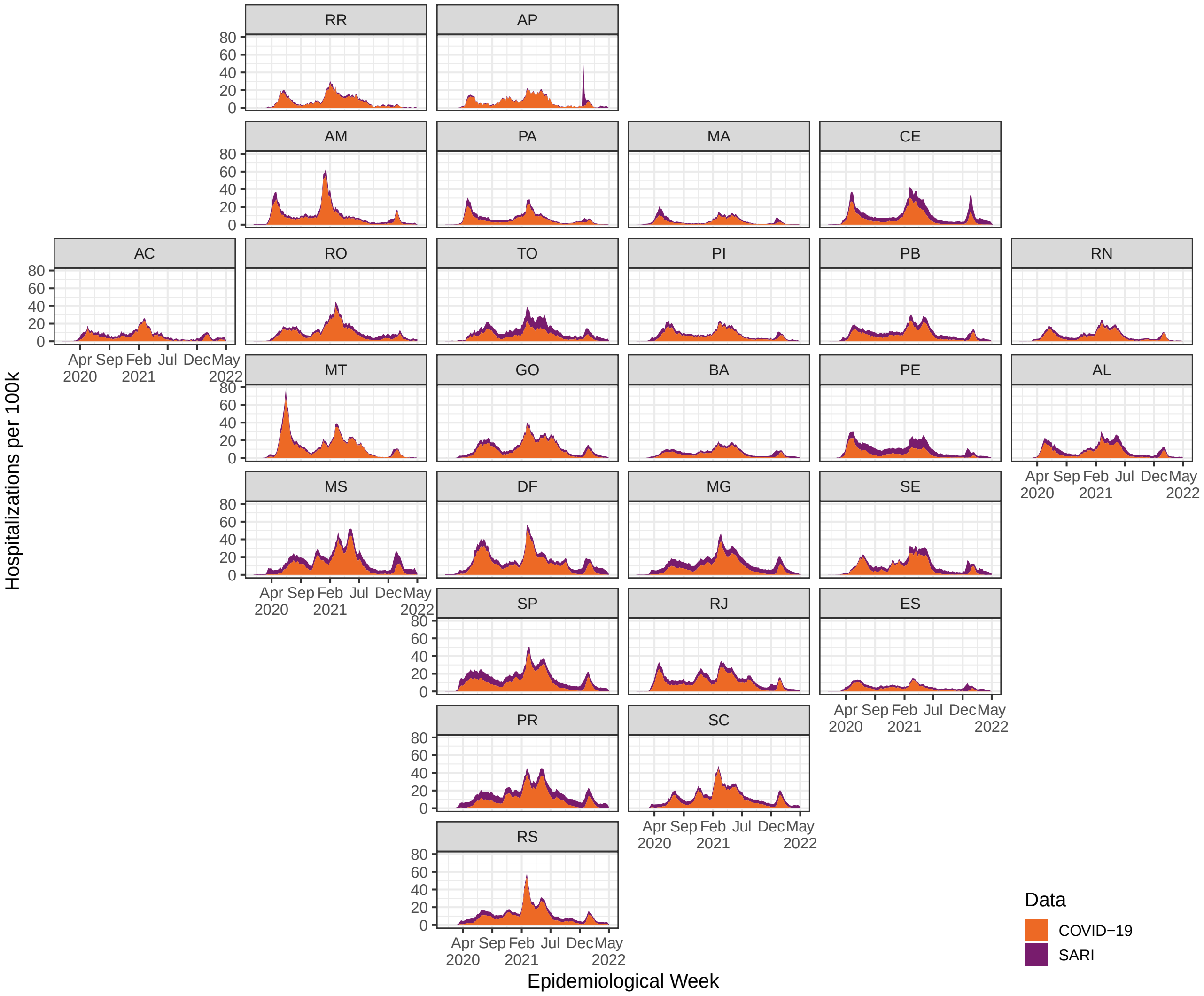


**Figure S1:** Weekly hospitalisations by date of first symptoms per 100k individuals by Severe Acute Respiratory Infection (SARI) and COVID-19, per state. SARI hospitalisations include COVID-19 hospitalisations. Last four weeks should be ignored due to notification delay. Source: SIVEP-Gripe.

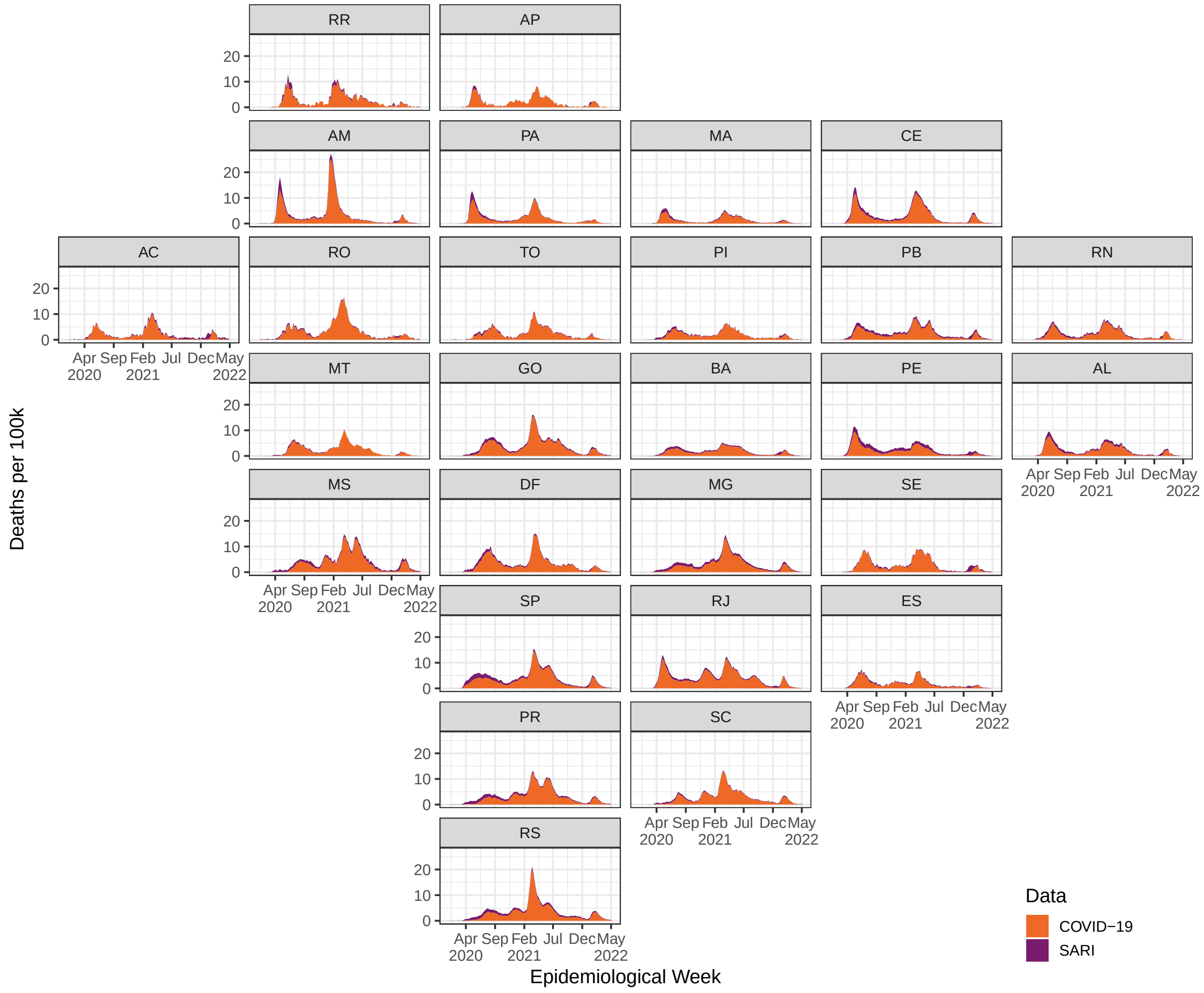


**Figure S2:** Weekly deaths by date of death per 100k individuals by Severe Acute Respiratory Infection (SARI) and COVID-19, per state. SARI deaths include COVID-19 deaths. Last four weeks should be ignored due to notification delay. Source: SIVEP-Gripe,

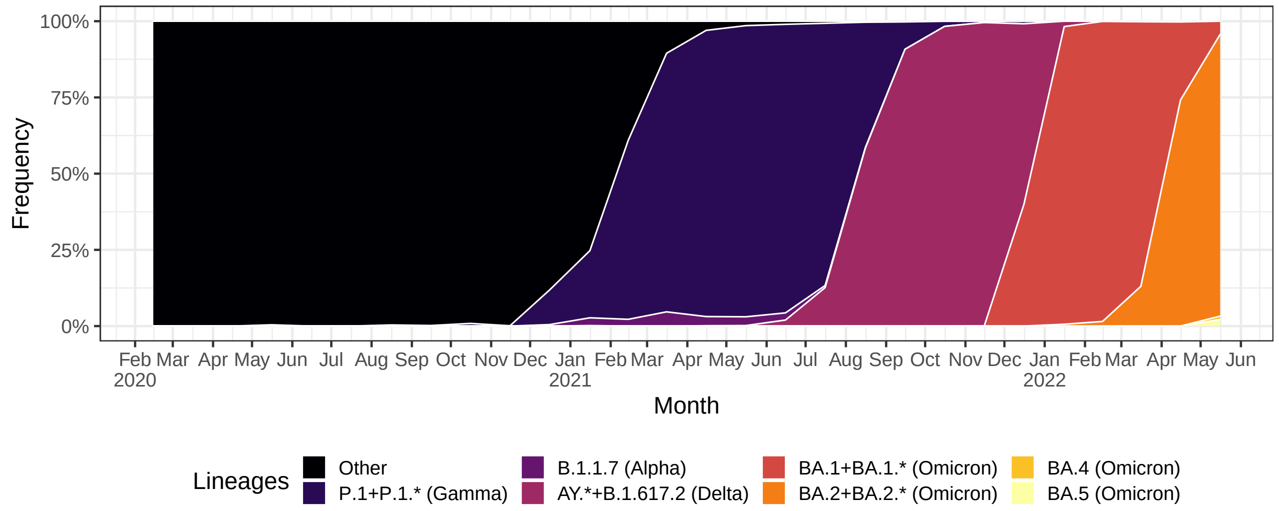


**Figure S3:** Frequency of SARS-CoV-2 variants in Brazil over time, simplified to consider only VOC. Source: https://www.genomahcov.fiocruz.br/.

### 2 Model Selection

To investigate if considering an autorregressive process may cause overfitting in the model, we did model comparison using the Watanabe-Akaike Information Criterion (WAIC). Besides the fixed effects in the model, we considered a model with no random effects (noeffect), and three random effects models: i.i.d, random walk of order 1 (rw1), autorregressive process of order 1 (ar1).

Our model can be generically written as:

$$
\begin{aligned}
Y^{(t)} &\sim \text{Normal}(\mu^{(t)}, \sigma), \quad \sigma > 0, \quad t = 1, 2, \ldots \\
\mu^{(t)} &= \beta_0 + \beta_1 X^{(t)} + \sum_v \beta_v X^{(t)} \text{c}_{Y,v}^{(t)} + \gamma^{(t)}
\end{aligned} \tag{1}
$$

where $\gamma^{(t)}$ denotes the random effect, if used.

The prior distributions for the fixed effects are:

$$
\begin{aligned}
p(\beta_0) &\propto 1, \\
\beta_1 &\sim \text{Normal}(0, 0.001), \\
\beta_v &\sim \text{Normal}(0, 0.001), \\
\sigma &\sim \text{Gamma}(1, 0.00001),
\end{aligned} \tag{2}
$$

The random effect models and prior distributions to their parameters are given in table below:

| Model | Formula | Priors |
|---|---|---|
| No effect | - | - |
| i.i.d | $\gamma^{(t)} = \varepsilon_i$<br>$\varepsilon_i \sim \text{Normal}(0, \theta^{-1})$ | $\log(\theta) \sim \text{Gamma}(2, 1e-4)$ |
| rw1 | $\gamma^{(t)} = \Delta x_i$<br>$\Delta x_1 = x_i - x_{i-1} \sim \text{Normal}(0, \theta^{-1})$ | $\log(\theta) \sim \text{Gamma}(2, 1e-4)$<br>except SP, where<br>$\log(\theta) \sim \text{Gamma}(1, 100)$ |
| ar1 | $\gamma^{(t)} = \rho x_{t-1} + \varepsilon_t$<br>$x_1 \sim \text{Normal}(0, (\tau(1-\rho^2))^{-1})$<br>$\varepsilon_t \sim \text{Normal}(0, \tau^{-1})$ | $\log\left(\frac{1+\rho}{1-\rho}\right) \sim \text{Normal}(0, 0.15)$<br>$\log(\tau(1-\rho^2)) \sim \text{Gamma}(1, 100)$<br>except RS, where<br>$\log(\tau(1-\rho^2)) \sim \text{Gamma}(2, 100)$ |

We consider the relative likelihood of a model as given by:

$$
RL = \exp[-(WAIC - WAIC_{min})/2] \tag{3}
$$

where $WAIC_{min}$ denotes the lower $WAIC$ between the four models, computed separately for each state, outcome, and age group. The results are displayed in Figure S4.

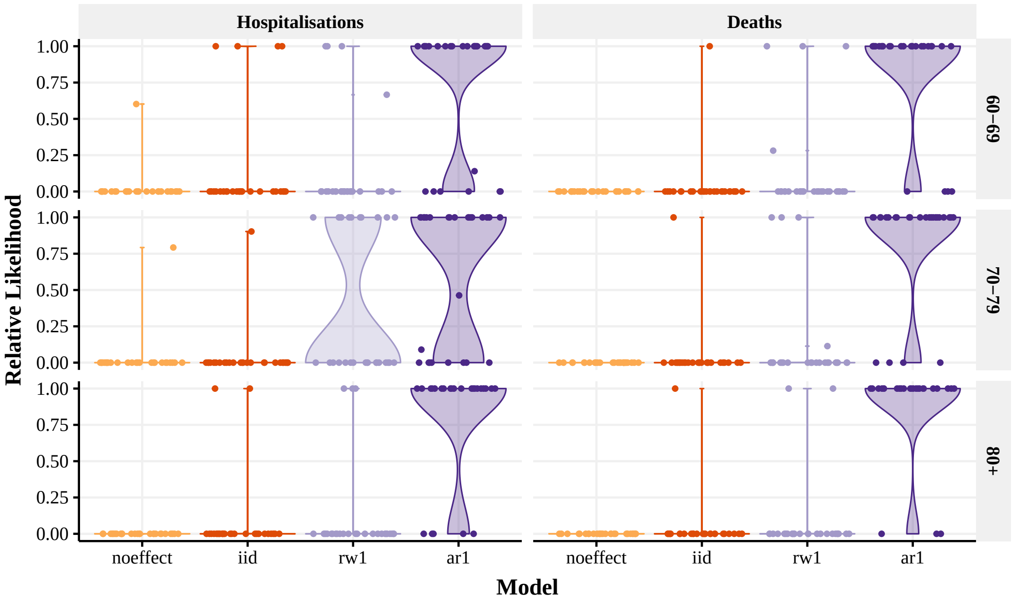


**Figure S4:** Relative likelihood as function of model, outcome, and age group. Each state (points) has the relative likelihood calculated independently. The higher, the better. The violin plot width displays the distribution of the points.

### 3 Fitting quality

**Table S1:** Name codes and their respective states

| Name Code | State |
|---|---|
| North Region | |
| AC | Acre |
| AM | Amazonas |
| AP | Amapá |
| PA | Pará |
| RR | Roraima |
| RO | Rondônia |
| TO | Tocantins |
| Northeast Region | |
| AL | Alagoas |
| BA | Bahia |
| CE | Ceará |
| MA | Maranhão |
| PB | Paraíba |
| PE | Pernambuco |
| PI | Piauí |
| RN | Rio Grande do Norte |
| SE | Sergipe |
| Southeast Region | |
| ES | Espírito Santo |
| MG | Minas Gerais |
| RJ | Rio de Janeiro |
| SP | São Paulo |
| Center-West Region | |
| DF | Distrito Federal |
| GO | Goiás |
| MS | Mato Grosso do Sul |
| MT | Mato Grosso |
| South Region | |
| PR | Paraná |
| RS | Rio Grande do Sul |
| SC | Santa Catarina |

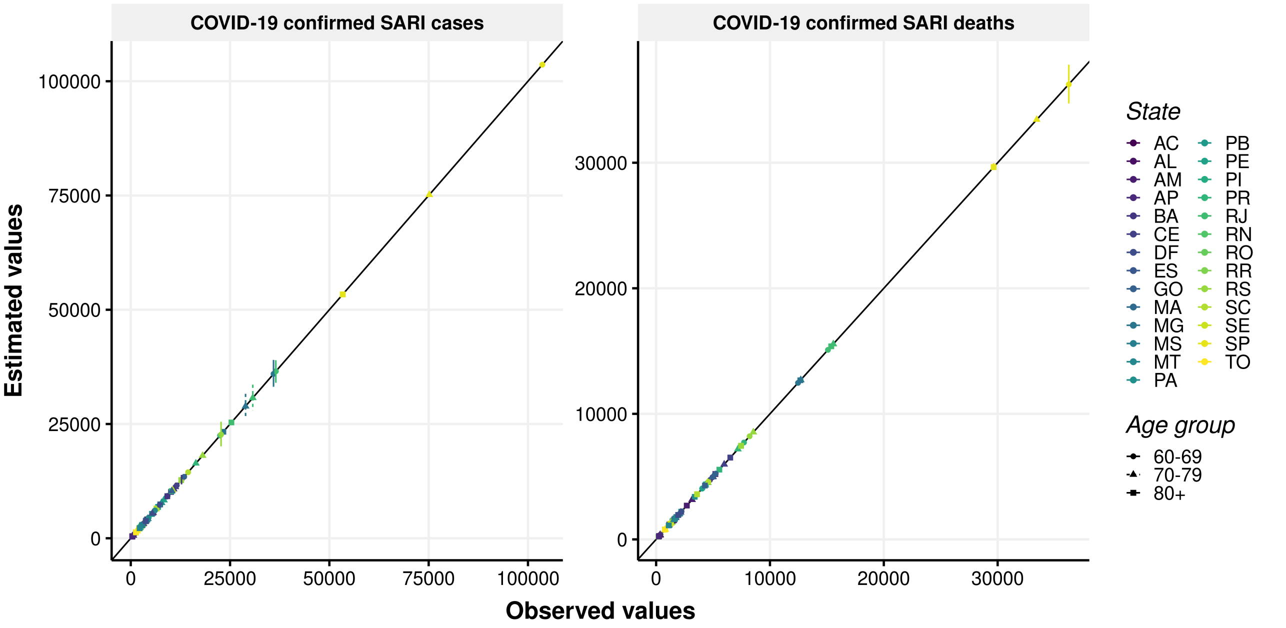


**Figure S5:** Estimated value of outcome (hospitalisation on the left and deaths on the right) as a function of the observed value, per state (color) and age group (point type). A black line with 1:1 relationship is plotted for reference.

### 4 Vaccine coverage

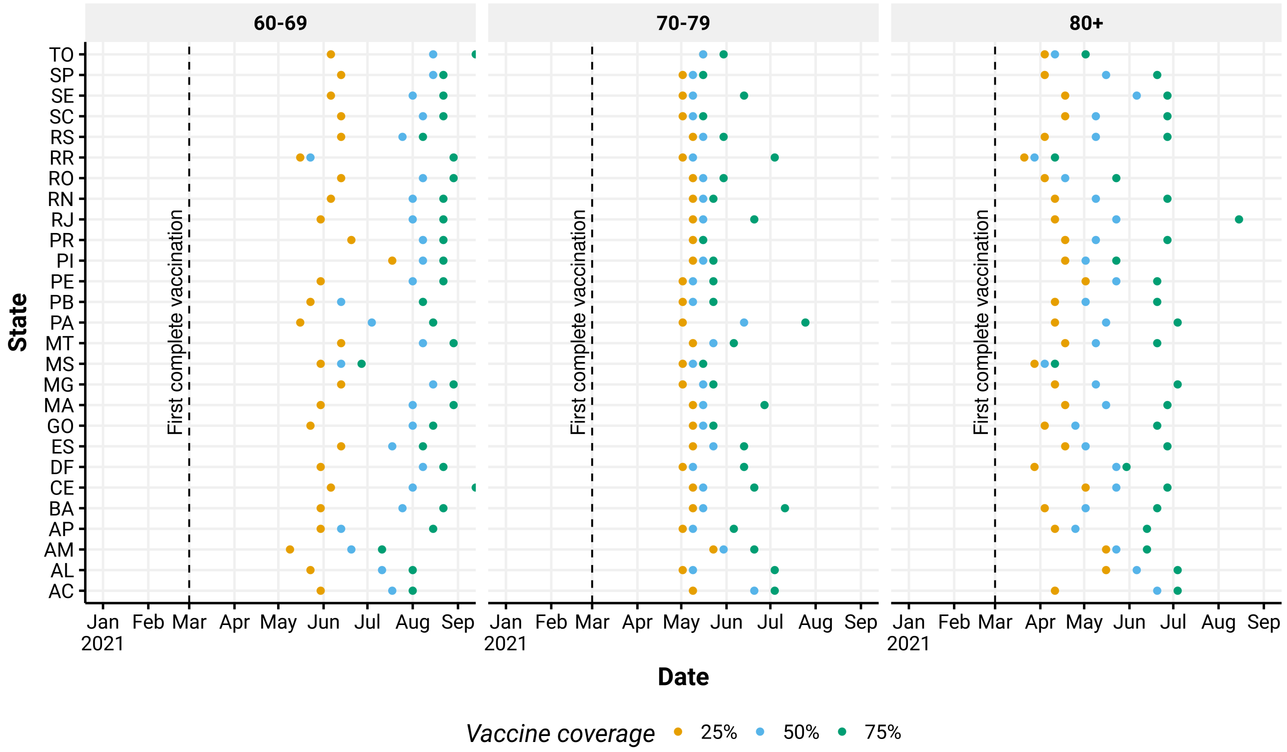


**Figure S6:** Week (x-axis) in which different vaccine coverage thresholds were reached (colors), by state (y axis) for the analysed age groups (panels). The dashed line denotes the day that the first vaccination could be completed considering the official start as January 17, 2021 (first dose + 28 days + second dose + 14 days, as the first vaccines were CoronaVac).

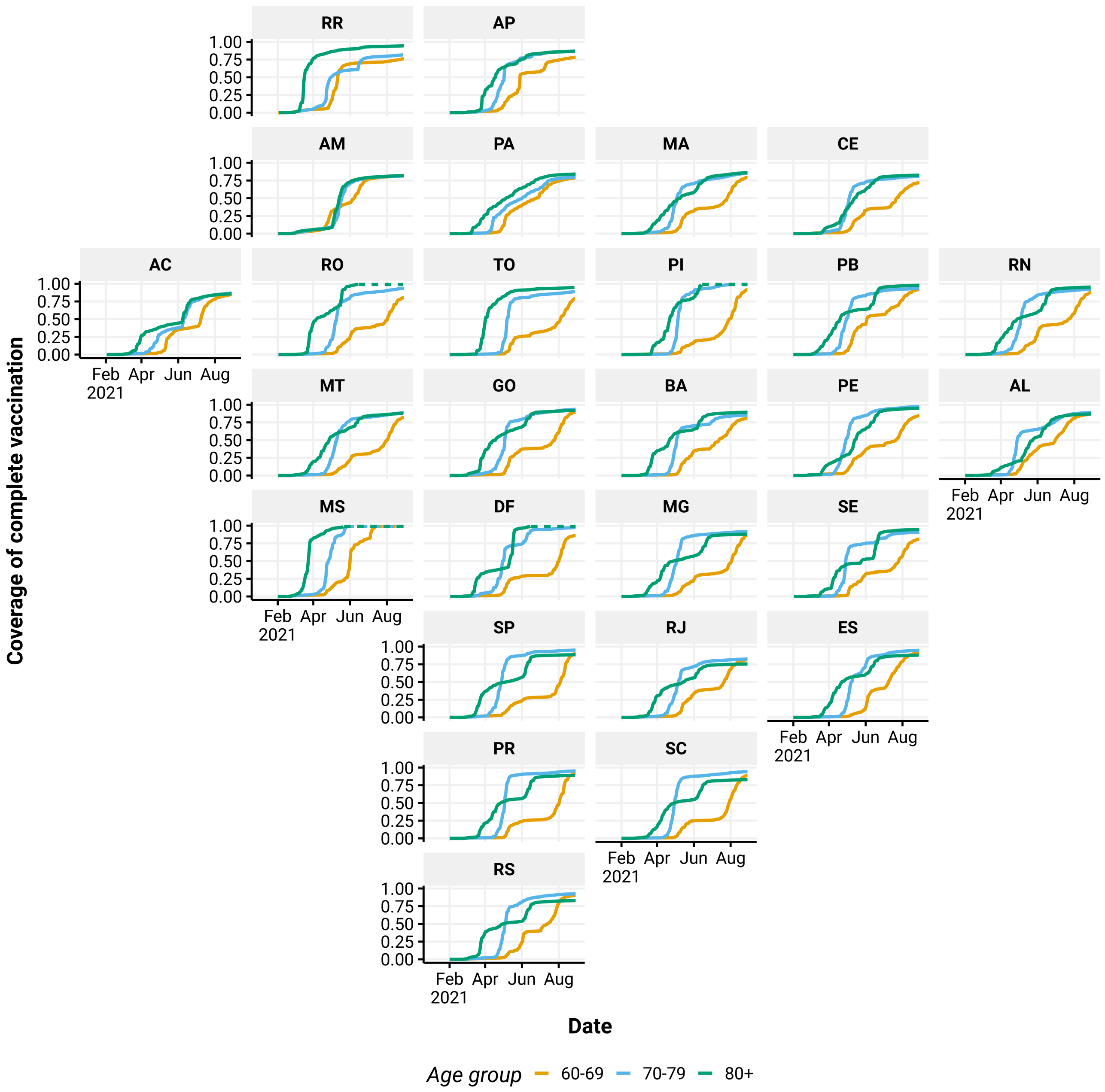


**Figure S7:** Vaccine coverage per age group. Due to imprecise estimates of population size, some age groups surpassed 100% coverage and these cases are denoted with a dashed line.

### 5 Results

#### 5.1 Sensitivity analysis per age group of reference

**Table S2:** Number of hospitalisations and deaths averted by COVID-19 vaccination, by timing of vaccination rollout, using 30-39 age group as age of reference.

| | | Vaccination | | |
|---|---|---|---|---|
| Outcome | Agegroup | Realised | Moderate acceleration | High acceleration |
| Hospitalisations | 60+ | 160,958 (151,753-170,325) | 208,845 (195,093-222,588) | 249,089 (226,295-272,062) |
| | 60-69 | 66,732 (61,185-72,493) | 91,820 (82,659-101,392) | 110,147 (92,141-128,504) |
| | 70-79 | 52,419 (46,745-57,905) | 63,744 (56,107-71,568) | 76,278 (64,862-87,499) |
| | 80+ | 41,807 (37,273-46,410) | 53,281 (47,148-59,406) | 62,664 (54,471-70,892) |
| Deaths | 60+ | 65,386 (61,109-69,569) | 92,410 (85,526-99,286) | 122,811 (111,729-134,059) |
| | 60-69 | 23,542 (21,606-25,477) | 36,621 (32,829-40,270) | 52,635 (44,147-60,925) |
| | 70-79 | 21,664 (19,187-24,130) | 28,695 (25,005-32,473) | 37,756 (31,994-43,506) |
| | 80+ | 20,179 (17,478-22,969) | 27,094 (23,248-30,963) | 32,420 (27,327-37,390) |

**Table S3:** Number of hospitalisations and deaths averted by COVID-19 vaccination, by timing of vaccination rollout, using 40-49 age group as age of reference.

| | | Vaccination | | |
|---|---|---|---|---|
| Outcome | Age group | Realised | Moderate acceleration | High acceleration |
| Hospitalisations | 60+ | 147,041 (138,619-155,472) | 196,445 (182,678-210,730) | 239,072 (215,128-263,310) |
| | 60-69 | 61,013 (56,600-65,572) | 88,357 (80,238-96,376) | 111,536 (92,998-128,795) |
| | 70-79 | 46,647 (41,536-51,777) | 57,286 (49,220-65,508) | 67,598 (55,526-80,508) |
| | 80+ | 39,382 (34,900-44,152) | 50,803 (44,638-57,513) | 59,938 (51,857-68,886) |
| Deaths | 60+ | 63,780 (59,257-68,362) | 90,551 (83,309-97,716) | 121,254 (107,444-134,433) |
| | 60-69 | 21,358 (19,592-23,111) | 34,478 (30,767-38,299) | 51,730 (41,770-62,499) |
| | 70-79 | 20,743 (18,236-23,048) | 27,382 (23,358-31,156) | 35,657 (29,244-41,692) |
| | 80+ | 21,679 (18,323-25,192) | 28,691 (24,164-33,682) | 33,867 (27,963-40,347) |

#### 5.2 Results by Brazilian states

**Table S4:** Number of hospitalisations and deaths averted by COVID-19 vaccination, by state, by timing of vaccination rollout.

| | | | Vaccination | | |
|---|---|---|---|---|---|
| State | Outcome | Age group | Realised | Moderate acceleration | High acceleration |
| AC | Hospitalisations | 60+ | 132 (53-208) | 198 (89-304) | 285 (128-432) |
| | | 60-69 | 46 (7-85) | 74 (15-130) | 110 (29-192) |
| | | 70-79 | 43 (-5-93) | 62 (-2-130) | 90 (1-189) |
| | | 80+ | 43 (-1-83) | 63 (3-118) | 86 (4-158) |
| | Deaths | 60+ | -2 (-39-39) | -5 (-58-49) | -8 (-79-65) |
| | | 60-69 | 1 (-21-23) | 1 (-27-28) | 0 (-38-41) |
| | | 70-79 | 3 (-21-30) | 1 (-35-37) | 2 (-48-51) |
| | | 80+ | -7 (-28-12) | -8 (-38-20) | -10 (-51-26) |
| AL | Hospitalisations | 60+ | 944 (437-1,453) | 1,386 (586-2,221) | 1,898 (662-3,181) |

| | | | | | |
|---|---|---|---|---|---|
| | | 60-69 | 417 (83-738) | 699 (91-1,279) | 1,047 (42-2,014) |
| | | 70-79 | 370 (66-652) | 551 (65-1,004) | 745 (36-1,427) |
| | | 80+ | 157 (-100-439) | 136 (-204-494) | 106 (-311-552) |
| | Deaths | 60+ | 33 (-129-190) | 65 (-187-315) | 116 (-249-482) |
| | | 60-69 | 33 (-48-113) | 60 (-81-202) | 95 (-145-347) |
| | | 70-79 | -7 (-108-86) | -8 (-177-150) | -3 (-231-217) |
| | | 80+ | 6 (-94-106) | 13 (-117-140) | 24 (-124-174) |
| AM | Hospitalisations | 60+ | 1,750 (732-2,866) | 2,360 (1,012-3,788) | 3,164 (1,340-5,098) |
| | | 60-69 | 560 (-149-1,354) | 776 (-129-1,736) | 1,033 (-152-2,276) |
| | | 70-79 | 667 (12-1,329) | 886 (25-1,740) | 1,189 (26-2,386) |
| | | 80+ | 523 (107-970) | 697 (144-1,291) | 942 (177-1,759) |
| | Deaths | 60+ | 693 (382-984) | 989 (551-1,400) | 1,435 (790-2,036) |
| | | 60-69 | 251 (57-419) | 359 (82-599) | 508 (117-847) |
| | | 70-79 | 251 (74-414) | 356 (109-591) | 518 (151-852) |
| | | 80+ | 192 (34-350) | 274 (45-500) | 408 (71-743) |
| AP | Hospitalisations | 60+ | 79 (-86-231) | 115 (-154-363) | 165 (-219-550) |
| | | 60-69 | 73 (-34-179) | 127 (-70-317) | 194 (-140-510) |
| | | 70-79 | -19 (-93-58) | -28 (-141-88) | -38 (-202-127) |
| | | 80+ | 24 (-56-104) | 17 (-97-130) | 9 (-133-156) |
| | Deaths | 60+ | 9 (-31-48) | 15 (-44-74) | 25 (-59-109) |
| | | 60-69 | 7 (-17-30) | 12 (-22-48) | 19 (-30-73) |
| | | 70-79 | 5 (-16-24) | 8 (-23-37) | 11 (-34-55) |
| | | 80+ | -3 (-27-21) | -4 (-41-32) | -4 (-54-44) |
| BA | Hospitalisations | 60+ | 4,290 (2,804-5,674) | 6,081 (3,637-8,400) | 7,937 (4,052-11,803) |
| | | 60-69 | 1,393 (496-2,294) | 2,298 (460-4,045) | 3,325 (-1-6,431) |
| | | 70-79 | 1,268 (497-2,025) | 1,682 (444-2,941) | 2,089 (131-4,047) |
| | | 80+ | 1,629 (845-2,507) | 2,100 (1,044-3,260) | 2,523 (1,188-3,950) |
| | Deaths | 60+ | 707 (261-1,163) | 1,113 (386-1,849) | 1,585 (484-2,745) |
| | | 60-69 | 149 (-53-359) | 290 (-132-724) | 485 (-295-1,240) |
| | | 70-79 | 54 (-172-271) | 102 (-272-472) | 166 (-409-719) |
| | | 80+ | 505 (155-847) | 720 (214-1,207) | 933 (245-1,627) |
| CE | Hospitalisations | 60+ | 3,539 (1,941-5,197) | 5,600 (2,576-8,697) | 7,688 (2,537-12,933) |
| | | 60-69 | 1,217 (191-2,066) | 2,248 (95-4,179) | 3,426 (-743-7,418) |
| | | 70-79 | 1,201 (281-2,054) | 1,802 (148-3,345) | 2,351 (-247-4,920) |
| | | 80+ | 1,121 (165-2,120) | 1,550 (-81-3,207) | 1,912 (-528-4,424) |
| | Deaths | 60+ | 1,962 (1,101-2,741) | 3,364 (1,861-4,732) | 4,743 (2,465-6,877) |
| | | 60-69 | 558 (225-892) | 1,114 (421-1,818) | 1,738 (551-2,947) |
| | | 70-79 | 565 (104-1,015) | 983 (161-1,805) | 1,387 (177-2,587) |
| | | 80+ | 839 (235-1,413) | 1,267 (283-2,267) | 1,618 (155-3,059) |
| DF | Hospitalisations | 60+ | 3,446 (2,778-4,114) | 4,444 (3,567-5,335) | 5,587 (4,445-6,724) |
| | | 60-69 | 1,633 (1,191-2,089) | 2,104 (1,523-2,699) | 2,712 (1,896-3,505) |
| | | 70-79 | 1,139 (749-1,515) | 1,484 (963-1,967) | 1,899 (1,238-2,522) |
| | | 80+ | 674 (377-980) | 856 (483-1,241) | 977 (556-1,416) |
| | Deaths | 60+ | 540 (278-786) | 781 (401-1,143) | 1,094 (556-1,613) |
| | | 60-69 | 216 (50-378) | 309 (73-543) | 460 (110-808) |
| | | 70-79 | 208 (54-360) | 314 (80-547) | 447 (114-776) |
| | | 80+ | 116 (-28-244) | 157 (-40-330) | 187 (-56-398) |
| ES | Hospitalisations | 60+ | 488 (218-756) | 710 (306-1,111) | 965 (378-1,554) |
| | | 60-69 | 92 (-76-264) | 134 (-135-387) | 200 (-226-603) |
| | | 70-79 | 166 (-11-330) | 248 (-24-500) | 358 (-37-716) |
| | | 80+ | 230 (100-362) | 328 (134-521) | 407 (152-660) |
| | Deaths | 60+ | 208 (87-351) | 325 (126-565) | 459 (121-848) |
| | | 60-69 | 27 (-44-105) | 41 (-84-172) | 70 (-158-315) |

| | | | | | |
|---|---|---|---|---|---|
| | | 70-79 | 59 (-22-144) | 101 (-37-246) | 159 (-59-387) |
| | | 80+ | 121 (50-187) | 183 (71-284) | 229 (63-373) |
| GO | Hospitalisations | 60+ | 8,306 (6,679-9,987) | 10,267 (8,163-12,323) | 12,269 (9,744-14,794) |
| | | 60-69 | 3,183 (2,113-4,246) | 4,216 (2,874-5,635) | 5,275 (3,548-7,066) |
| | | 70-79 | 2,781 (1,738-3,731) | 3,309 (2,013-4,507) | 3,921 (2,370-5,438) |
| | | 80+ | 2,341 (1,617-3,057) | 2,743 (1,877-3,596) | 3,073 (2,070-4,054) |
| | Deaths | 60+ | 1,732 (1,149-2,348) | 2,389 (1,528-3,273) | 3,177 (2,002-4,396) |
| | | 60-69 | 660 (343-955) | 972 (508-1,416) | 1,392 (713-2,083) |
| | | 70-79 | 481 (97-878) | 643 (109-1,194) | 879 (141-1,648) |
| | | 80+ | 591 (253-913) | 775 (306-1,233) | 906 (327-1,465) |
| MA | Hospitalisations | 60+ | 1,910 (1,016-2,825) | 2,636 (1,189-4,129) | 3,401 (1,051-5,726) |
| | | 60-69 | 823 (464-1,191) | 1,244 (458-2,035) | 1,721 (126-3,360) |
| | | 70-79 | 487 (-115-1,139) | 689 (-337-1,708) | 899 (-605-2,350) |
| | | 80+ | 600 (61-1,098) | 703 (-21-1,411) | 781 (-152-1,635) |
| | Deaths | 60+ | 822 (476-1,167) | 1,078 (559-1,607) | 1,354 (583-2,147) |
| | | 60-69 | 264 (95-431) | 372 (70-652) | 479 (-38-994) |
| | | 70-79 | 240 (37-442) | 313 (29-597) | 409 (20-799) |
| | | 80+ | 319 (83-548) | 393 (56-720) | 466 (8-901) |
| MG | Hospitalisations | 60+ | 25,670 (21,430-29,877) | 33,594 (27,900-39,661) | 40,953 (33,103-49,412) |
| | | 60-69 | 9,005 (6,590-11,615) | 13,265 (9,285-17,417) | 17,469 (11,341-23,861) |
| | | 70-79 | 10,010 (7,942-12,027) | 12,153 (9,742-14,643) | 14,107 (11,139-17,230) |
| | | 80+ | 6,655 (4,205-9,089) | 8,176 (4,843-11,526) | 9,377 (5,219-13,529) |
| | Deaths | 60+ | 6,887 (4,843-8,804) | 9,872 (6,671-12,983) | 12,907 (8,355-17,338) |
| | | 60-69 | 2,043 (1,085-3,010) | 3,299 (1,465-5,024) | 4,730 (1,615-7,533) |
| | | 70-79 | 2,063 (977-3,100) | 2,906 (1,318-4,370) | 3,878 (1,738-5,841) |
| | | 80+ | 2,781 (1,281-4,103) | 3,667 (1,459-5,786) | 4,300 (1,348-7,142) |
| MS | Hospitalisations | 60+ | 913 (190-1,668) | 1,133 (89-2,235) | 1,266 (-712-3,246) |
| | | 60-69 | 407 (-49-833) | 640 (-162-1,399) | 773 (-460-1,909) |
| | | 70-79 | 315 (-120-739) | 359 (-246-928) | 386 (-532-1,372) |
| | | 80+ | 192 (-206-584) | 134 (-356-637) | 107 (-1,129-1,380) |
| | Deaths | 60+ | 414 (161-651) | 642 (267-1,019) | 822 (295-1,365) |
| | | 60-69 | 231 (89-372) | 412 (159-670) | 543 (134-980) |
| | | 70-79 | 153 (16-300) | 211 (18-428) | 273 (9-567) |
| | | 80+ | 30 (-114-179) | 20 (-182-219) | 6 (-255-264) |
| MT | Hospitalisations | 60+ | 340 (-354-1,021) | 745 (-607-1,998) | 1,339 (-943-3,511) |
| | | 60-69 | 252 (-253-729) | 666 (-369-1,630) | 1,284 (-584-3,041) |
| | | 70-79 | -30 (-441-380) | -71 (-848-670) | -125 (-1,394-990) |
| | | 80+ | 119 (-186-423) | 150 (-261-594) | 180 (-353-770) |
| | Deaths | 60+ | 83 (-164-337) | 43 (-332-430) | -49 (-663-558) |
| | | 60-69 | 35 (-91-171) | -10 (-267-242) | -102 (-646-442) |
| | | 70-79 | -6 (-153-160) | -10 (-210-209) | -16 (-299-295) |
| | | 80+ | 54 (-94-203) | 63 (-137-258) | 69 (-179-310) |
| PA | Hospitalisations | 60+ | 4,965 (3,794-6,156) | 6,623 (4,900-8,330) | 8,236 (5,880-10,612) |
| | | 60-69 | 1,763 (1,028-2,486) | 2,323 (1,418-3,201) | 2,923 (1,734-4,010) |
| | | 70-79 | 1,650 (886-2,370) | 2,259 (1,111-3,376) | 2,899 (1,238-4,480) |
| | | 80+ | 1,552 (898-2,166) | 2,041 (1,111-2,876) | 2,414 (1,205-3,504) |
| | Deaths | 60+ | 1,400 (759-2,055) | 2,089 (1,099-3,096) | 2,842 (1,398-4,359) |
| | | 60-69 | 241 (-73-588) | 373 (-148-914) | 580 (-291-1,441) |
| | | 70-79 | 487 (47-894) | 743 (47-1,370) | 1,069 (26-2,003) |
| | | 80+ | 672 (273-1,065) | 973 (399-1,539) | 1,193 (432-1,922) |
| PB | Hospitalisations | 60+ | 1,569 (1,006-2,169) | 2,167 (1,329-3,049) | 2,746 (1,504-4,082) |
| | | 60-69 | 389 (130-657) | 624 (182-1,082) | 847 (100-1,664) |
| | | 70-79 | 601 (213-977) | 809 (210-1,370) | 1,019 (128-1,848) |

| | | | | | |
|---|---|---|---|---|---|
| | | 80+ | 579 (204-949) | 733 (226-1,241) | 879 (175-1,564) |
| | Deaths | 60+ | 323 (23-624) | 465 (71-873) | 594 (19-1,202) |
| | | 60-69 | 163 (43-273) | 232 (34-427) | 294 (-99-712) |
| | | 70-79 | 160 (-15-329) | 207 (-21-438) | 255 (-53-575) |
| | | 80+ | 0 (-211-218) | 25 (-227-305) | 45 (-252-370) |
| PE | Hospitalisations | 60+ | 1,142 (264-2,039) | 1,905 (380-3,431) | 2,695 (206-5,201) |
| | | 60-69 | 469 (-41-954) | 809 (-188-1,749) | 1,181 (-595-2,869) |
| | | 70-79 | 386 (-158-956) | 608 (-403-1,613) | 827 (-769-2,428) |
| | | 80+ | 287 (-205-772) | 488 (-184-1,103) | 687 (-206-1,482) |
| | Deaths | 60+ | 1,179 (617-1,750) | 1,849 (1,207-2,475) | 2,480 (1,664-3,291) |
| | | 60-69 | 180 (47-311) | 314 (75-554) | 471 (45-865) |
| | | 70-79 | 617 (107-1,106) | 999 (452-1,576) | 1,357 (748-1,978) |
| | | 80+ | 383 (147-609) | 536 (215-856) | 651 (255-1,068) |
| PI | Hospitalisations | 60+ | 939 (543-1,321) | 1,475 (823-2,079) | 2,082 (1,133-3,028) |
| | | 60-69 | 402 (144-650) | 663 (218-1,077) | 963 (268-1,624) |
| | | 70-79 | 304 (113-486) | 466 (157-759) | 661 (188-1,104) |
| | | 80+ | 232 (-5-442) | 346 (-25-686) | 459 (-36-945) |
| | Deaths | 60+ | 88 (-15-193) | 158 (-13-341) | 250 (-18-513) |
| | | 60-69 | 29 (-23-83) | 53 (-41-147) | 86 (-69-239) |
| | | 70-79 | 33 (-27-91) | 56 (-45-156) | 88 (-69-245) |
| | | 80+ | 26 (-49-101) | 49 (-67-163) | 75 (-79-230) |
| PR | Hospitalisations | 60+ | 13,958 (11,634-16,222) | 17,691 (14,722-20,638) | 20,831 (17,081-24,570) |
| | | 60-69 | 6,070 (4,670-7,449) | 8,012 (6,251-9,856) | 9,470 (7,358-11,696) |
| | | 70-79 | 3,671 (2,287-5,118) | 4,638 (2,806-6,500) | 5,550 (3,243-7,965) |
| | | 80+ | 4,217 (3,234-5,234) | 5,041 (3,870-6,263) | 5,810 (4,422-7,219) |
| | Deaths | 60+ | 4,877 (4,026-5,720) | 6,289 (5,082-7,394) | 7,488 (6,029-8,901) |
| | | 60-69 | 1,787 (1,306-2,279) | 2,457 (1,753-3,171) | 2,936 (1,990-3,851) |
| | | 70-79 | 1,683 (1,210-2,169) | 2,107 (1,491-2,732) | 2,530 (1,740-3,305) |
| | | 80+ | 1,408 (969-1,857) | 1,724 (1,129-2,328) | 2,023 (1,253-2,784) |
| RJ | Hospitalisations | 60+ | 14,562 (10,492-18,256) | 18,207 (12,806-22,935) | 21,397 (15,189-27,401) |
| | | 60-69 | 7,042 (4,716-9,441) | 8,943 (6,401-11,496) | 10,806 (7,652-13,794) |
| | | 70-79 | 4,071 (1,841-6,262) | 5,056 (1,889-8,113) | 5,939 (1,828-9,921) |
| | | 80+ | 3,449 (1,023-5,755) | 4,209 (1,109-7,178) | 4,651 (850-8,260) |
| | Deaths | 60+ | 5,884 (3,704-8,171) | 7,763 (5,020-10,787) | 9,485 (6,042-12,944) |
| | | 60-69 | 2,335 (1,268-3,373) | 3,208 (1,912-4,410) | 4,278 (2,849-5,749) |
| | | 70-79 | 1,532 (331-2,735) | 2,130 (416-3,835) | 2,676 (426-4,874) |
| | | 80+ | 2,017 (446-3,545) | 2,426 (402-4,313) | 2,531 (126-4,730) |
| RN | Hospitalisations | 60+ | 1,060 (611-1,459) | 1,644 (845-2,358) | 2,322 (988-3,540) |
| | | 60-69 | 299 (119-461) | 583 (210-938) | 943 (94-1,724) |
| | | 70-79 | 340 (106-582) | 476 (81-870) | 619 (-1-1,258) |
| | | 80+ | 421 (128-695) | 585 (104-1,028) | 759 (93-1,420) |
| | Deaths | 60+ | 390 (243-531) | 603 (362-862) | 818 (451-1,213) |
| | | 60-69 | 69 (15-128) | 137 (25-251) | 187 (9-373) |
| | | 70-79 | 136 (53-222) | 195 (71-324) | 267 (83-454) |
| | | 80+ | 185 (74-294) | 271 (84-448) | 364 (69-636) |
| RO | Hospitalisations | 60+ | 507 (273-725) | 825 (386-1,225) | 1,222 (399-1,990) |
| | | 60-69 | 127 (-16-281) | 228 (-65-539) | 361 (-301-1,023) |
| | | 70-79 | 211 (72-353) | 340 (107-563) | 521 (148-880) |
| | | 80+ | 168 (46-286) | 257 (78-435) | 341 (105-576) |
| | Deaths | 60+ | 191 (85-304) | 336 (148-525) | 554 (215-891) |
| | | 60-69 | 48 (-23-120) | 81 (-47-205) | 134 (-105-366) |
| | | 70-79 | 81 (24-142) | 128 (36-224) | 243 (69-427) |
| | | 80+ | 62 (6-116) | 126 (17-232) | 177 (19-328) |

| | | | | | |
|---|---|---|---|---|---|
| RR | Hospitalisations | 60+ | 88 (-16-195) | 81 (-58-233) | 59 (-143-279) |
| | | 60-69 | 2 (-71-73) | -11 (-118-89) | -42 (-202-112) |
| | | 70-79 | 88 (36-144) | 99 (30-173) | 112 (18-211) |
| | | 80+ | -2 (-58-47) | -6 (-79-64) | -11 (-107-85) |
| | Deaths | 60+ | 11 (-22-48) | 15 (-32-67) | 10 (-60-82) |
| | | 60-69 | 14 (-6-35) | 22 (-10-53) | 24 (-17-64) |
| | | 70-79 | -2 (-23-21) | -6 (-37-26) | -13 (-67-38) |
| | | 80+ | -2 (-20-17) | -1 (-25-23) | 0 (-32-30) |
| RS | Hospitalisations | 60+ | 10,252 (7,885-12,855) | 13,975 (10,097-18,125) | 17,526 (11,617-23,551) |
| | | 60-69 | 2,529 (724-4,325) | 4,151 (1,170-7,265) | 5,652 (1,053-10,194) |
| | | 70-79 | 4,088 (2,660-5,562) | 5,101 (2,822-7,552) | 6,181 (2,838-9,753) |
| | | 80+ | 3,636 (2,474-4,809) | 4,722 (3,059-6,452) | 5,694 (3,510-7,976) |
| | Deaths | 60+ | 3,259 (2,096-4,499) | 4,945 (2,995-7,060) | 6,770 (3,834-9,841) |
| | | 60-69 | 765 (174-1,358) | 1,374 (232-2,534) | 1,970 (-21-4,048) |
| | | 70-79 | 1,326 (708-1,940) | 1,835 (968-2,716) | 2,565 (1,334-3,850) |
| | | 80+ | 1,168 (250-2,028) | 1,736 (199-3,222) | 2,235 (174-4,265) |
| SC | Hospitalisations | 60+ | 8,545 (6,859-10,169) | 11,083 (8,903-13,192) | 13,886 (10,963-16,604) |
| | | 60-69 | 4,034 (3,018-5,017) | 5,351 (4,031-6,611) | 6,773 (5,031-8,451) |
| | | 70-79 | 2,501 (1,457-3,515) | 3,224 (1,853-4,555) | 4,093 (2,342-5,883) |
| | | 80+ | 2,011 (1,194-2,748) | 2,508 (1,509-3,466) | 3,020 (1,747-4,177) |
| | Deaths | 60+ | 2,433 (1,773-3,098) | 3,247 (2,350-4,163) | 4,302 (3,099-5,537) |
| | | 60-69 | 780 (428-1,156) | 1,061 (583-1,578) | 1,440 (739-2,153) |
| | | 70-79 | 735 (332-1,148) | 955 (421-1,499) | 1,298 (559-2,038) |
| | | 80+ | 918 (549-1,299) | 1,231 (721-1,728) | 1,564 (916-2,201) |
| SE | Hospitalisations | 60+ | 528 (252-803) | 964 (459-1,479) | 1,565 (587-2,586) |
| | | 60-69 | 222 (76-386) | 390 (116-680) | 664 (29-1,308) |
| | | 70-79 | 189 (19-364) | 344 (3-696) | 561 (-78-1,200) |
| | | 80+ | 117 (-53-300) | 229 (-74-556) | 340 (-111-827) |
| | Deaths | 60+ | -17 (-125-103) | -24 (-250-229) | -25 (-457-390) |
| | | 60-69 | 1 (-45-46) | 2 (-95-95) | 4 (-174-175) |
| | | 70-79 | 0 (-61-59) | -1 (-107-107) | -2 (-178-173) |
| | | 80+ | -17 (-96-61) | -25 (-196-138) | -27 (-364-297) |
| SP | Hospitalisations | 60+ | 57,772 (51,251-64,534) | 74,391 (64,988-84,359) | 90,304 (76,086-105,041) |
| | | 60-69 | 26,564 (23,100-30,045) | 36,516 (30,901-42,152) | 45,788 (35,961-55,857) |
| | | 70-79 | 19,556 (14,547-24,534) | 23,411 (16,578-30,165) | 28,220 (19,068-37,311) |
| | | 80+ | 11,652 (8,783-14,403) | 14,464 (10,757-18,101) | 16,295 (11,873-20,471) |
| | Deaths | 60+ | 24,395 (21,132-27,824) | 33,029 (28,038-38,112) | 42,010 (34,155-49,997) |
| | | 60-69 | 8,784 (7,083-10,347) | 12,934 (9,740-15,918) | 17,476 (11,508-23,326) |
| | | 70-79 | 8,311 (6,363-10,312) | 10,635 (7,939-13,559) | 13,763 (10,110-17,620) |
| | | 80+ | 7,300 (5,147-9,422) | 9,460 (6,953-11,976) | 10,771 (7,900-13,542) |
| TO | Hospitalisations | 60+ | 424 (192-665) | 513 (193-838) | 563 (101-1,030) |
| | | 60-69 | 193 (74-310) | 231 (43-420) | 240 (-91-568) |
| | | 70-79 | 57 (-62-180) | 77 (-90-257) | 94 (-131-332) |
| | | 80+ | 173 (8-335) | 206 (5-400) | 229 (-11-456) |
| | Deaths | 60+ | 159 (89-230) | 198 (95-300) | 219 (74-363) |
| | | 60-69 | 27 (-5-62) | 13 (-41-71) | -5 (-101-90) |
| | | 70-79 | 30 (-5-64) | 43 (-8-95) | 59 (-13-131) |
| | | 80+ | 103 (56-150) | 142 (77-207) | 165 (90-243) |

#### 5.3 North Region

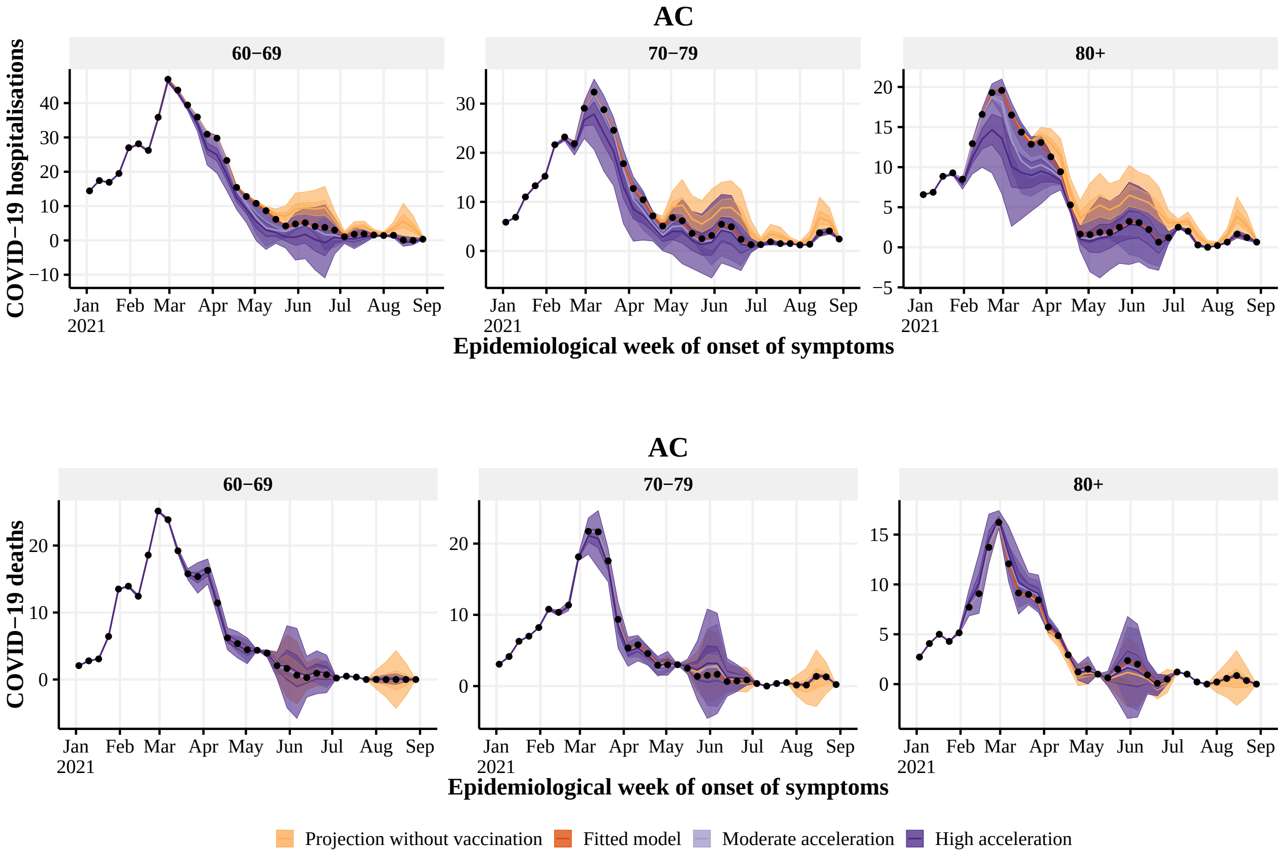


**Figure S8:** Estimated number of hospitalisations (top) and deaths (bottom) by epidemiological week with the realized (dark orange), no vaccination (light orange), 4 (light purple) and 8 (dark purple) weeks earlier vaccination rollout, by age group (panels), in AC state. The observed number of hospitalisations and deaths are given by the black points. With 50% and 95% Credible Intervals.

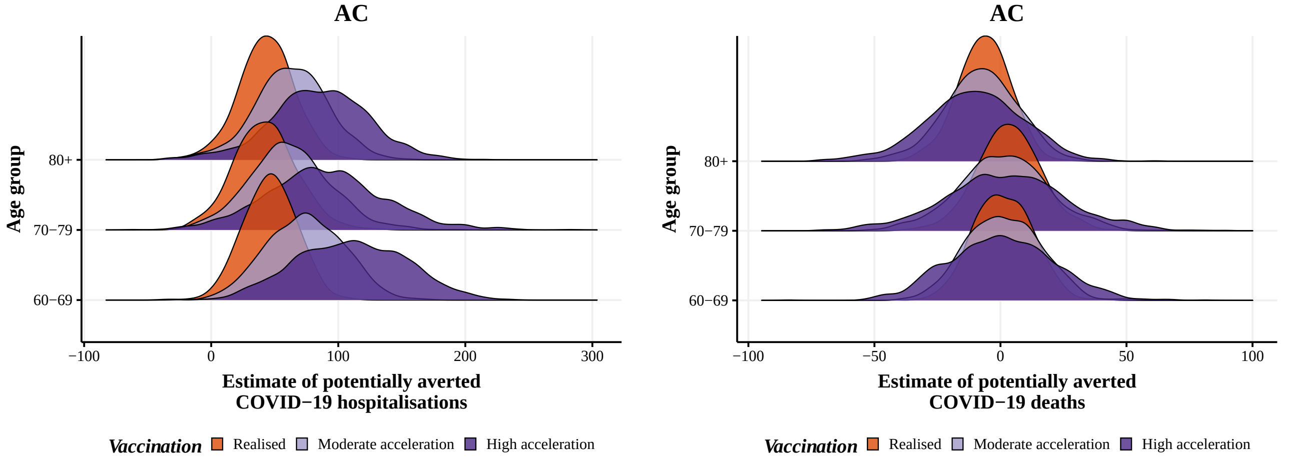


**Figure S9:** Posterior distribution of hospitalisations (left) and deaths (right) potentially averted by vaccination between 2021-01-01 and 2021-08-29 by age group, with the realized (orange), 4 (light purple) and 8 weeks earlier (dark purple) vaccination rollout in AC state.

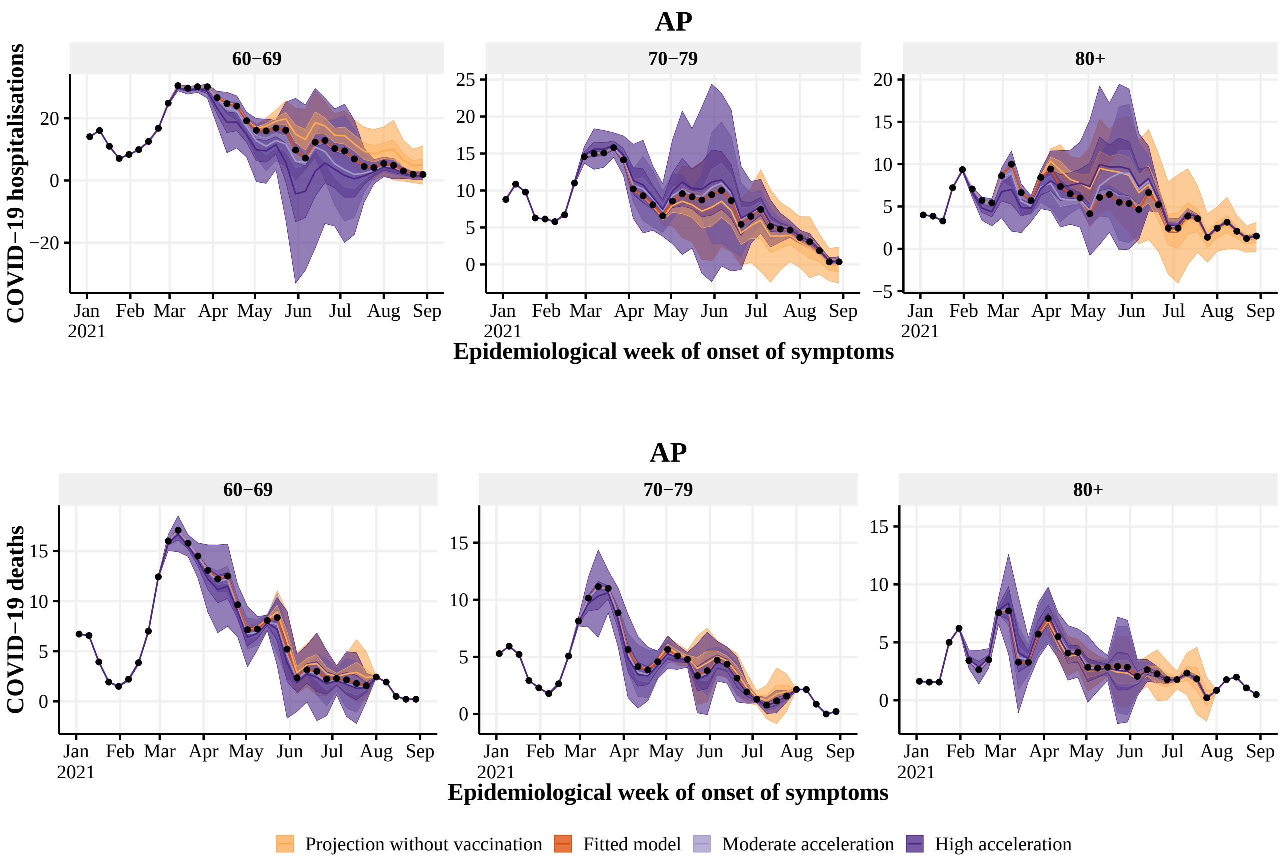


**Figure S10:** Estimated number of hospitalisations (top) and deaths (bottom) by epidemiological week with the realized (dark orange), no vaccination (light orange), 4 (light purple) and 8 (dark purple) weeks earlier vaccination rollout, by age group (panels), in AP state. The observed number of hospitalisations and deaths are given by the black points. With 50% and 95% Credible Intervals.

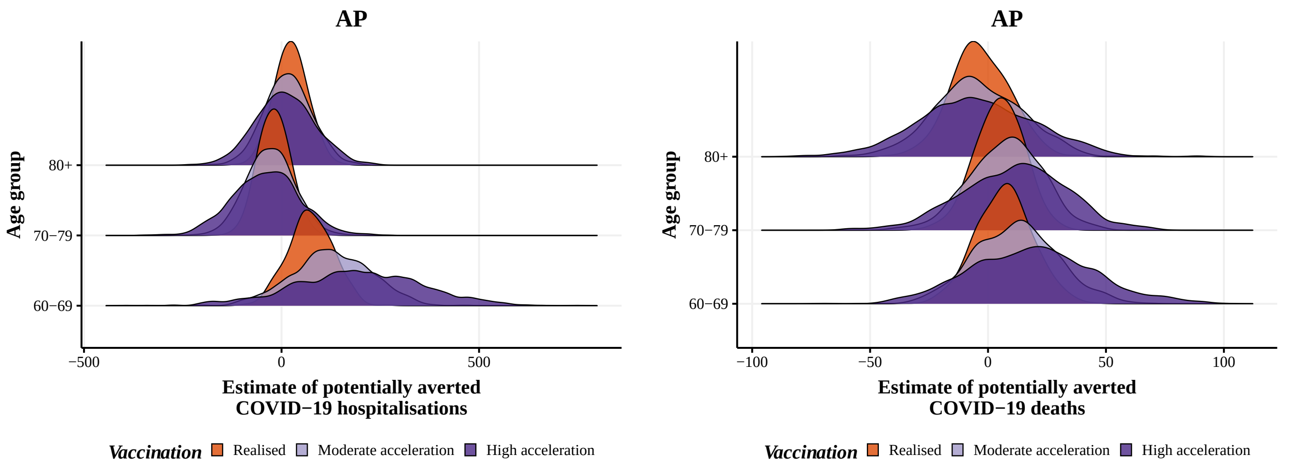


**Figure S11:** Posterior distribution of hospitalisations (left) and deaths (right) potentially averted by vaccination between 2021-01-01 and 2021-08-29 by age group, with the realized (orange), 4 (light purple) and 8 weeks earlier (dark purple) vaccination rollout in AP state.

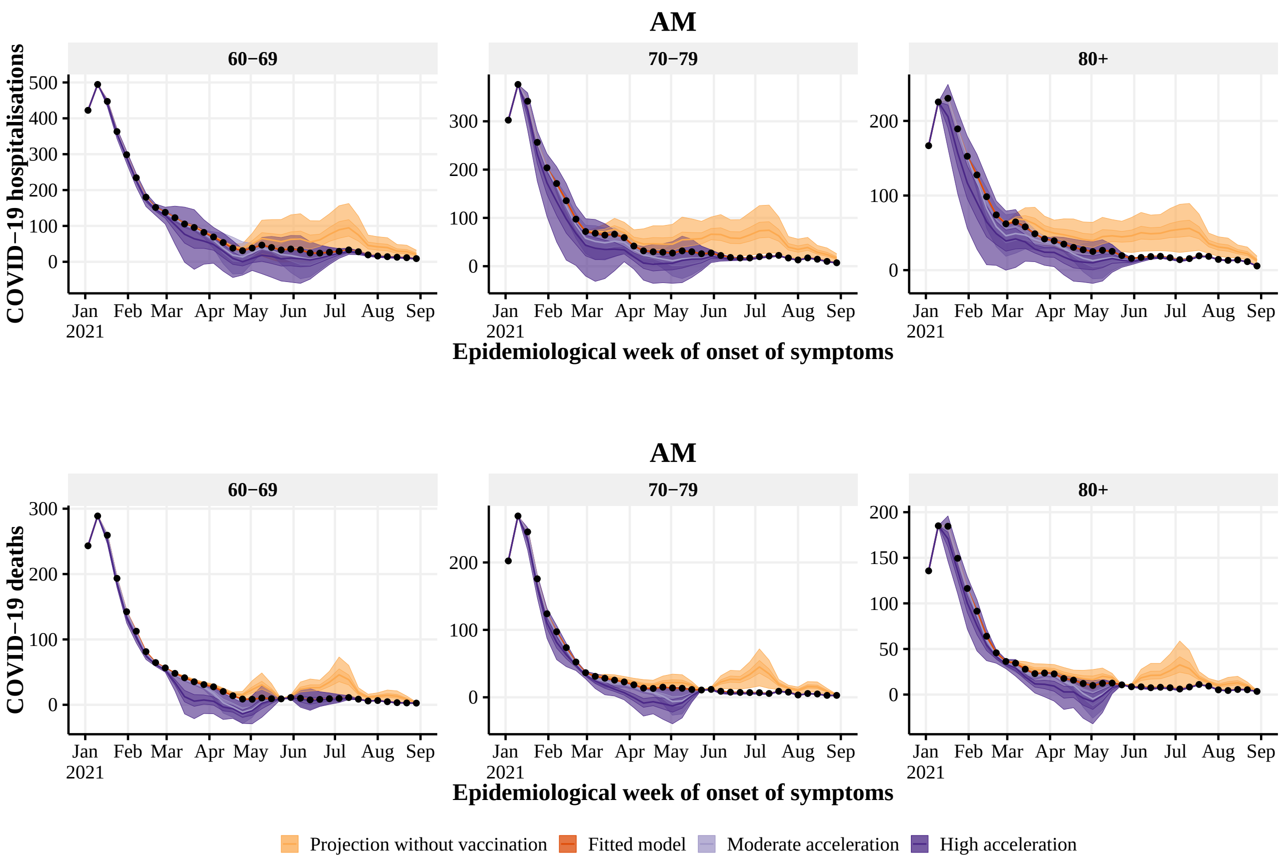


**Figure S12:** Estimated number of hospitalisations (top) and deaths (bottom) by epidemiological week with the realized (dark orange), no vaccination (light orange), 4 (light purple) and 8 (dark purple) weeks earlier vaccination rollout, by age group (panels), in AM state. The observed number of hospitalisations and deaths are given by the black points. With 50% and 95% Credible Intervals.

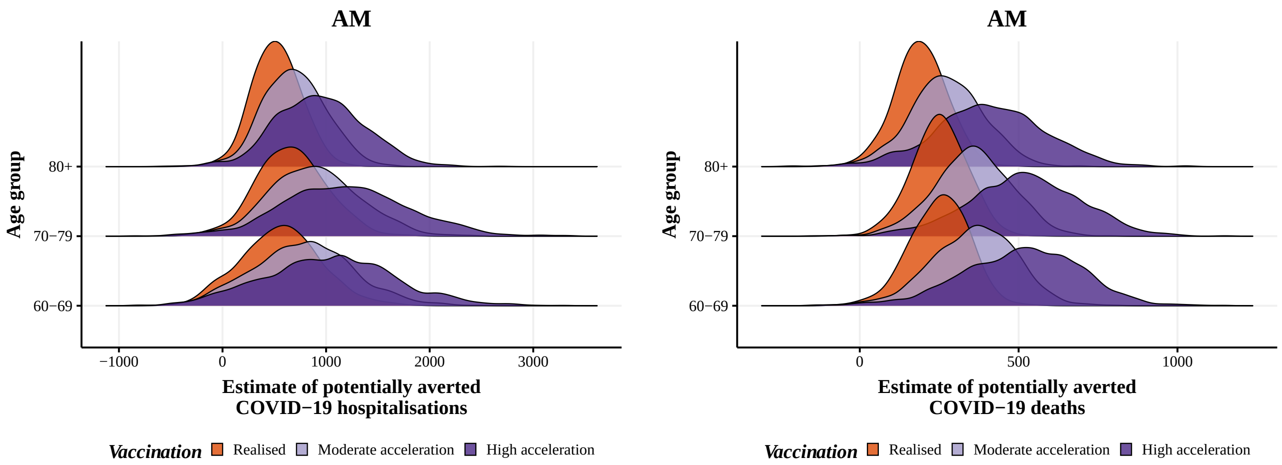


**Figure S13:** Posterior distribution of hospitalisations (left) and deaths (right) potentially averted by vaccination between 2021-01-01 and 2021-08-29 by age group, with the realized (orange), 4 (light purple) and 8 weeks earlier (dark purple) vaccination rollout in AM state.

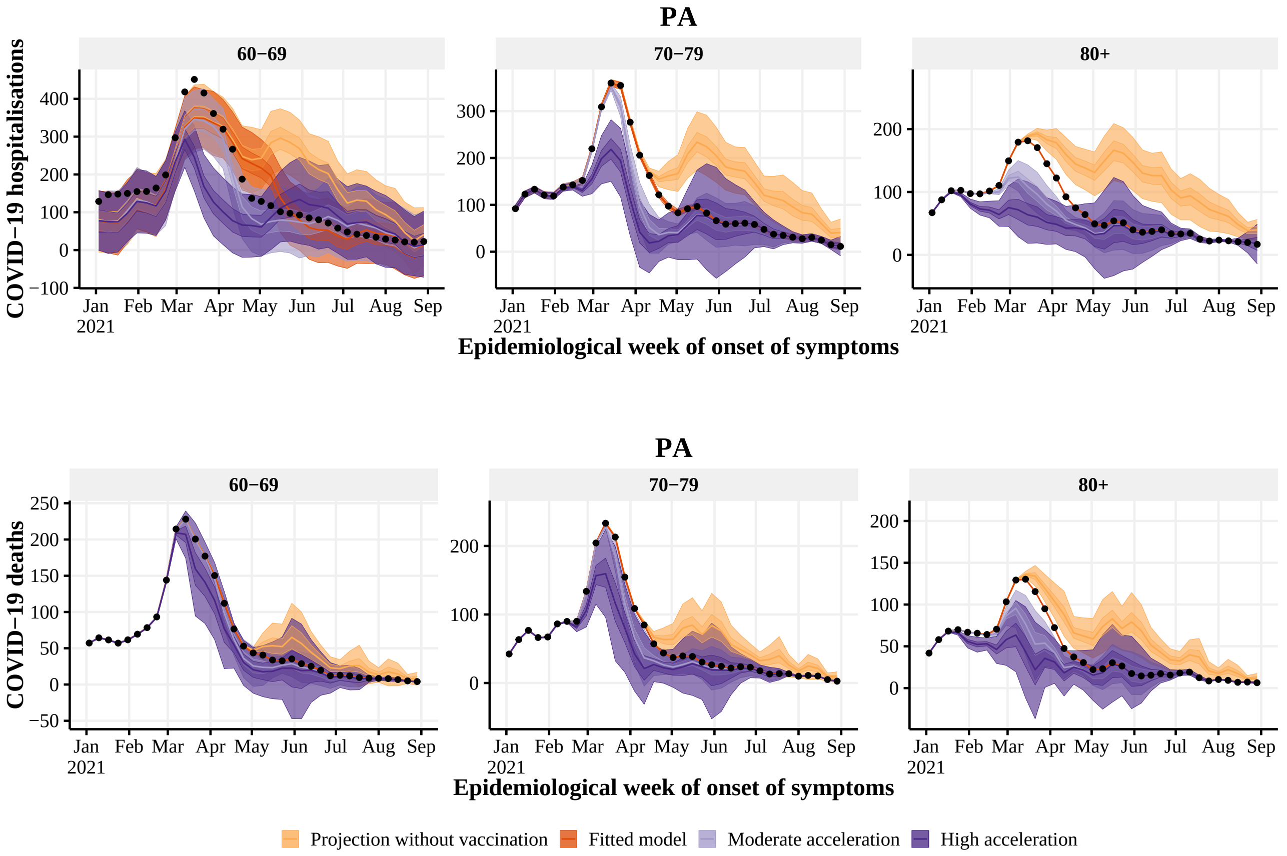


**Figure S14:** Estimated number of hospitalisations (top) and deaths (bottom) by epidemiological week with the realized (dark orange), no vaccination (light orange), 4 (light purple) and 8 (dark purple) weeks earlier vaccination rollout, by age group (panels), in PA state. The observed number of hospitalisations and deaths are given by the black points. With 50% and 95% Credible Intervals.

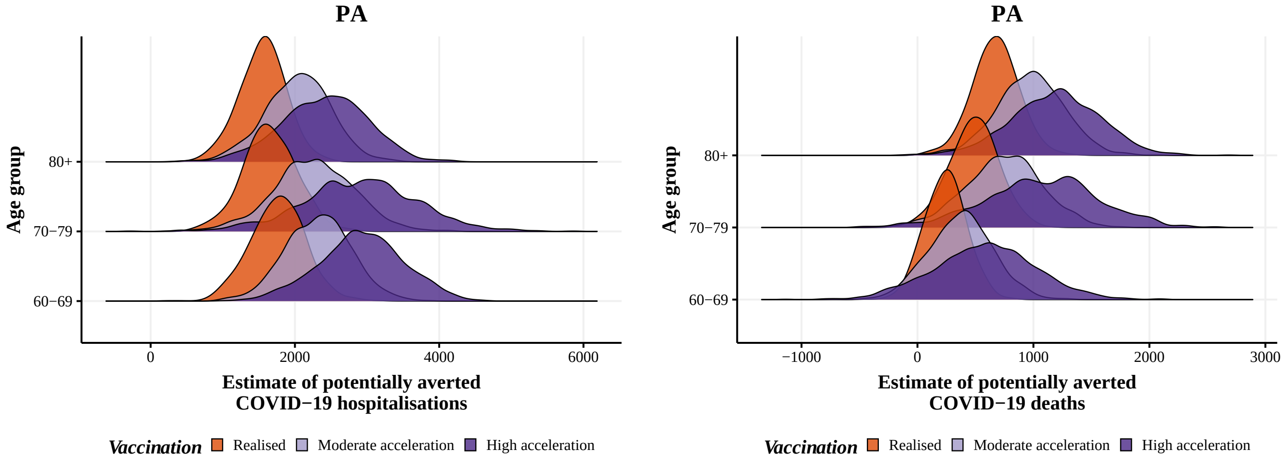


**Figure S15:** Posterior distribution of hospitalisations (left) and deaths (right) potentially averted by vaccination between 2021-01-01 and 2021-08-29 by age group, with the realized (orange), 4 (light purple) and 8 weeks earlier (dark purple) vaccination rollout in PA state.

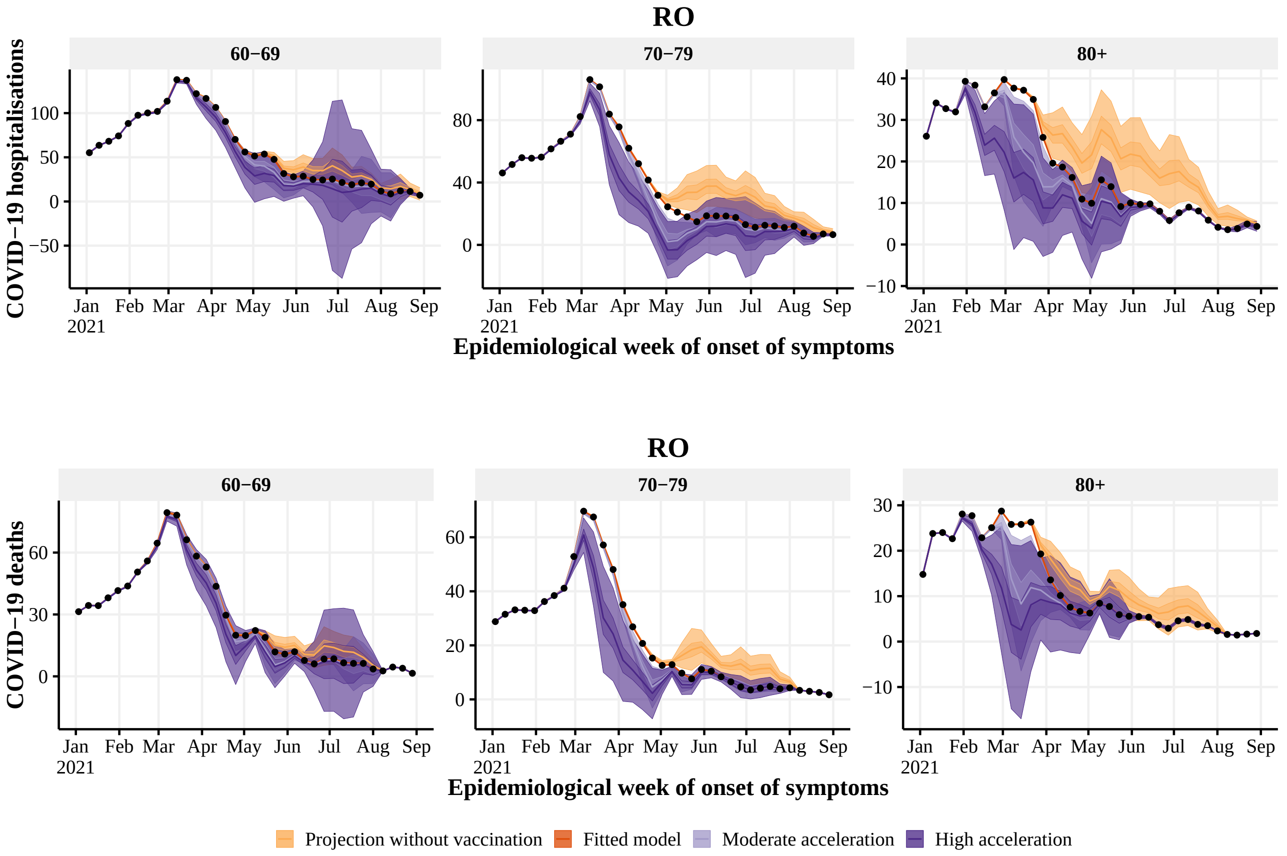


**Figure S16:** Estimated number of hospitalisations (top) and deaths (bottom) by epidemiological week with the realized (dark orange), no vaccination (light orange), 4 (light purple) and 8 (dark purple) weeks earlier vaccination rollout, by age group (panels), in RO state. The observed number of hospitalisations and deaths are given by the black points. With 50% and 95% Credible Intervals.

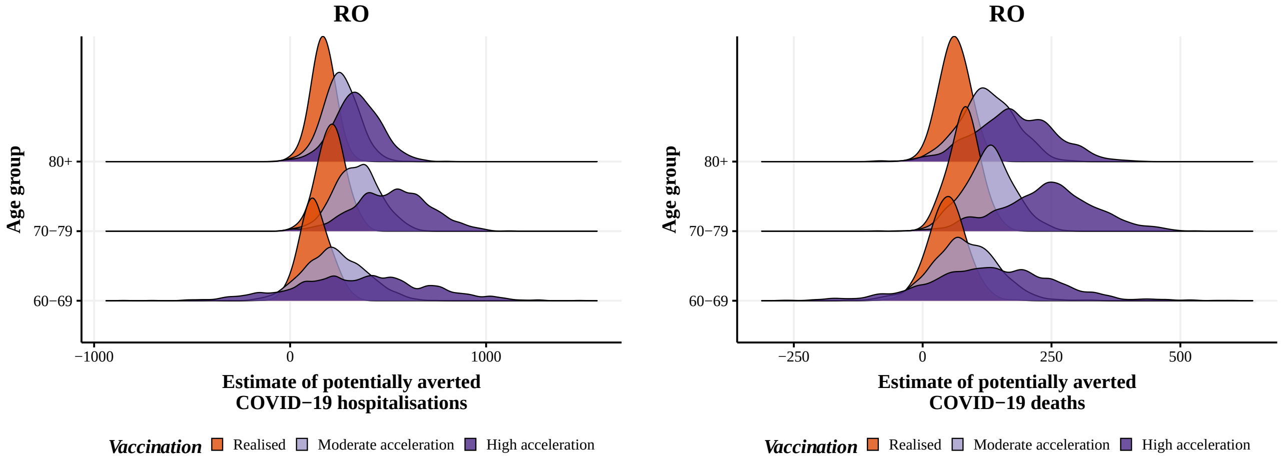


**Figure S17:** Posterior distribution of hospitalisations (left) and deaths (right) potentially averted by vaccination between 2021-01-01 and 2021-08-29 by age group, with the realized (orange), 4 (light purple) and 8 weeks earlier (dark purple) vaccination rollout in RO state.

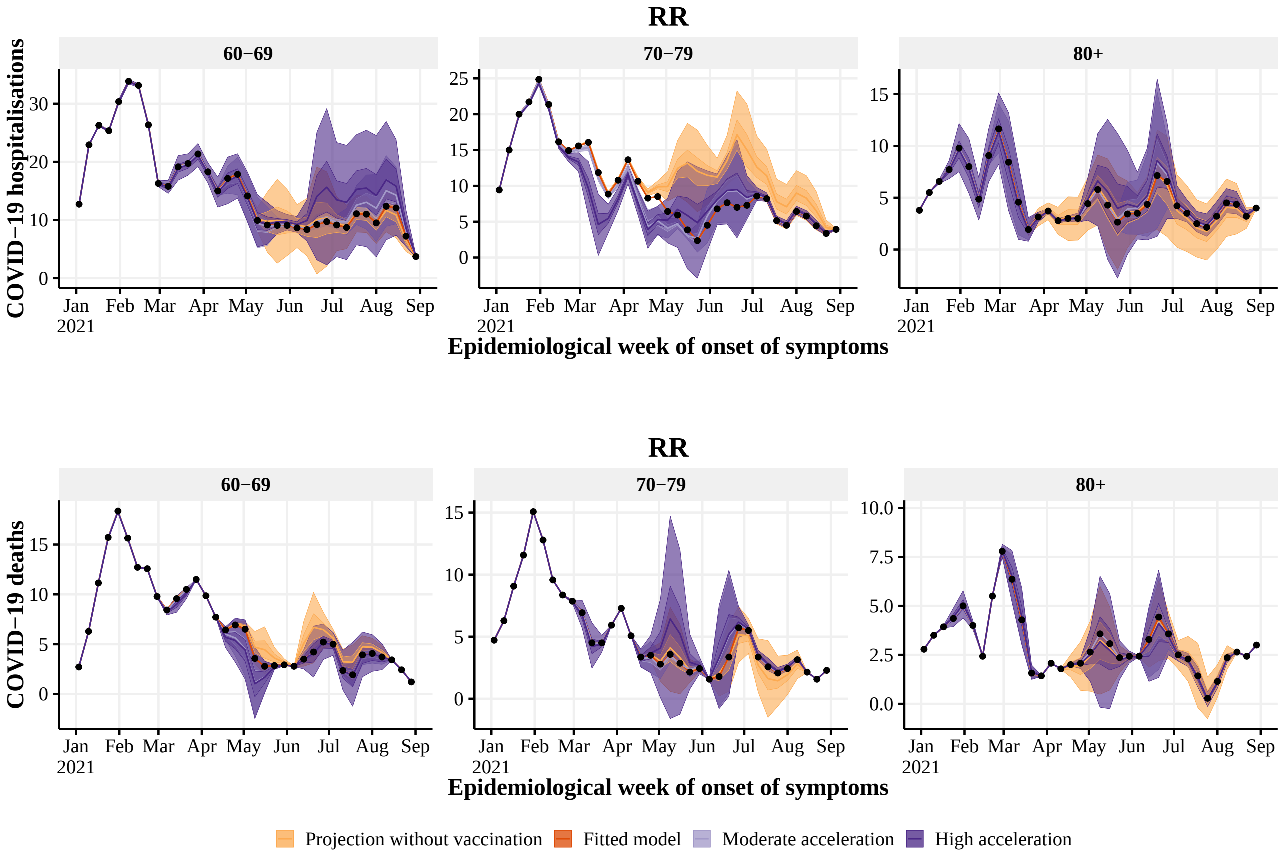


**Figure S18:** Estimated number of hospitalisations (top) and deaths (bottom) by epidemiological week with the realized (dark orange), no vaccination (light orange), 4 (light purple) and 8 (dark purple) weeks earlier vaccination rollout, by age group (panels), in RR state. The observed number of hospitalisations and deaths are given by the black points. With 50% and 95% Credible Intervals.

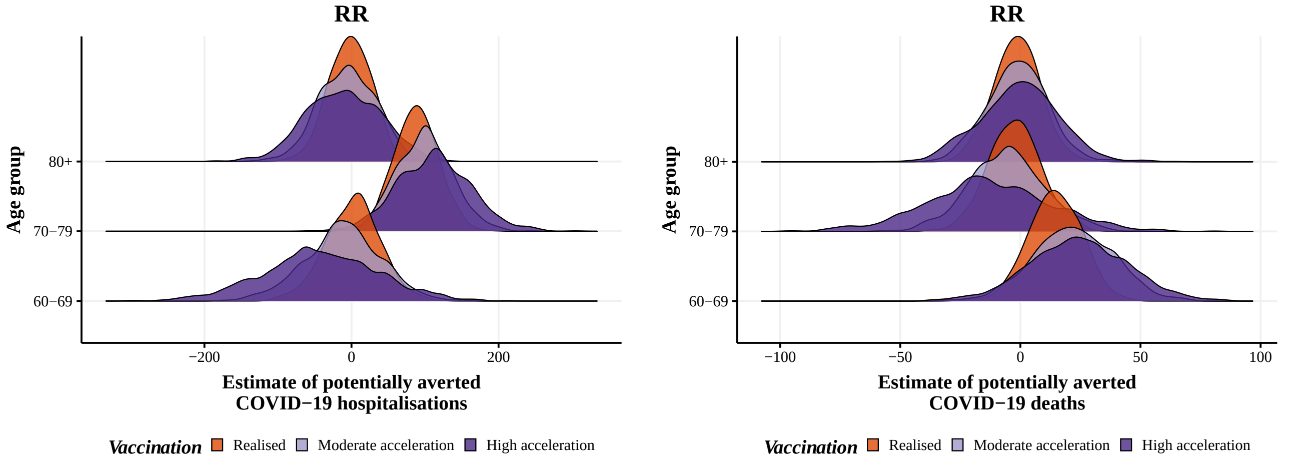


**Figure S19:** Posterior distribution of hospitalisations (left) and deaths (right) potentially averted by vaccination between 2021-01-01 and 2021-08-29 by age group, with the realized (orange), 4 (light purple) and 8 weeks earlier (dark purple) vaccination rollout in RR state.

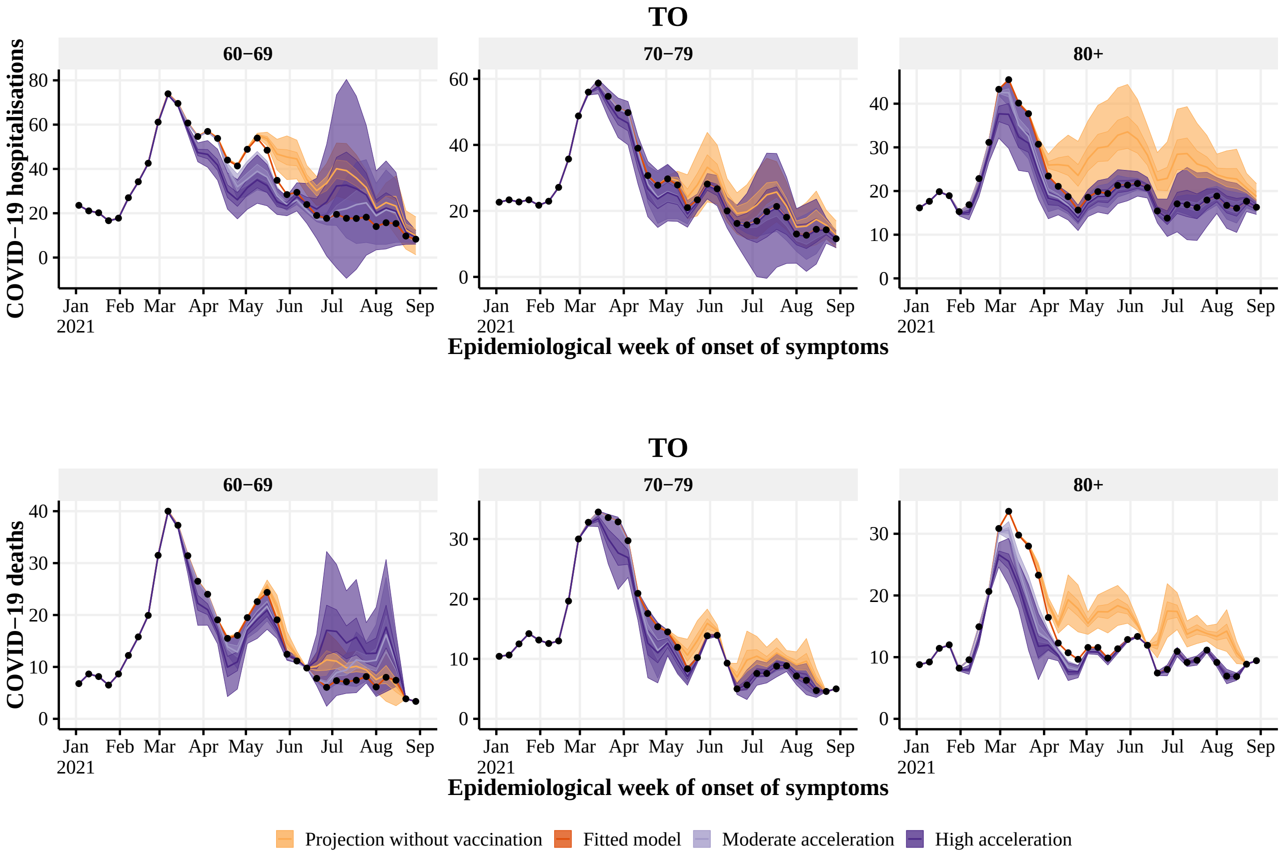


**Figure S20:** Estimated number of hospitalisations (top) and deaths (bottom) by epidemiological week with the realized (dark orange), no vaccination (light orange), 4 (light purple) and 8 (dark purple) weeks earlier vaccination rollout, by age group (panels), in TO state. The observed number of hospitalisations and deaths are given by the black points. With 50% and 95% Credible Intervals.

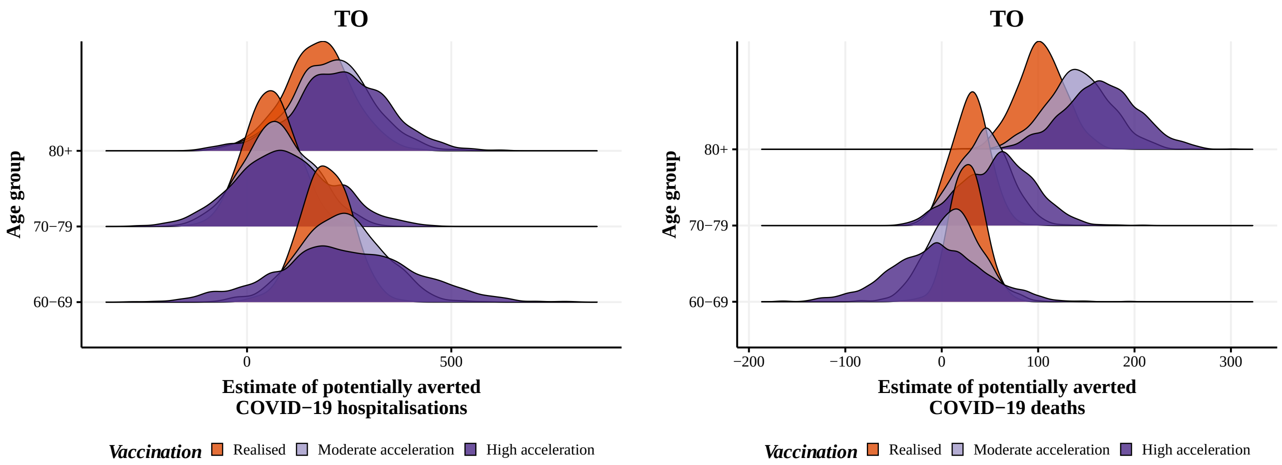


**Figure S21:** Posterior distribution of hospitalisations (left) and deaths (right) potentially averted by vaccination between 2021-01-01 and 2021-08-29 by age group, with the realized (orange), 4 (light purple) and 8 weeks earlier (dark purple) vaccination rollout in TO state.

#### 5.4 Northeast Region

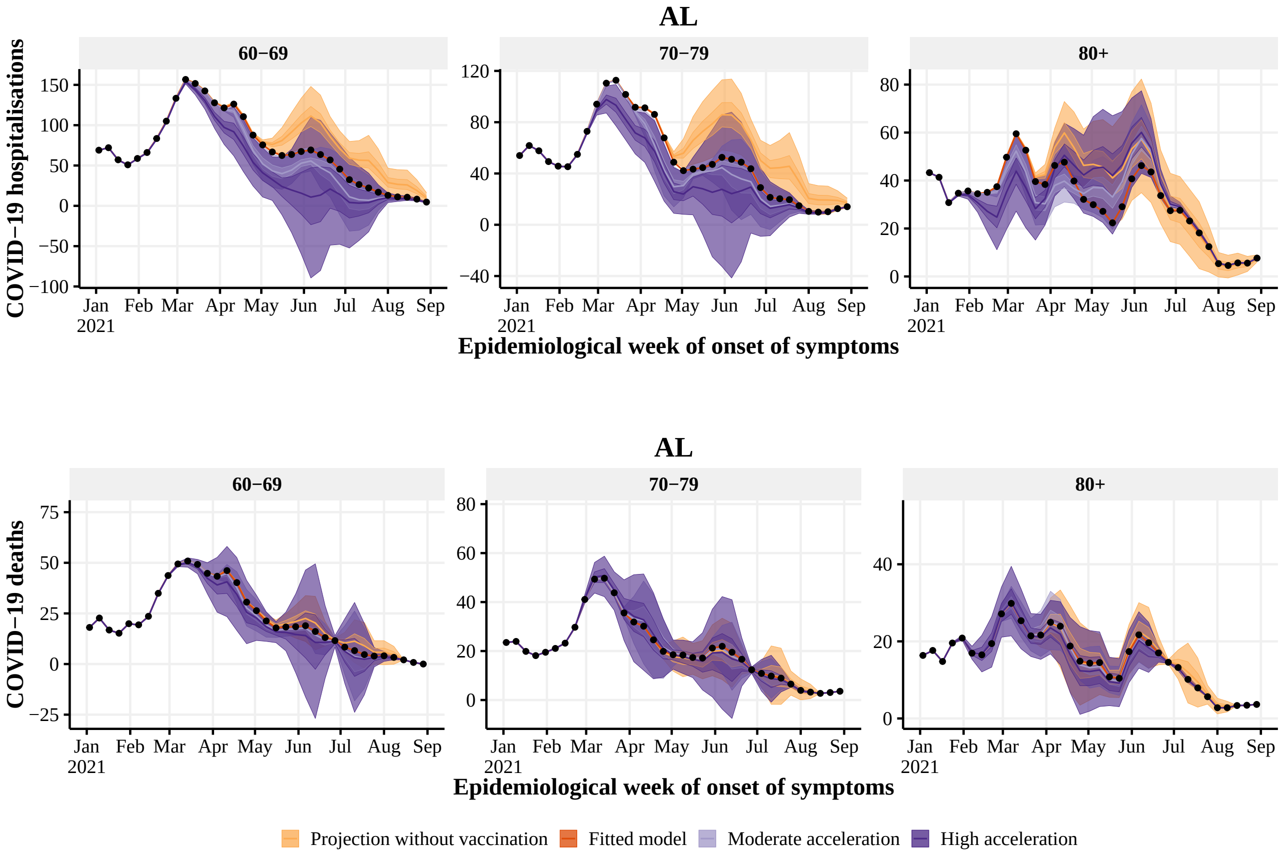


**Figure S22:** Estimated number of hospitalisations (top) and deaths (bottom) by epidemiological week with the realized (dark orange), no vaccination (light orange), 4 (light purple) and 8 (dark purple) weeks earlier vaccination rollout, by age group (panels), in AL state. The observed number of hospitalisations and deaths are given by the black points. With 50% and 95% Credible Intervals.

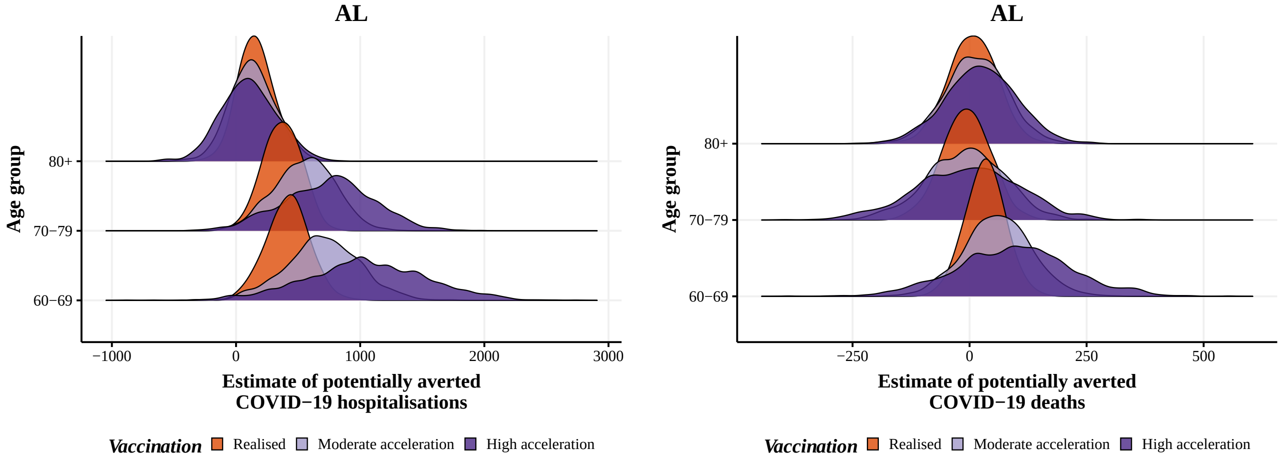


**Figure S23:** Posterior distribution of hospitalisations (left) and deaths (right) potentially averted by vaccination between 2021-01-01 and 2021-08-29 by age group, with the realized (orange), 4 (blue) and 8 weeks earlier (green) vaccination rollout in AL state.

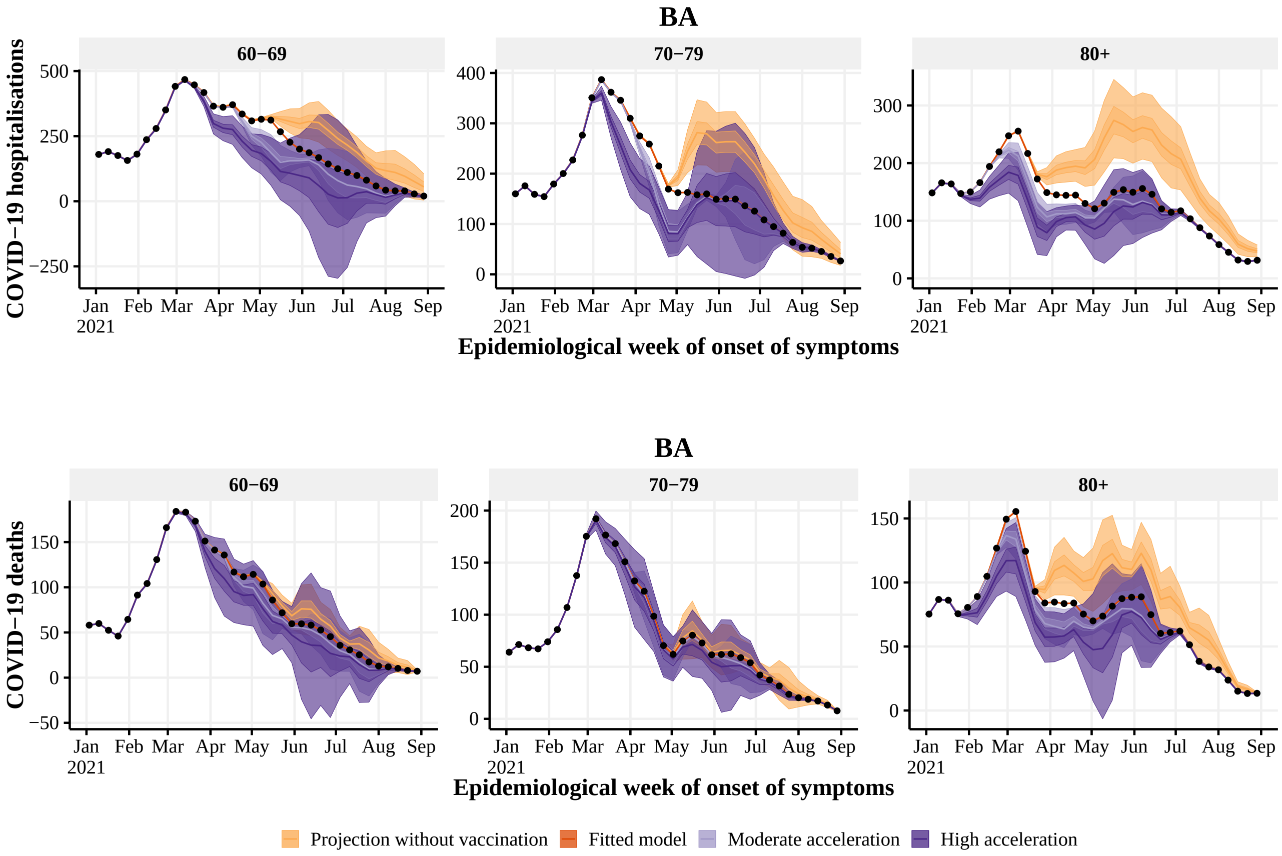


**Figure S24:** Estimated number of hospitalisations (top) and deaths (bottom) by epidemiological week with the realized (dark orange), no vaccination (light orange), 4 (light purple) and 8 (dark purple) weeks earlier vaccination rollout, by age group (panels), in BA state. The observed number of hospitalisations and deaths are given by the black points. With 50% and 95% Credible Intervals.

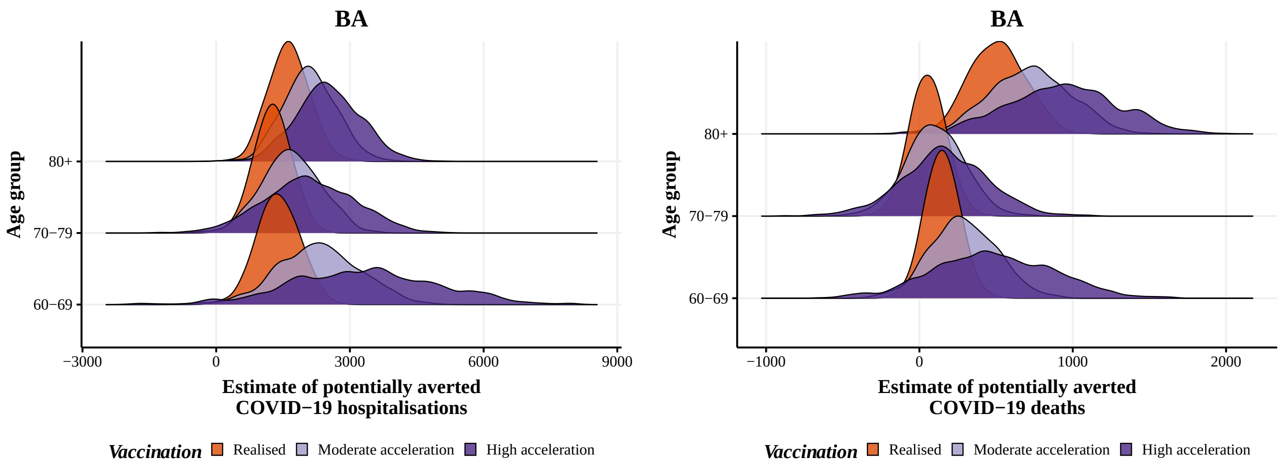


**Figure S25:** Posterior distribution of hospitalisations (left) and deaths (right) potentially averted by vaccination between 2021-01-01 and 2021-08-29 by age group, with the realized (orange), 4 (blue) and 8 weeks earlier (green) vaccination rollout in BA state.

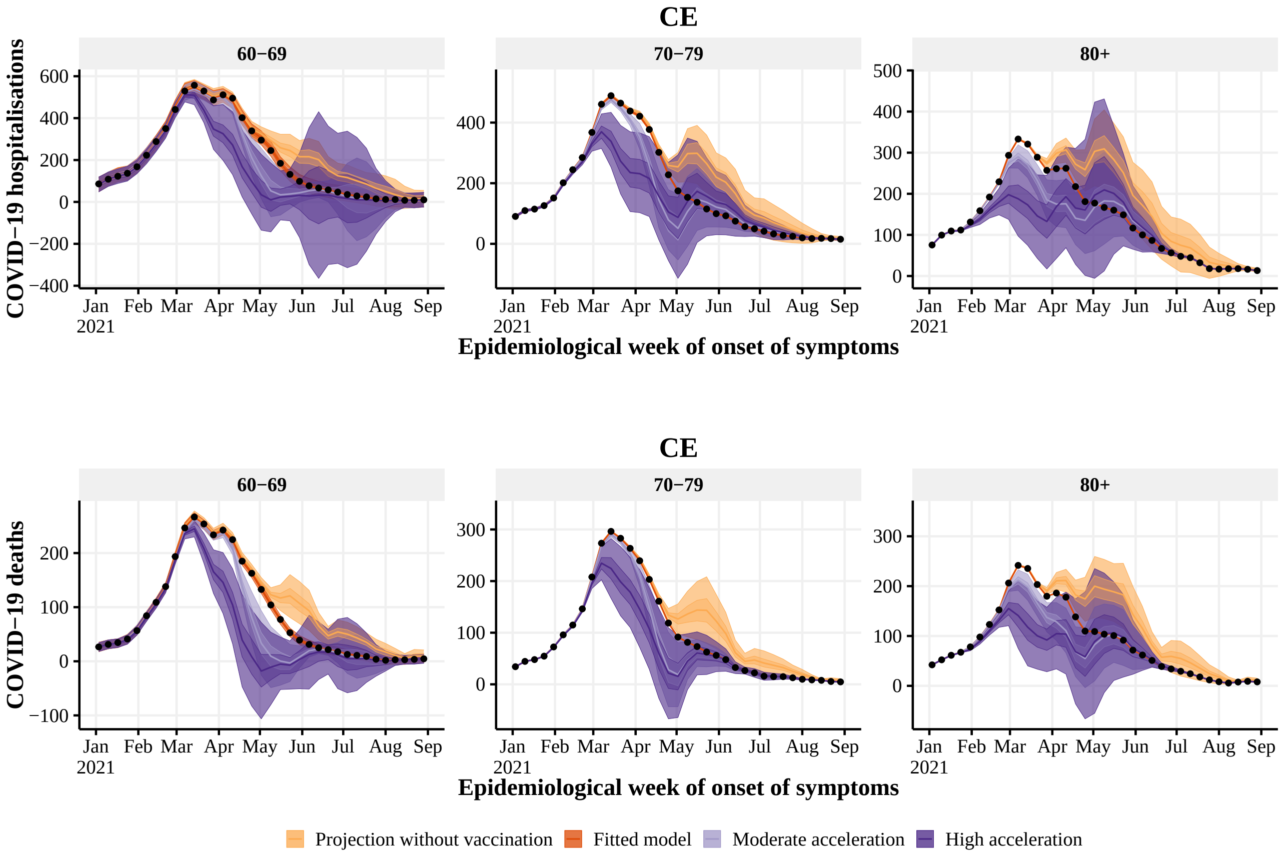


**Figure S26:** Estimated number of hospitalisations (top) and deaths (bottom) by epidemiological week with the realized (dark orange), no vaccination (light orange), 4 (light purple) and 8 (dark purple) weeks earlier vaccination rollout, by age group (panels), in CE state. The observed number of hospitalisations and deaths are given by the black points. With 50% and 95% Credible Intervals.

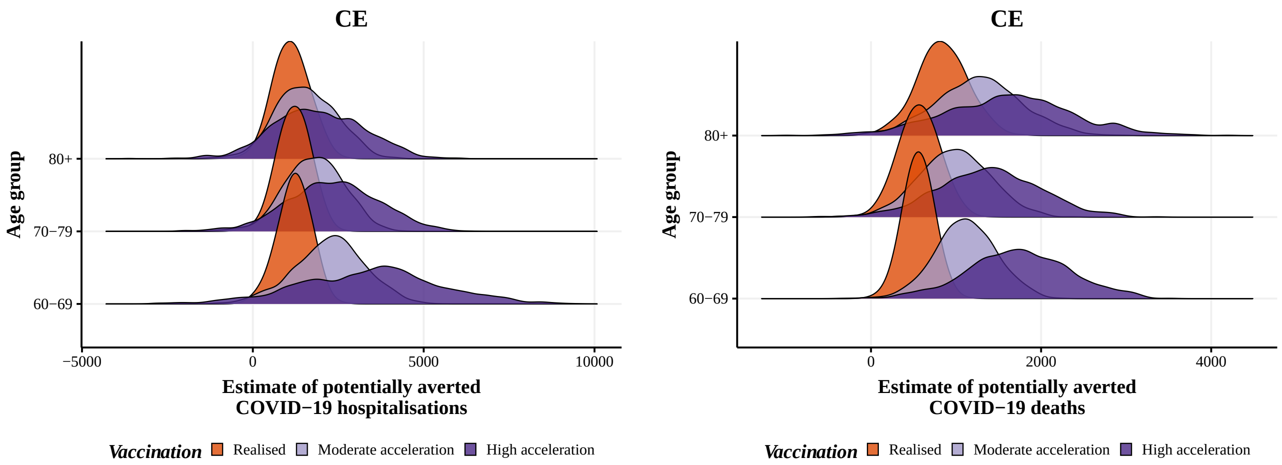


**Figure S27:** Posterior distribution of hospitalisations (left) and deaths (right) potentially averted by vaccination between 2021-01-01 and 2021-08-29 by age group, with the realized (orange), 4 (blue) and 8 weeks earlier (green) vaccination rollout in CE state.

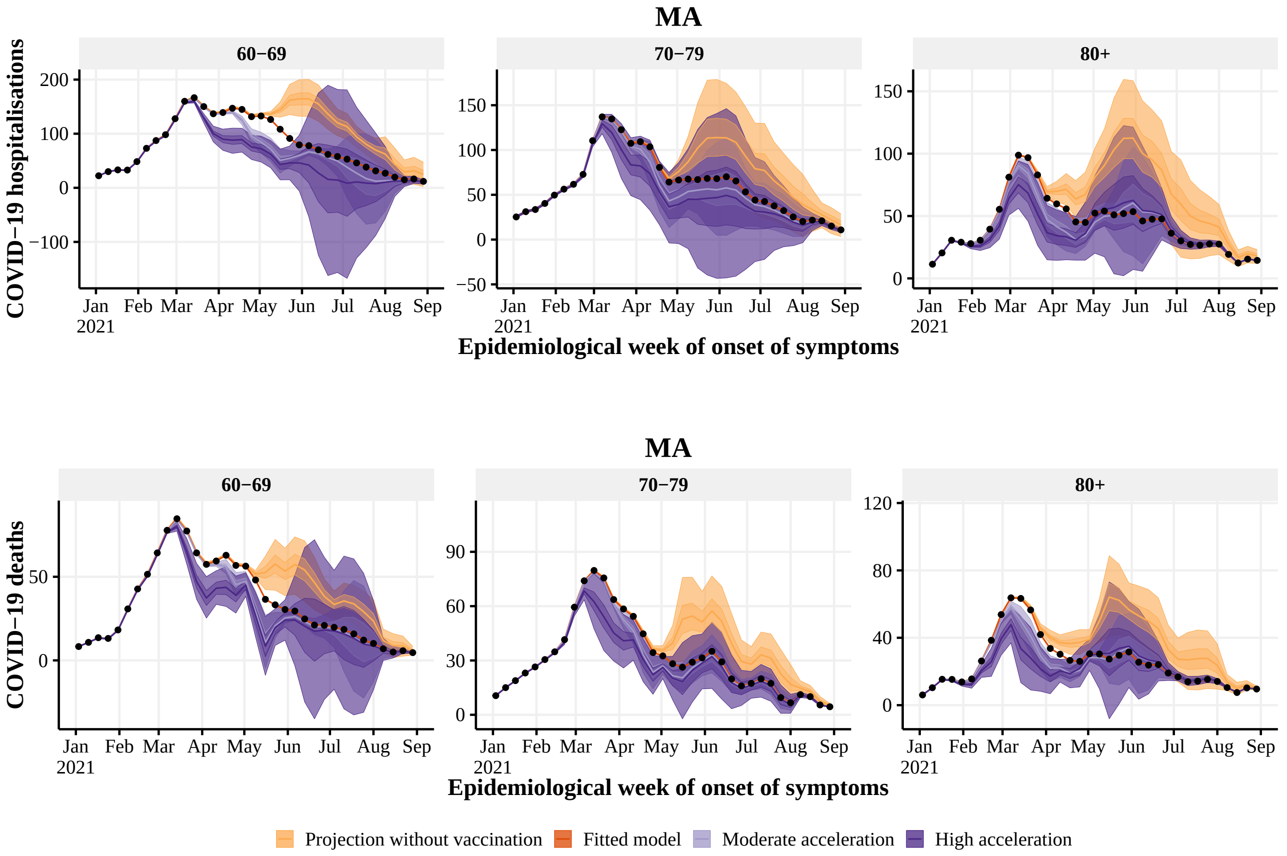


**Figure S28:** Estimated number of hospitalisations (top) and deaths (bottom) by epidemiological week with the realized (dark orange), no vaccination (light orange), 4 (light purple) and 8 (dark purple) weeks earlier vaccination rollout, by age group (panels), in MA state. The observed number of hospitalisations and deaths are given by the black points. With 50% and 95% Credible Intervals.

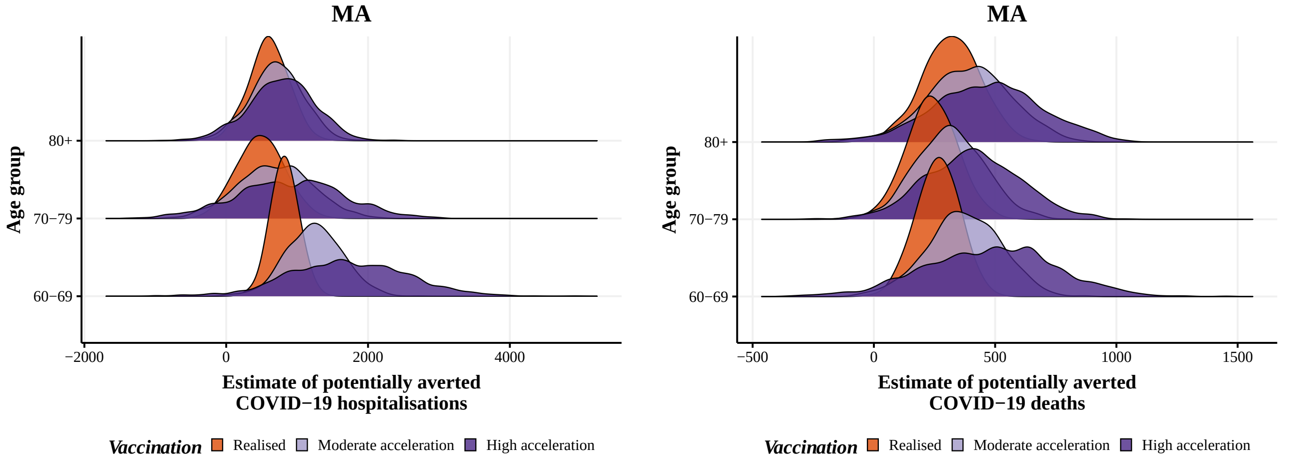


**Figure S29:** Posterior distribution of hospitalisations (left) and deaths (right) potentially averted by vaccination between 2021-01-01 and 2021-08-29 by age group, with the realized (orange), 4 (blue) and 8 weeks earlier (green) vaccination rollout in MA state.

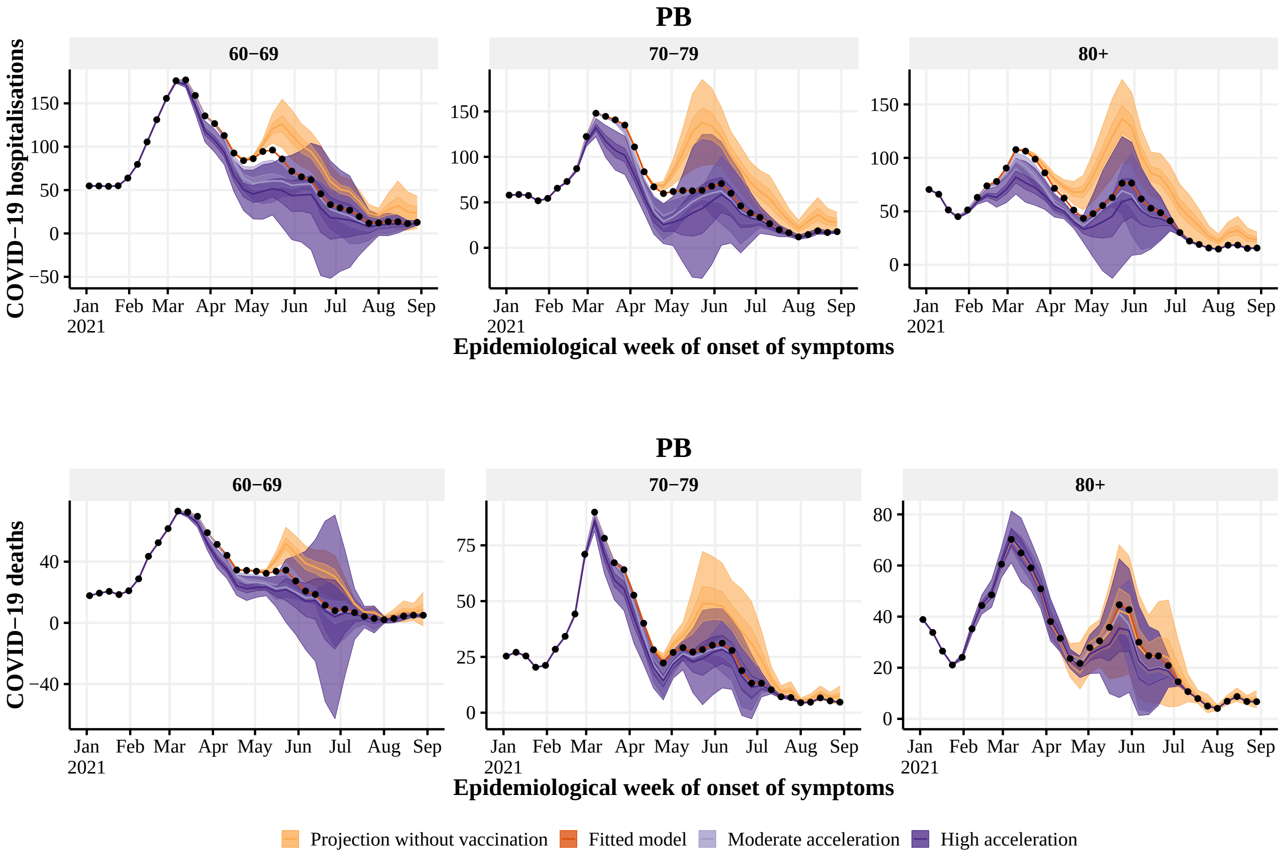


**Figure S30:** Estimated number of hospitalisations (top) and deaths (bottom) by epidemiological week with the realized (dark orange), no vaccination (light orange), 4 (light purple) and 8 (dark purple) weeks earlier vaccination rollout, by age group (panels), in PB state. The observed number of hospitalisations and deaths are given by the black points. With 50% and 95% Credible Intervals.

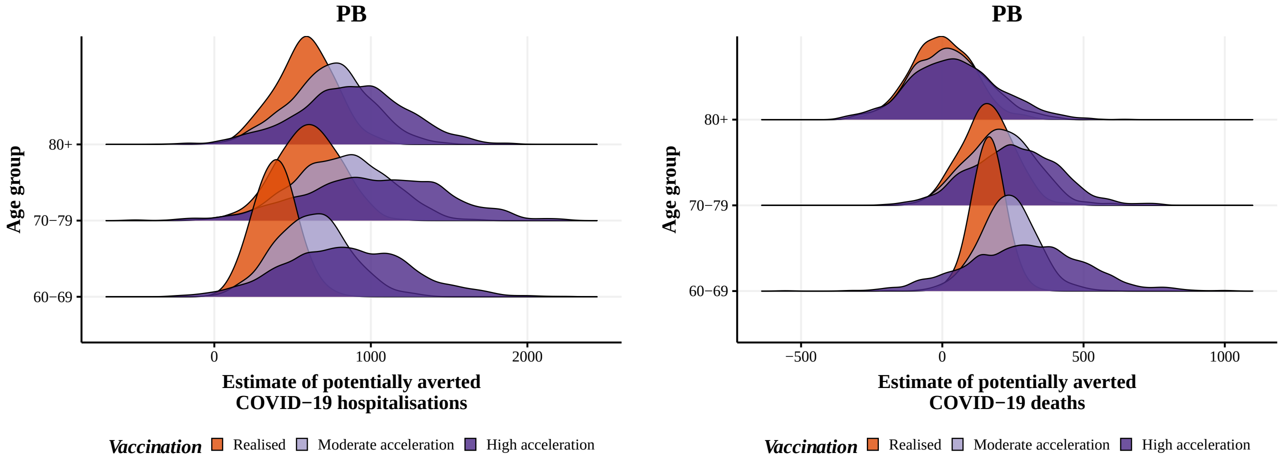


**Figure S31:** Posterior distribution of hospitalisations (left) and deaths (right) potentially averted by vaccination between 2021-01-01 and 2021-08-29 by age group, with the realized (orange), 4 (blue) and 8 weeks earlier (green) vaccination rollout in PB state.

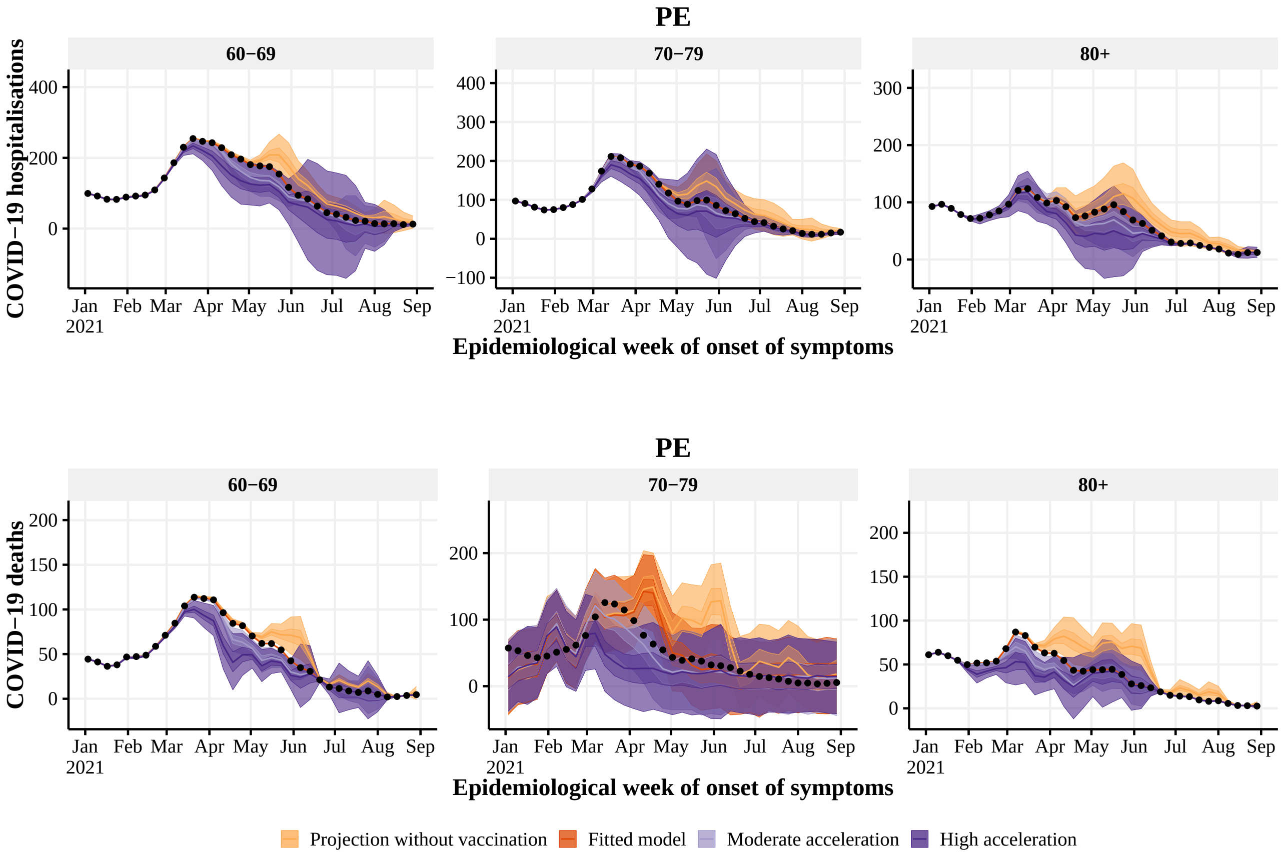


**Figure S32:** Estimated number of hospitalisations (top) and deaths (bottom) by epidemiological week with the realized (dark orange), no vaccination (light orange), 4 (light purple) and 8 (dark purple) weeks earlier vaccination rollout, by age group (panels), in PE state. The observed number of hospitalisations and deaths are given by the black points. With 50% and 95% Credible Intervals.

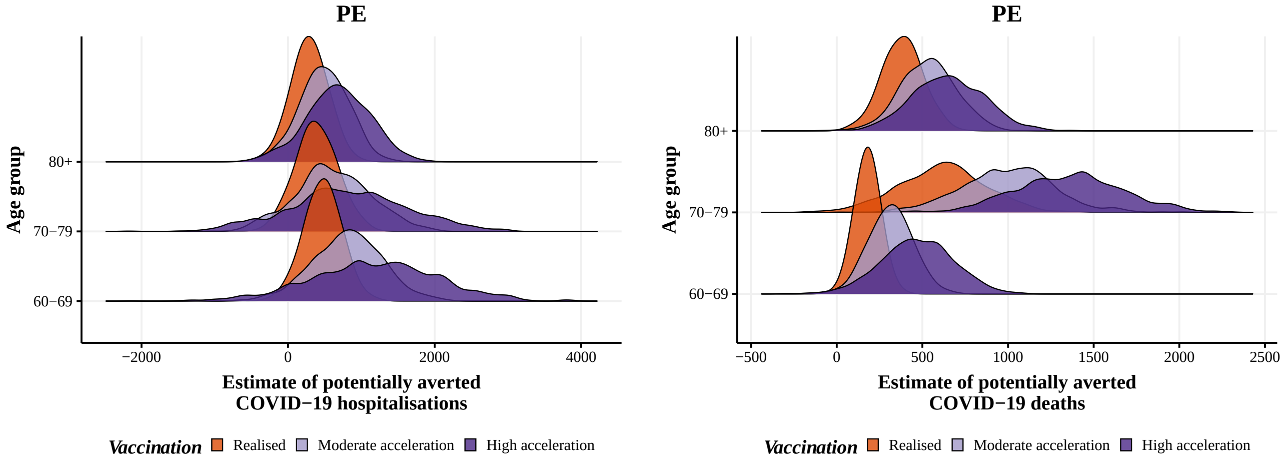


**Figure S33:** Posterior distribution of hospitalisations (left) and deaths (right) potentially averted by vaccination between 2021-01-01 and 2021-08-29 by age group, with the realized (orange), 4 (blue) and 8 weeks earlier (green) vaccination rollout in PE state.

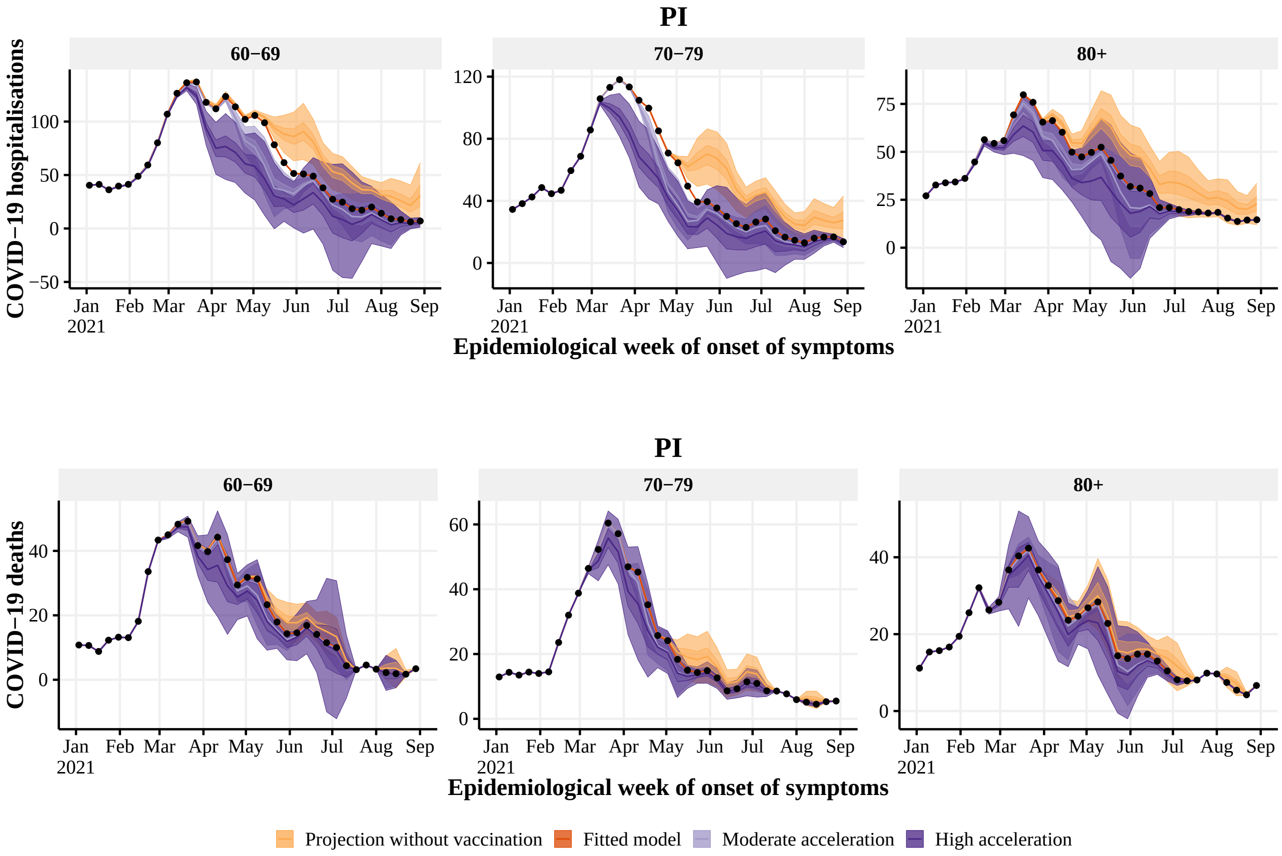


**Figure S34:** Estimated number of hospitalisations (top) and deaths (bottom) by epidemiological week with the realized (dark orange), no vaccination (light orange), 4 (light purple) and 8 (dark purple) weeks earlier vaccination rollout, by age group (panels), in PI state. The observed number of hospitalisations and deaths are given by the black points. With 50% and 95% Credible Intervals.

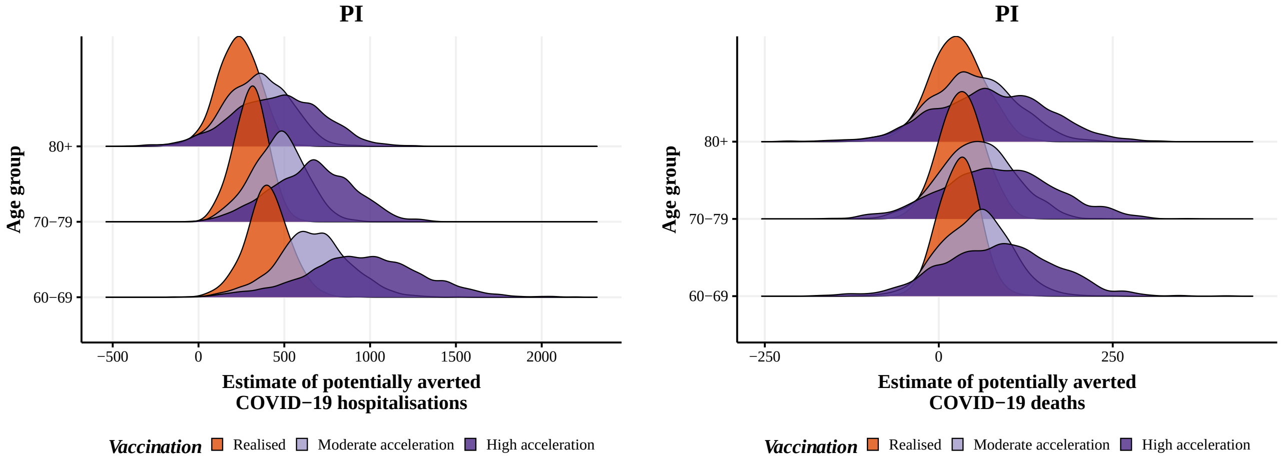


**Figure S35:** Posterior distribution of hospitalisations (left) and deaths (right) potentially averted by vaccination between 2021-01-01 and 2021-08-29 by age group, with the realized (orange), 4 (blue) and 8 weeks earlier (green) vaccination rollout in PI state.

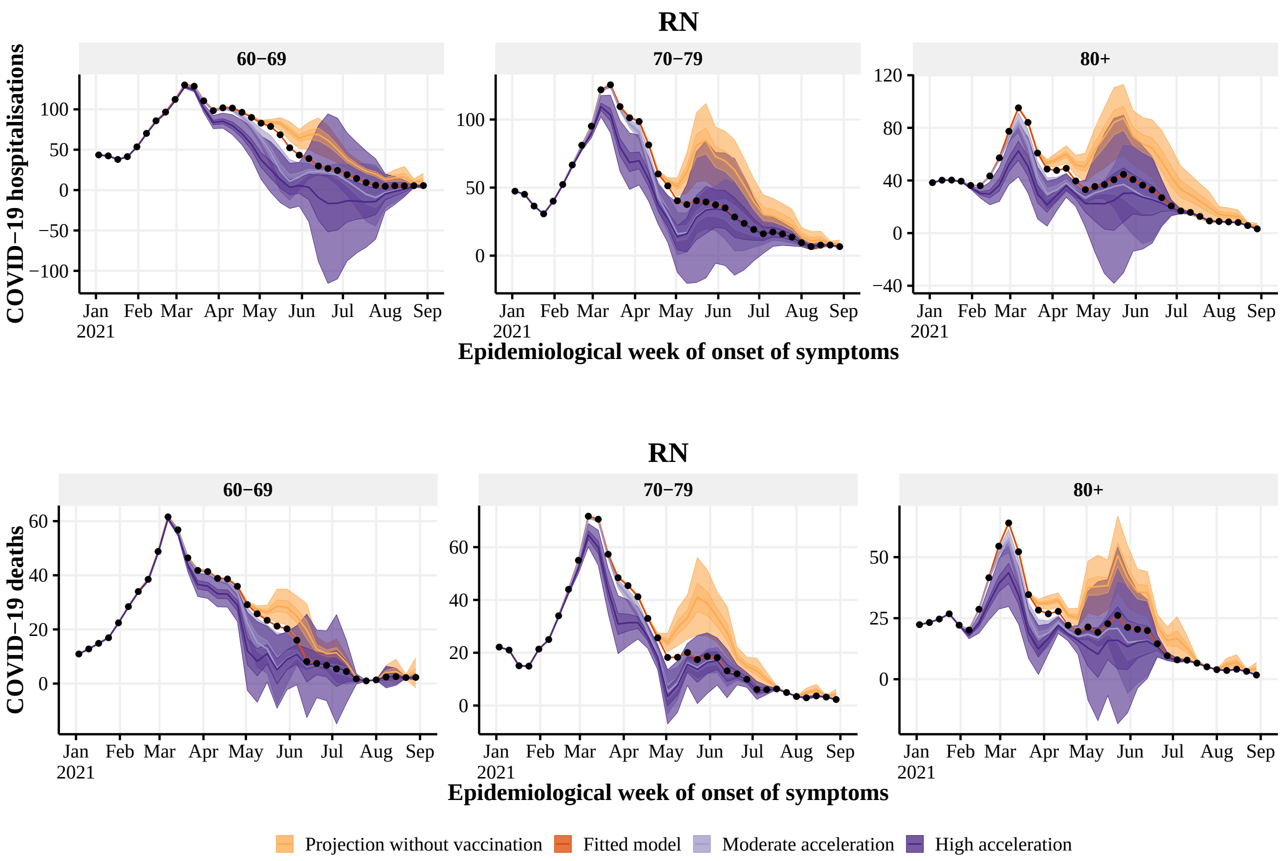


**Figure S36:** Estimated number of hospitalisations (top) and deaths (bottom) by epidemiological week with the realized (dark orange), no vaccination (light orange), 4 (light purple) and 8 (dark purple) weeks earlier vaccination rollout, by age group (panels), in RN state. The observed number of hospitalisations and deaths are given by the black points. With 50% and 95% Credible Intervals.

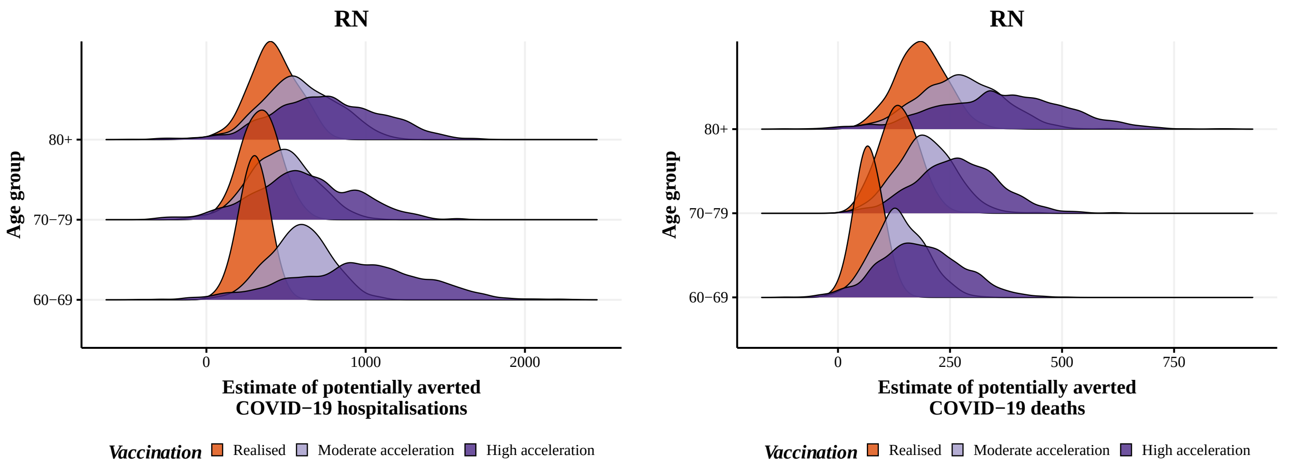


**Figure S37:** Posterior distribution of hospitalisations (left) and deaths (right) potentially averted by vaccination between 2021-01-01 and 2021-08-29 by age group, with the realized (orange), 4 (blue) and 8 weeks earlier (green) vaccination rollout in RN state.

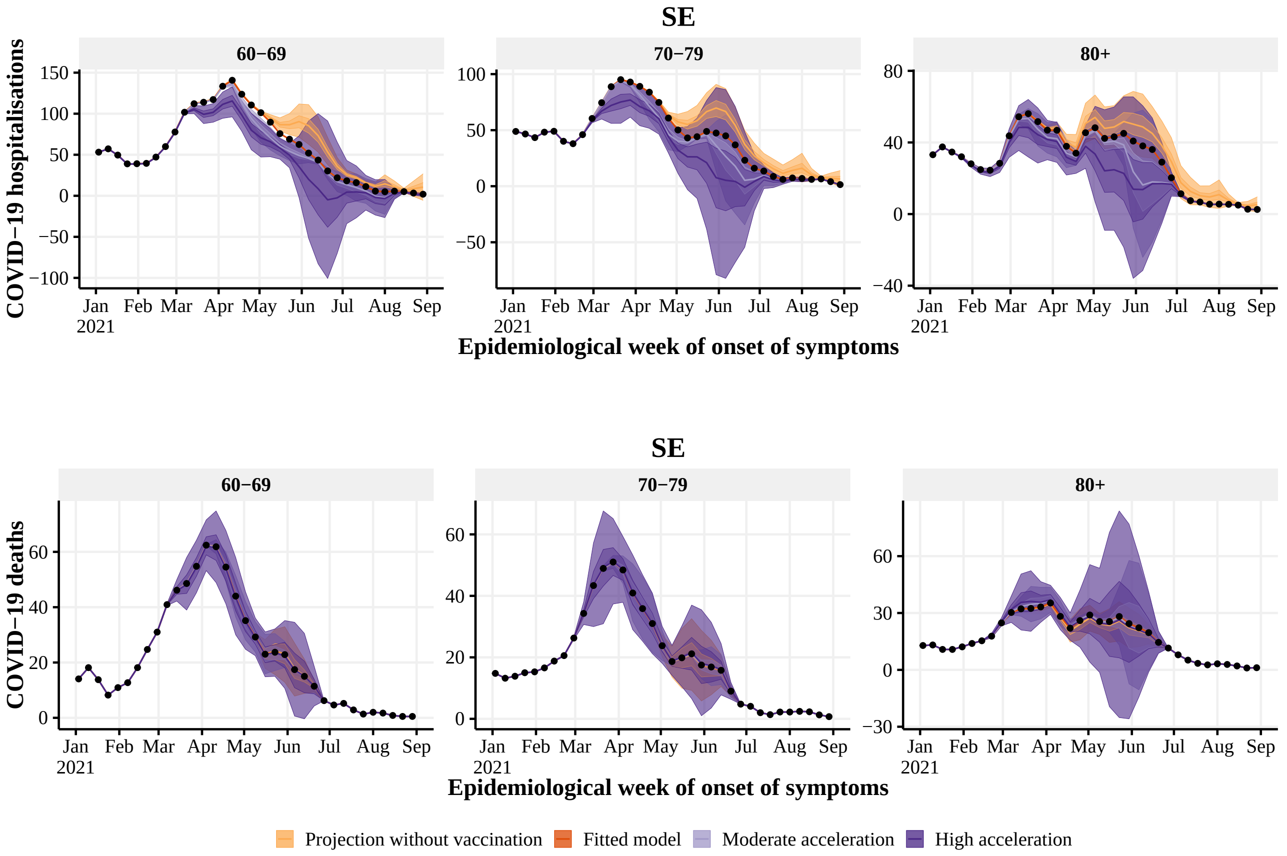


**Figure S38:** Estimated number of hospitalisations (top) and deaths (bottom) by epidemiological week with the realized (dark orange), no vaccination (light orange), 4 (light purple) and 8 (dark purple) weeks earlier vaccination rollout, by age group (panels), in SE state. The observed number of hospitalisations and deaths are given by the black points. With 50% and 95% Credible Intervals.

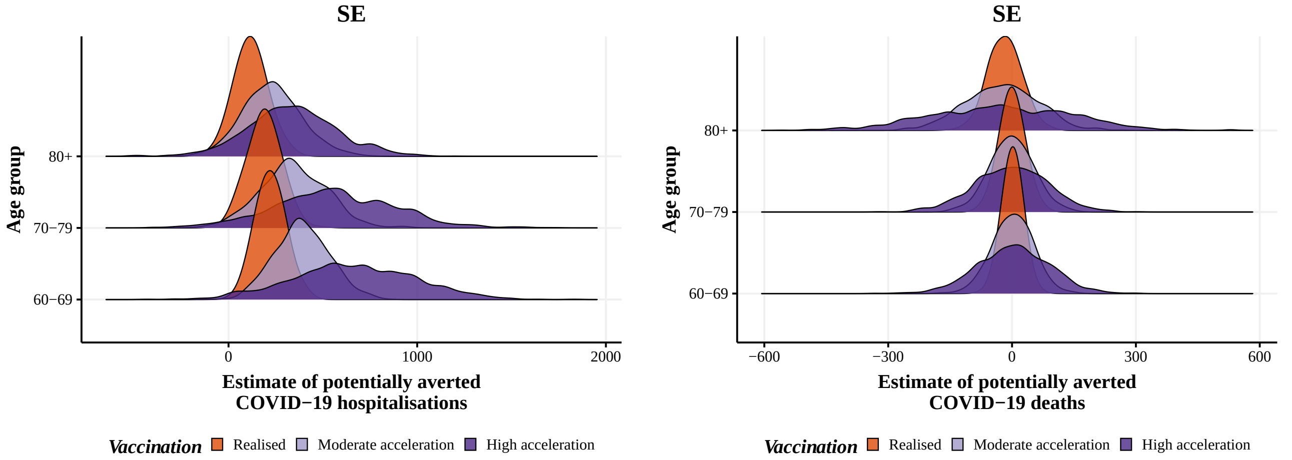


**Figure S39:** Posterior distribution of hospitalisations (left) and deaths (right) potentially averted by vaccination between 2021-01-01 and 2021-08-29 by age group, with the realized (orange), 4 (blue) and 8 weeks earlier (green) vaccination rollout in SE state.

#### 5.5 Southeast Region

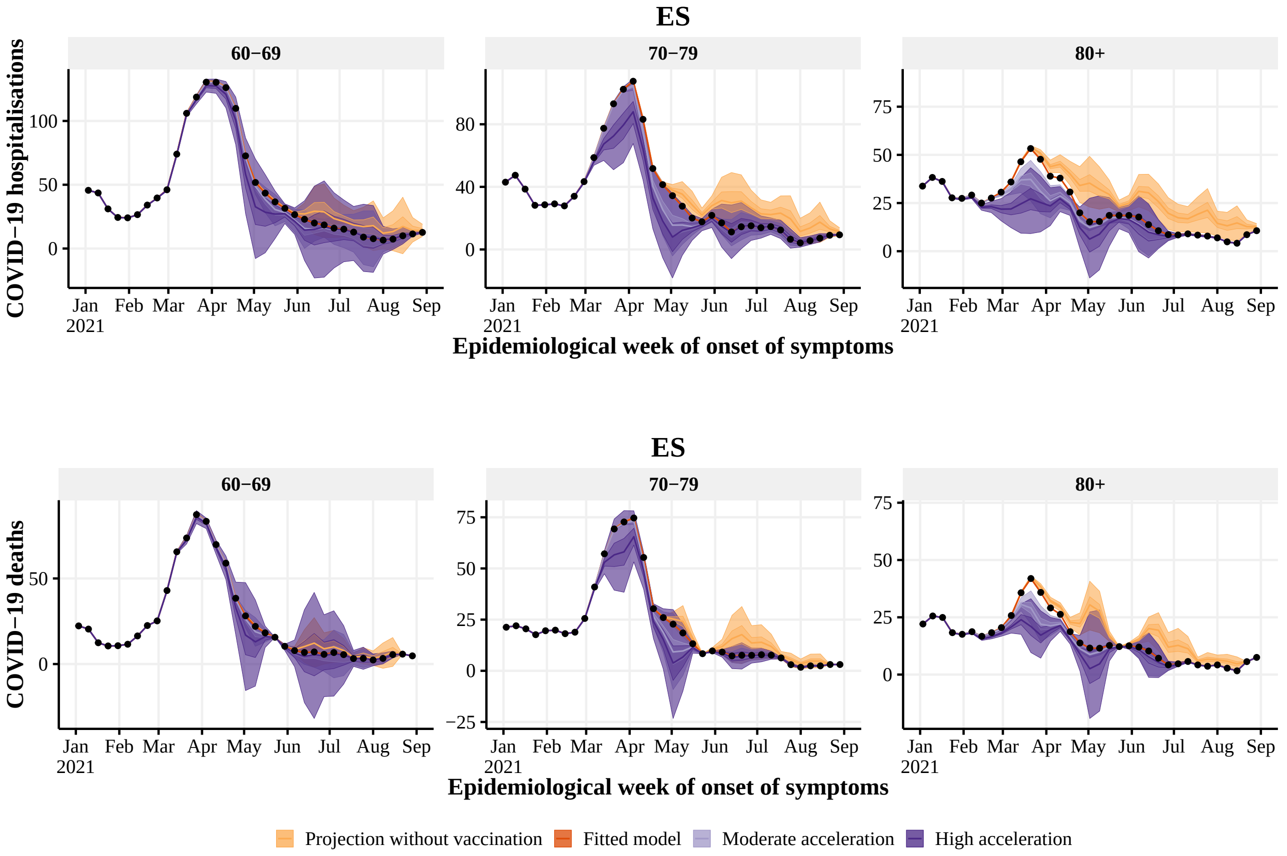


**Figure S40:** Estimated number of hospitalisations (top) and deaths (bottom) by epidemiological week with the realized (dark orange), no vaccination (light orange), 4 (light purple) and 8 (dark purple) weeks earlier vaccination rollout, by age group (panels), in ES state. The observed number of hospitalisations and deaths are given by the black points. With 50% and 95% Credible Intervals.

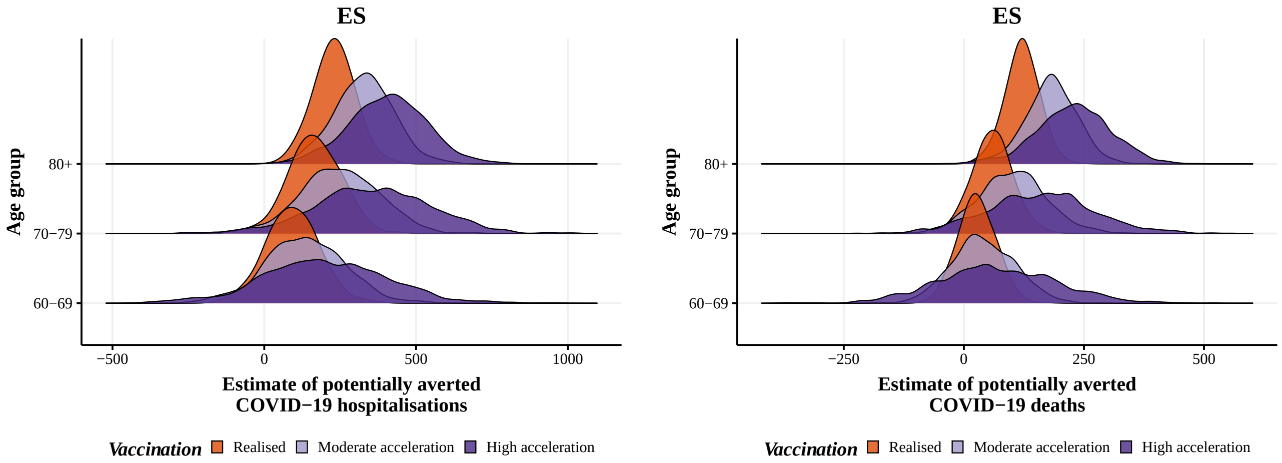


**Figure S41:** Posterior distribution of hospitalisations (left) and deaths (right) potentially averted by vaccination between 2021-01-01 and 2021-08-29 by age group, with the realized (orange), 4 (blue) and 8 weeks earlier (green) vaccination rollout in ES state.

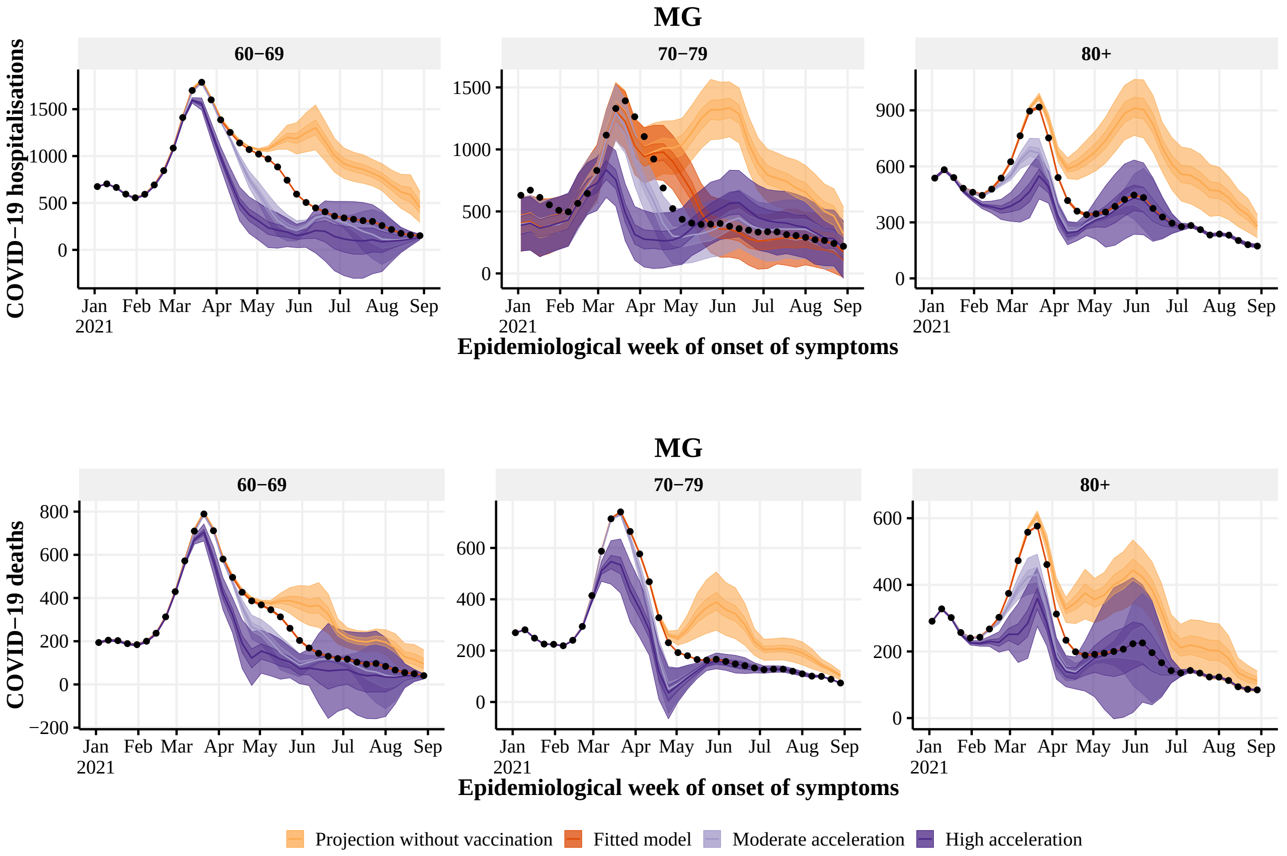


**Figure S42:** Estimated number of hospitalisations (top) and deaths (bottom) by epidemiological week with the realized (dark orange), no vaccination (light orange), 4 (light purple) and 8 (dark purple) weeks earlier vaccination rollout, by age group (panels), in MG state. The observed number of hospitalisations and deaths are given by the black points. With 50% and 95% Credible Intervals.

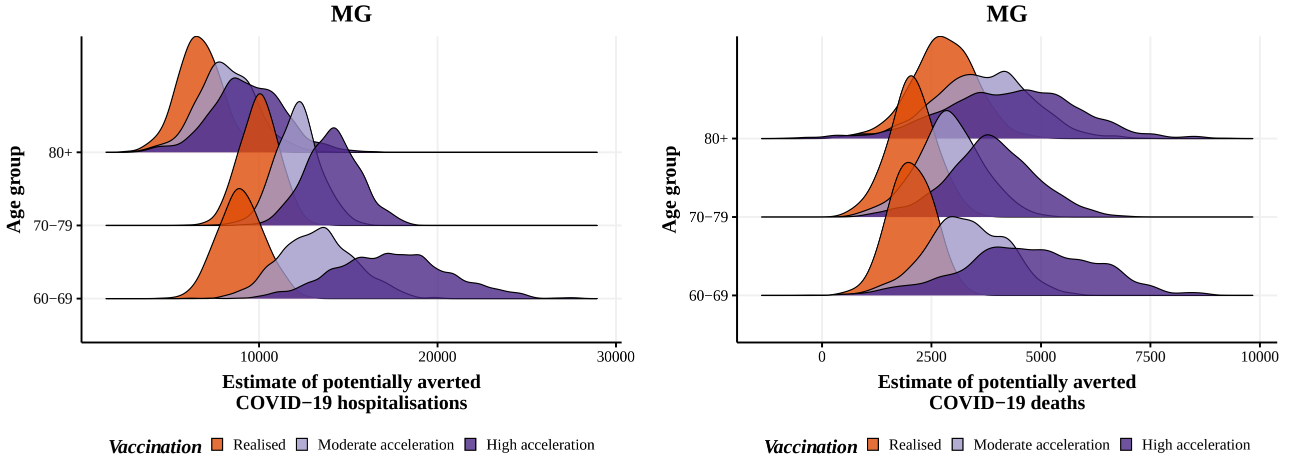


**Figure S43:** Posterior distribution of hospitalisations (left) and deaths (right) potentially averted by vaccination between 2021-01-01 and 2021-08-29 by age group, with the realized (orange), 4 (blue) and 8 weeks earlier (green) vaccination rollout in MG state.

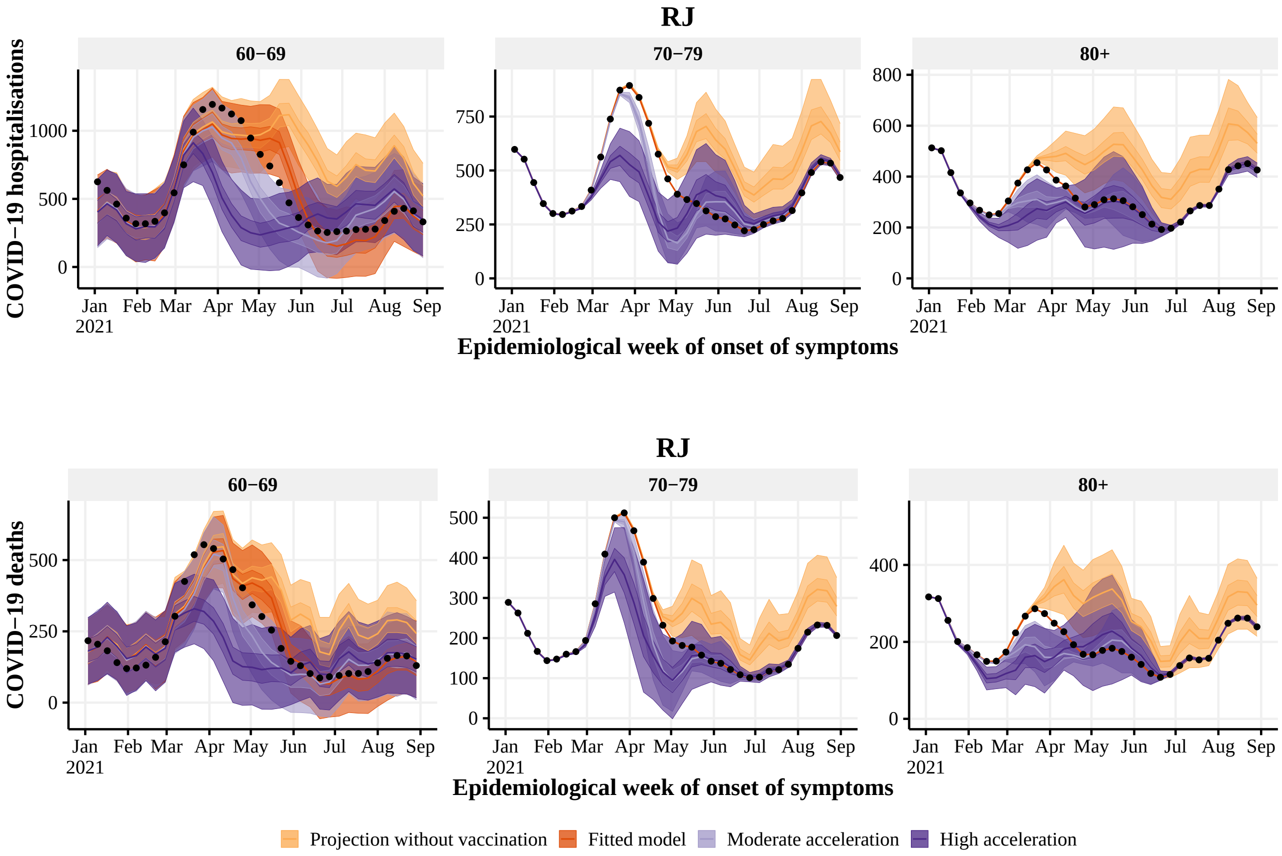


**Figure S44:** Estimated number of hospitalisations (top) and deaths (bottom) by epidemiological week with the realized (dark orange), no vaccination (light orange), 4 (light purple) and 8 (dark purple) weeks earlier vaccination rollout, by age group (panels), in RJ state. The observed number of hospitalisations and deaths are given by the black points. With 50% and 95% Credible Intervals.

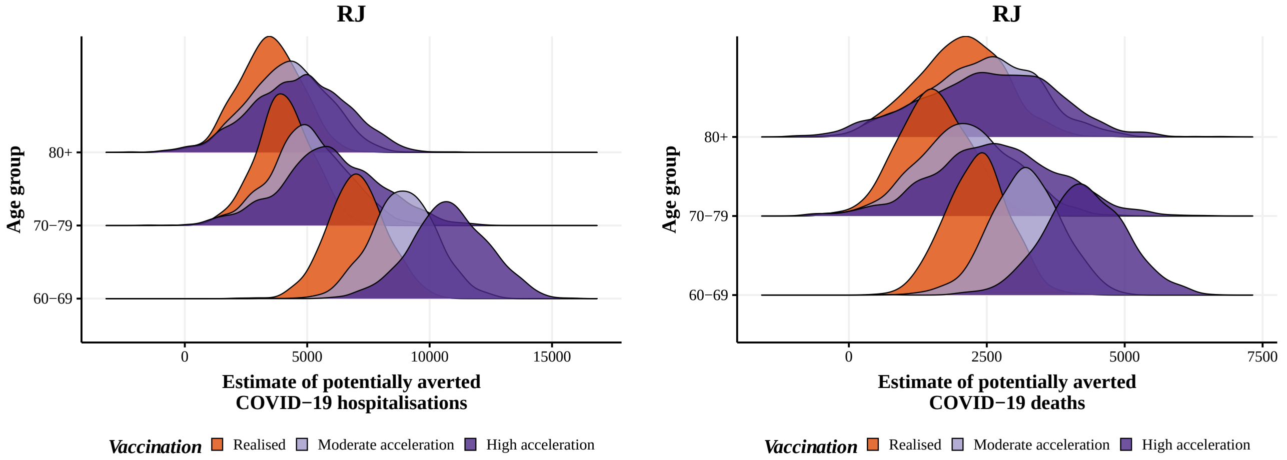


**Figure S45:** Posterior distribution of hospitalisations (left) and deaths (right) potentially averted by vaccination between 2021-01-01 and 2021-08-29 by age group, with the realized (orange), 4 (blue) and 8 weeks earlier (green) vaccination rollout in RJ state.

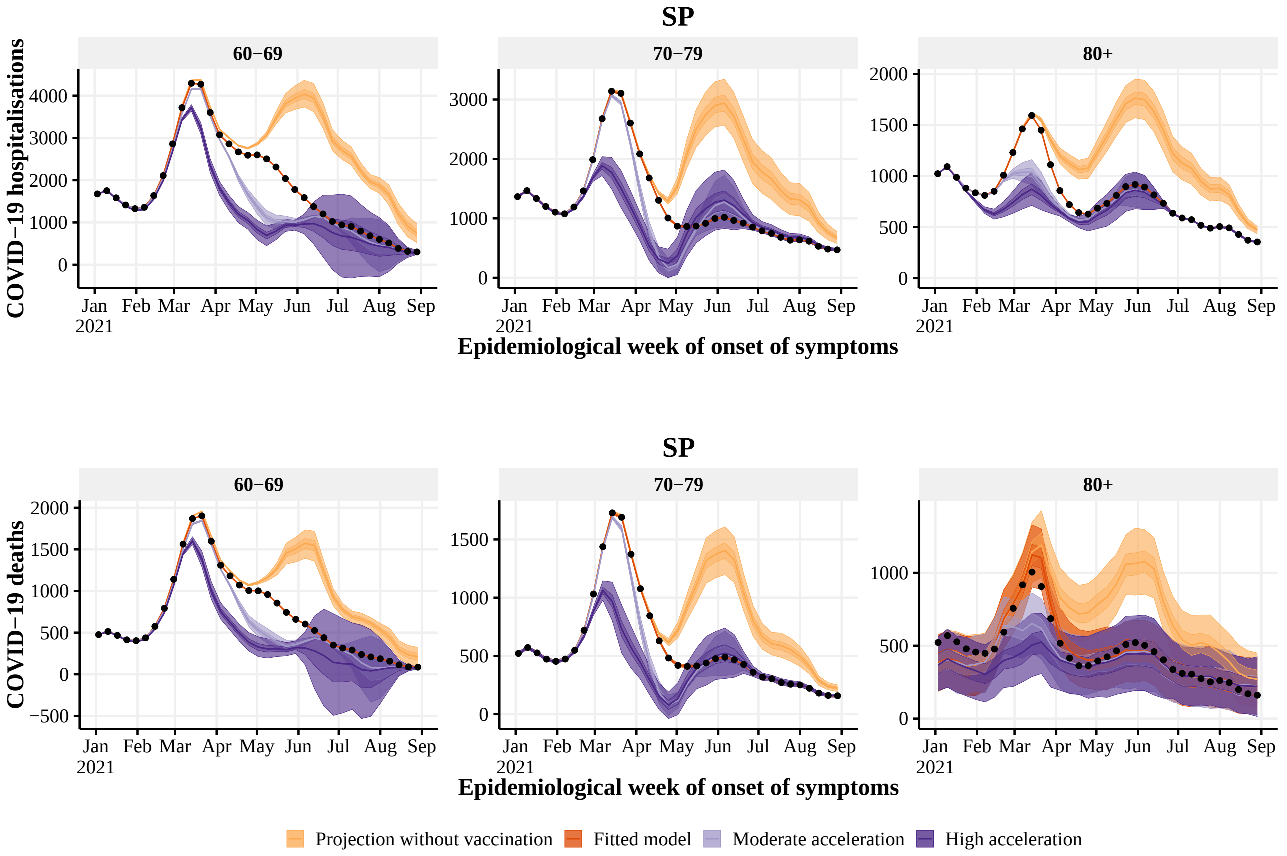


**Figure S46:** Estimated number of hospitalisations (top) and deaths (bottom) by epidemiological week with the realized (dark orange), no vaccination (light orange), 4 (light purple) and 8 (dark purple) weeks earlier vaccination rollout, by age group (panels), in SP state. The observed number of hospitalisations and deaths are given by the black points. With 50% and 95% Credible Intervals.

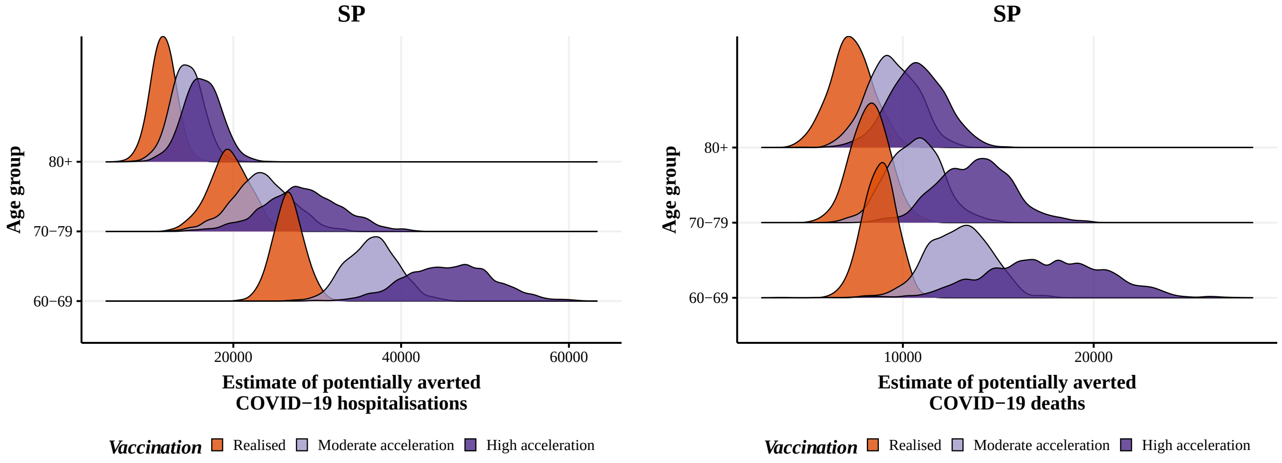


**Figure S47:** Posterior distribution of hospitalisations (left) and deaths (right) potentially averted by vaccination between 2021-01-01 and 2021-08-29 by age group, with the realized (orange), 4 (blue) and 8 weeks earlier (green) vaccination rollout in SP state.

#### 5.6 South Region

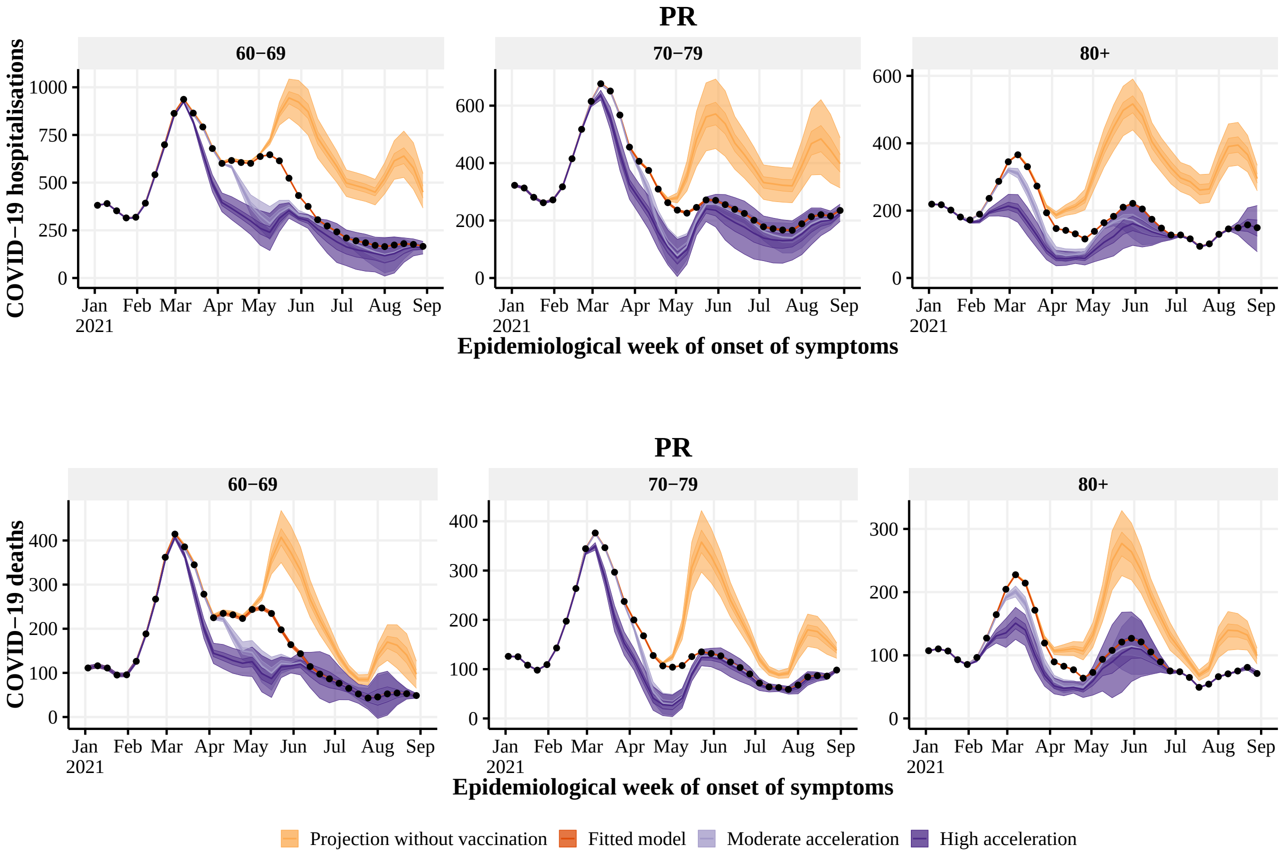


**Figure S48:** Estimated number of hospitalisations (top) and deaths (bottom) by epidemiological week with the realized (dark orange), no vaccination (light orange), 4 (light purple) and 8 (dark purple) weeks earlier vaccination rollout, by age group (panels), in PR state. The observed number of hospitalisations and deaths are given by the black points. With 50% and 95% Credible Intervals.

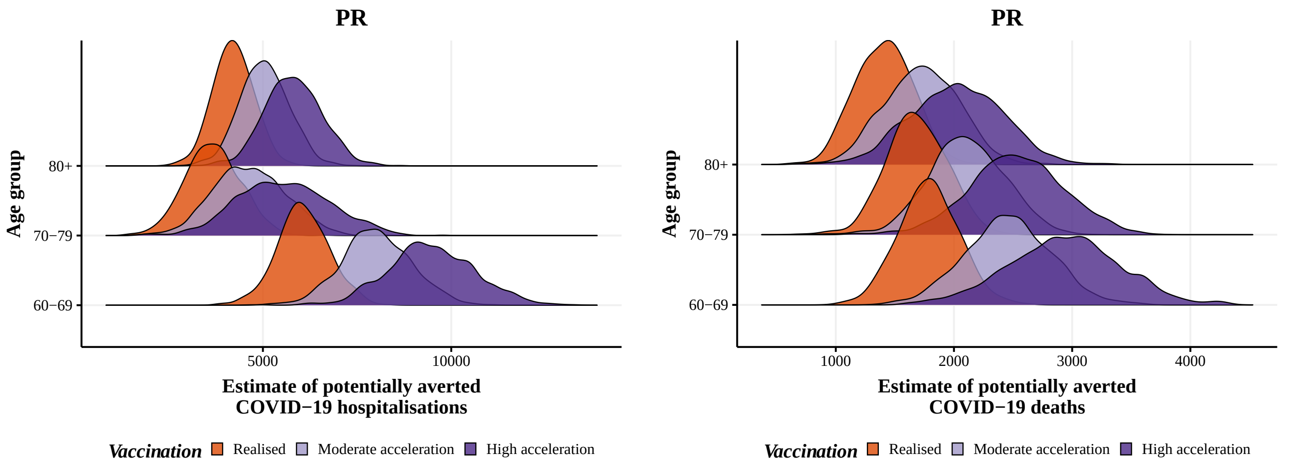


**Figure S49:** Posterior distribution of hospitalisations (left) and deaths (right) potentially averted by vaccination between 2021-01-01 and 2021-08-29 by age group, with the realized (orange), 4 (blue) and 8 weeks earlier (green) vaccination rollout in PR state.

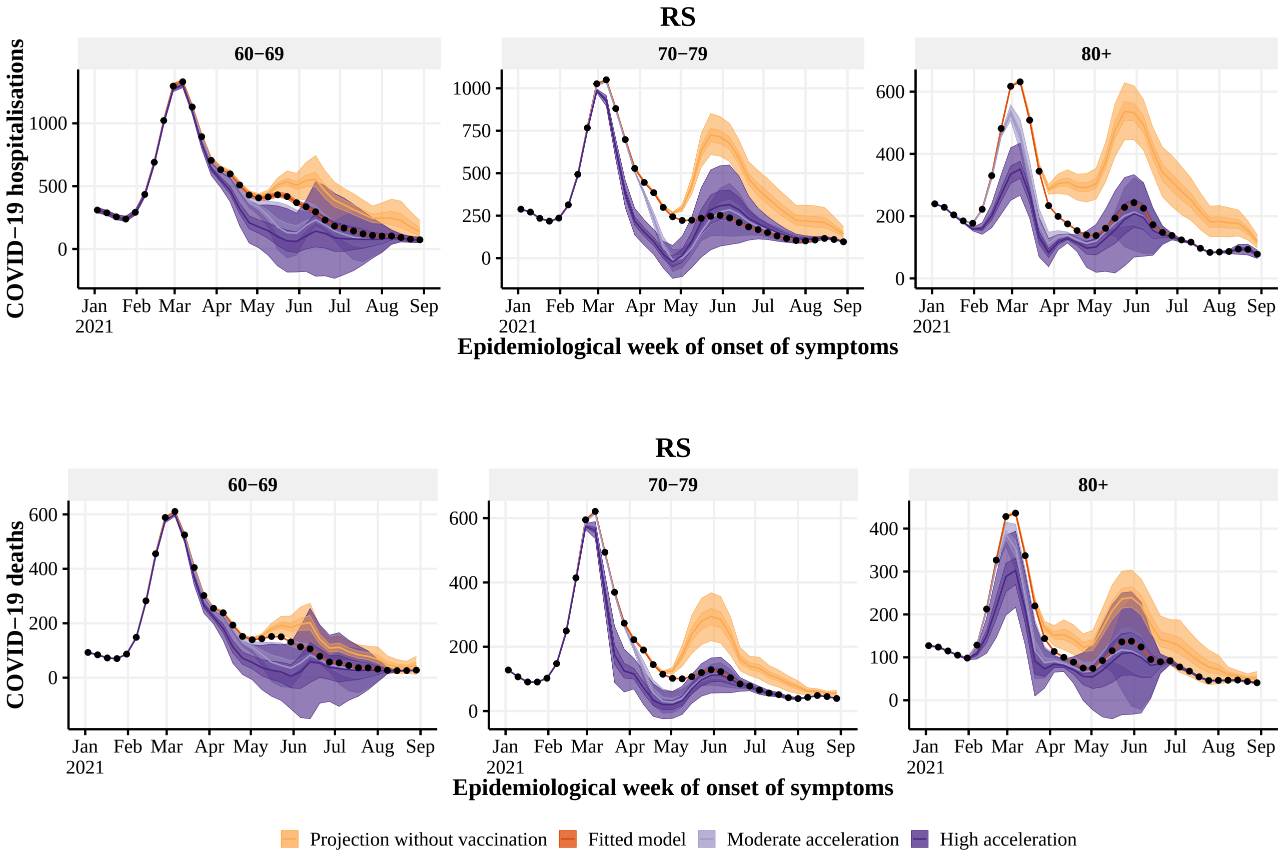


**Figure S50:** Estimated number of hospitalisations (top) and deaths (bottom) by epidemiological week with the realized (dark orange), no vaccination (light orange), 4 (light purple) and 8 (dark purple) weeks earlier vaccination rollout, by age group (panels), in RS state. The observed number of hospitalisations and deaths are given by the black points. With 50% and 95% Credible Intervals.

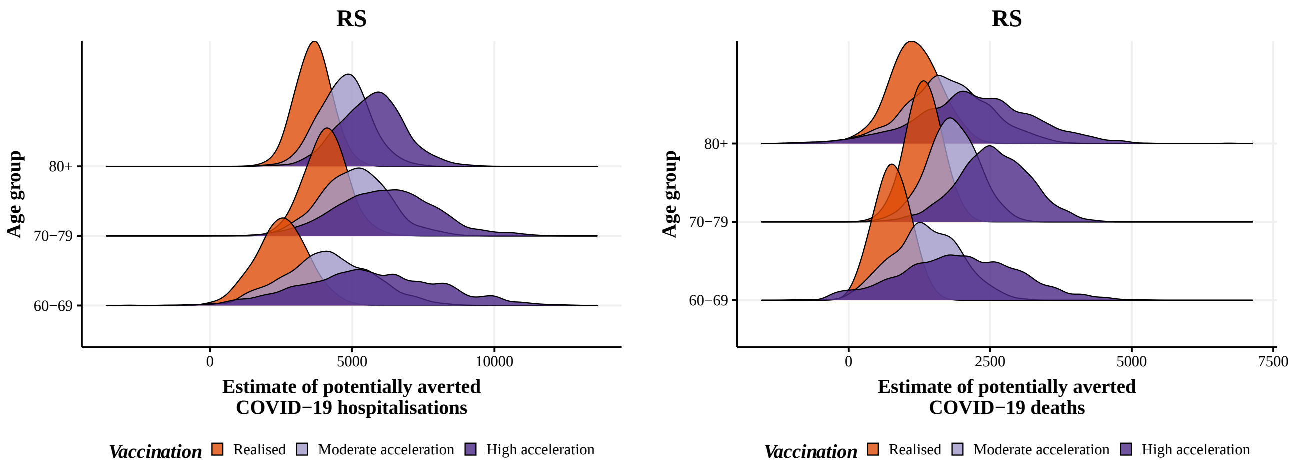


**Figure S51:** Posterior distribution of hospitalisations (left) and deaths (right) potentially averted by vaccination between 2021-01-01 and 2021-08-29 by age group, with the realized (orange), 4 (blue) and 8 weeks earlier (green) vaccination rollout in RS state.

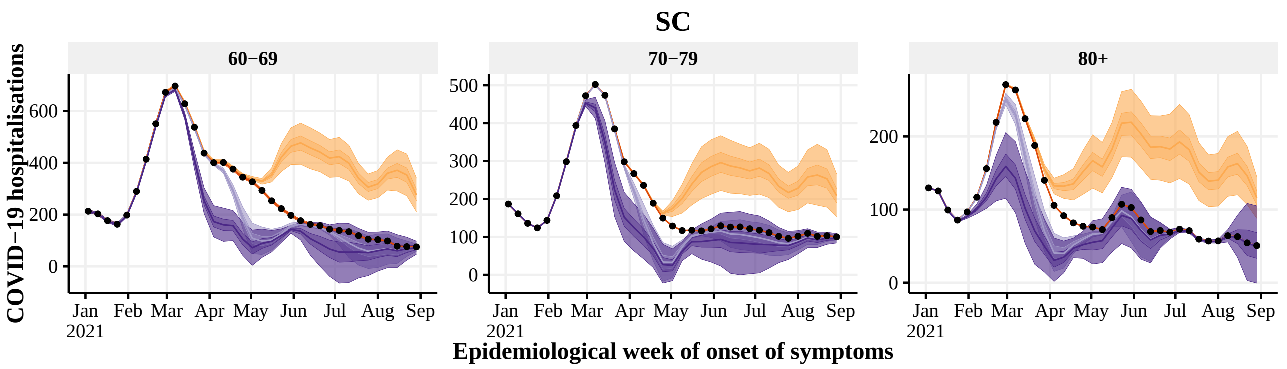


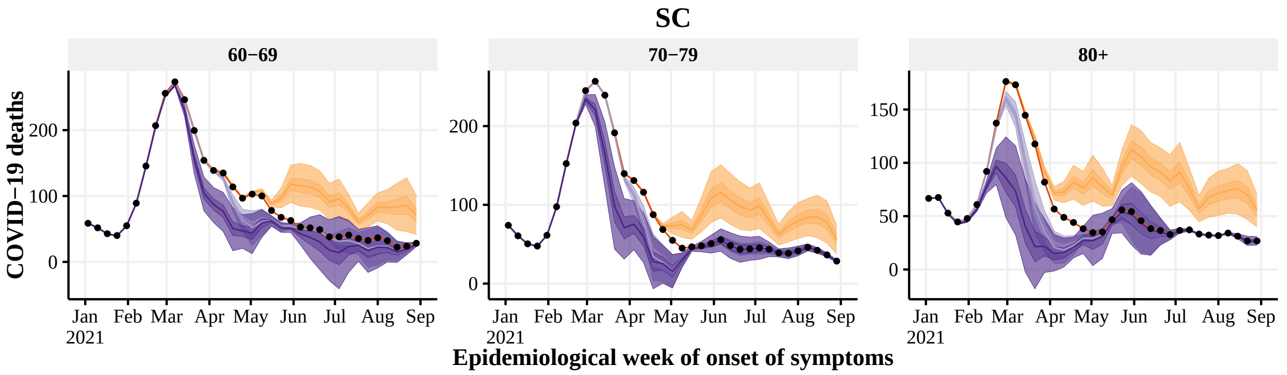


**Figure S52:** Estimated number of hospitalisations (top) and deaths (bottom) by epidemiological week with the realized (dark orange), no vaccination (light orange), 4 (light purple) and 8 (dark purple) weeks earlier vaccination rollout, by age group (panels), in SC state. The observed number of hospitalisations and deaths are given by the black points. With 50% and 95% Credible Intervals.

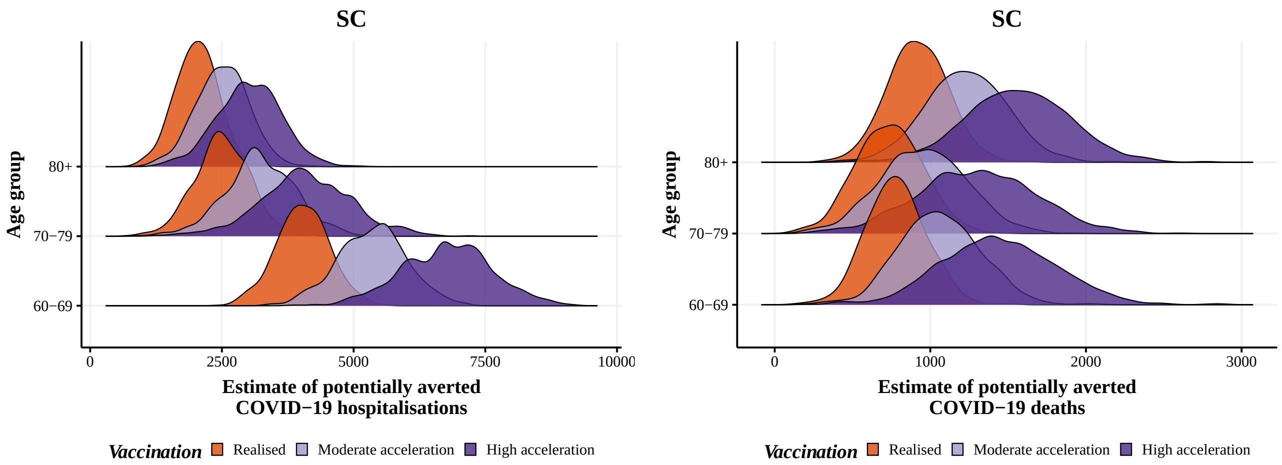


**Figure S53:** Posterior distribution of hospitalisations (left) and deaths (right) potentially averted by vaccination between 2021-01-01 and 2021-08-29 by age group, with the realized (orange), 4 (blue) and 8 weeks earlier (green) vaccination rollout in SC state.

#### 5.7 Center-west Region

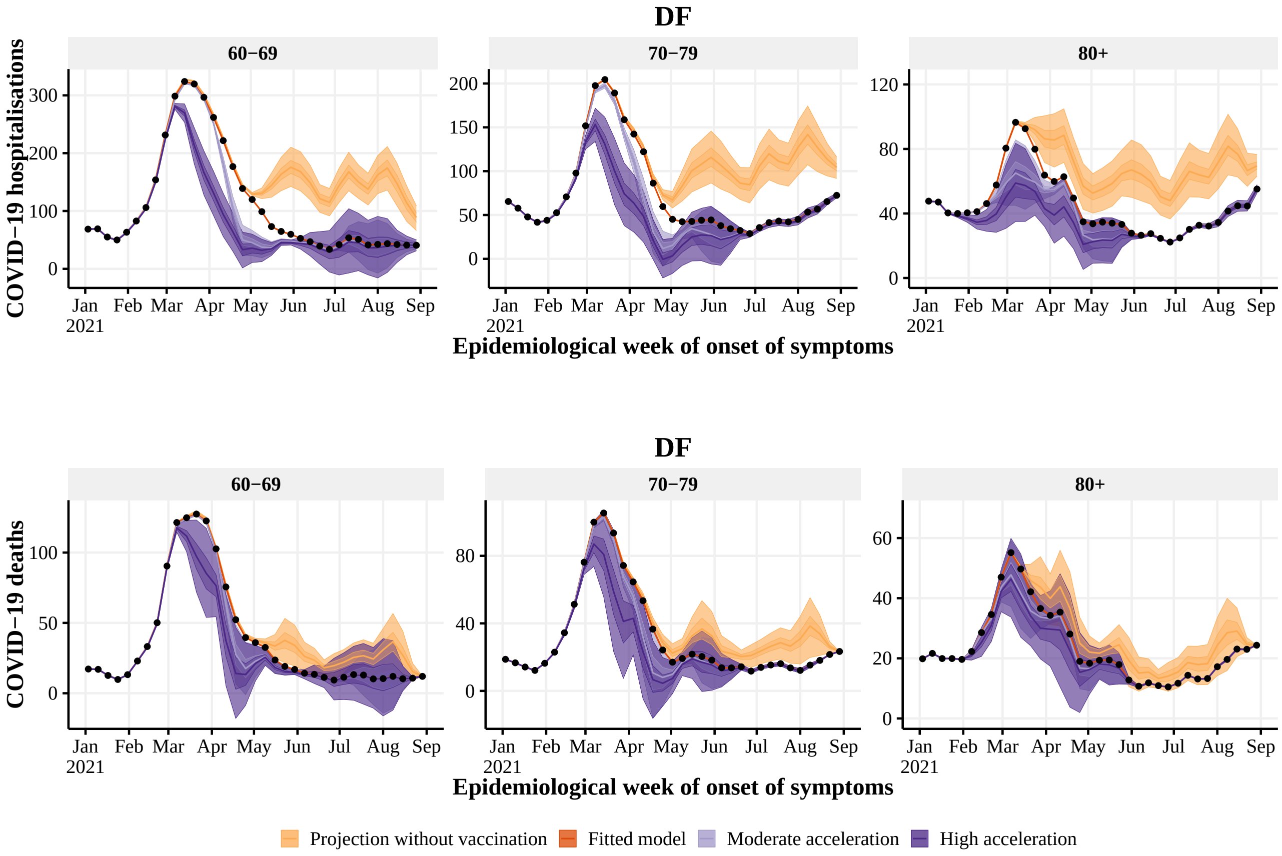


**Figure S54:** Estimated number of hospitalisations (top) and deaths (bottom) by epidemiological week with the realized (dark orange), no vaccination (light orange), 4 (light purple) and 8 (dark purple) weeks earlier vaccination rollout, by age group (panels), in DF state. The observed number of hospitalisations and deaths are given by the black points. With 50% and 95% Credible Intervals.

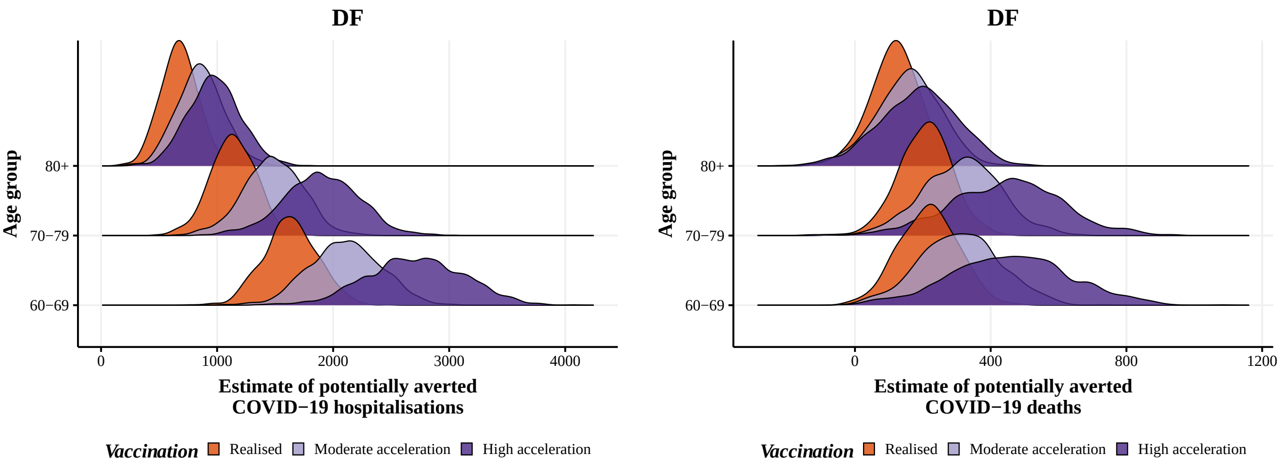


**Figure S55:** Posterior distribution of hospitalisations (left) and deaths (right) potentially averted by vaccination between 2021-01-01 and 2021-08-29 by age group, with the realized (orange), 4 (blue) and 8 weeks earlier (green) vaccination rollout in DF state.

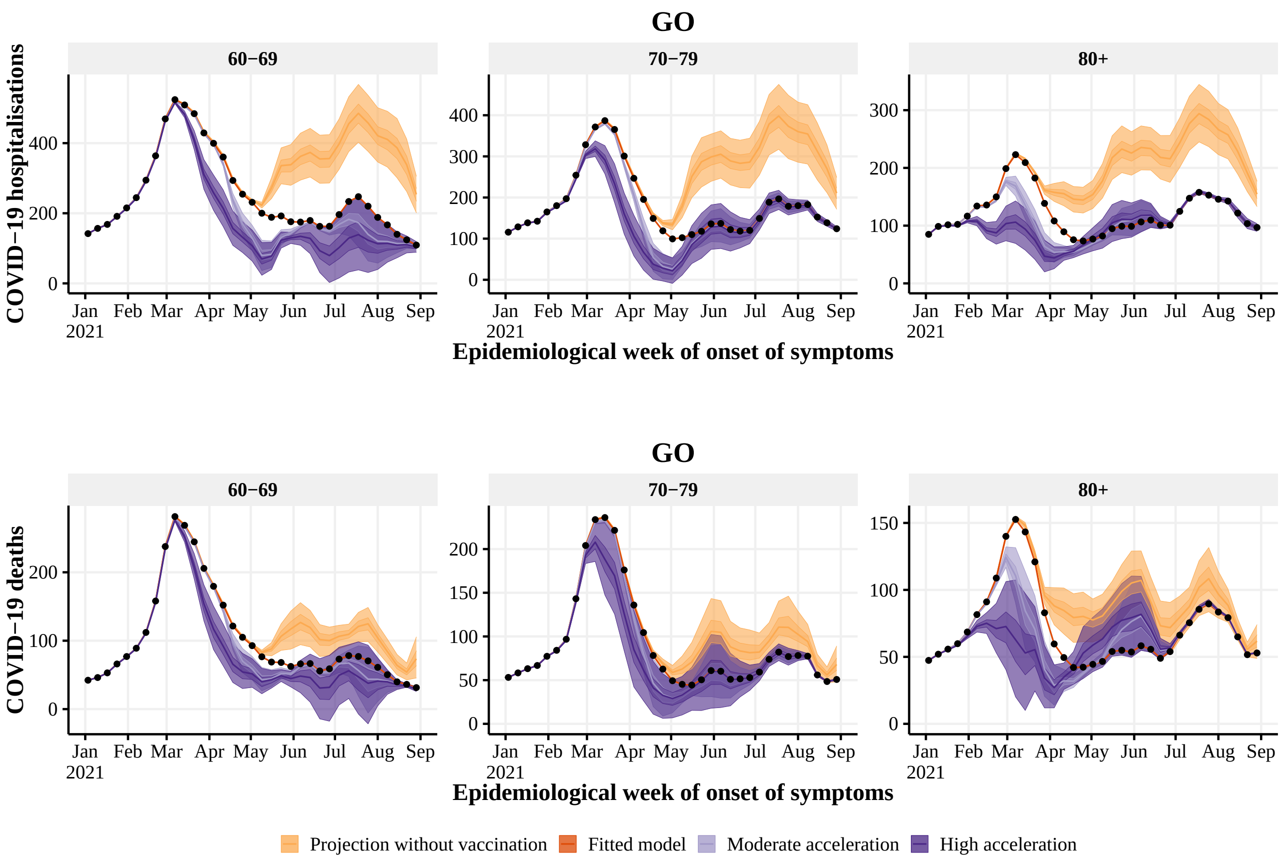


**Figure S56:** Estimated number of hospitalisations (top) and deaths (bottom) by epidemiological week with the realized (dark orange), no vaccination (light orange), 4 (light purple) and 8 (dark purple) weeks earlier vaccination rollout, by age group (panels), in GO state. The observed number of hospitalisations and deaths are given by the black points. With 50% and 95% Credible Intervals.

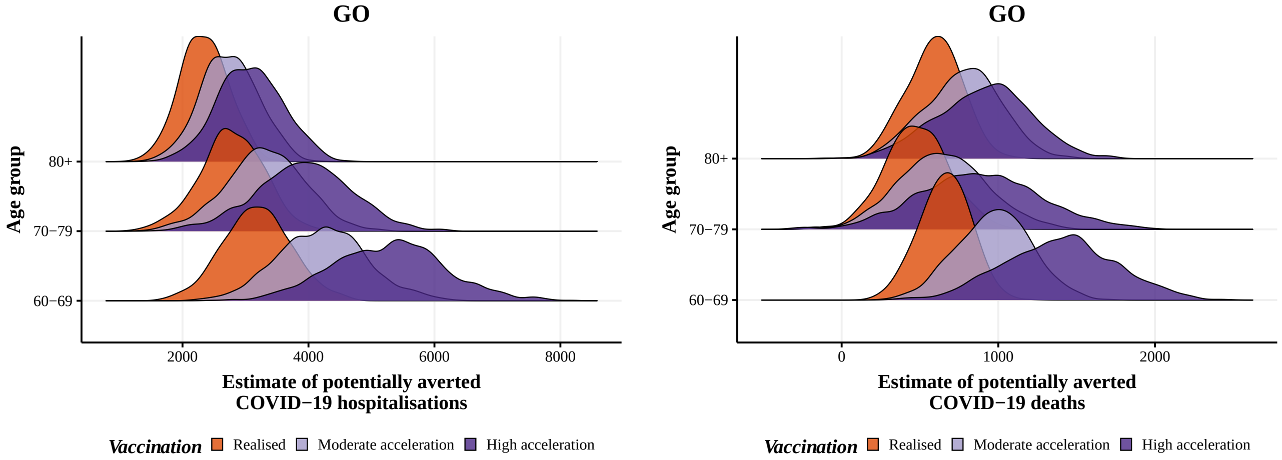


**Figure S57:** Posterior distribution of hospitalisations (left) and deaths (right) potentially averted by vaccination between 2021-01-01 and 2021-08-29 by age group, with the realized (orange), 4 (blue) and 8 weeks earlier (green) vaccination rollout in GO state.

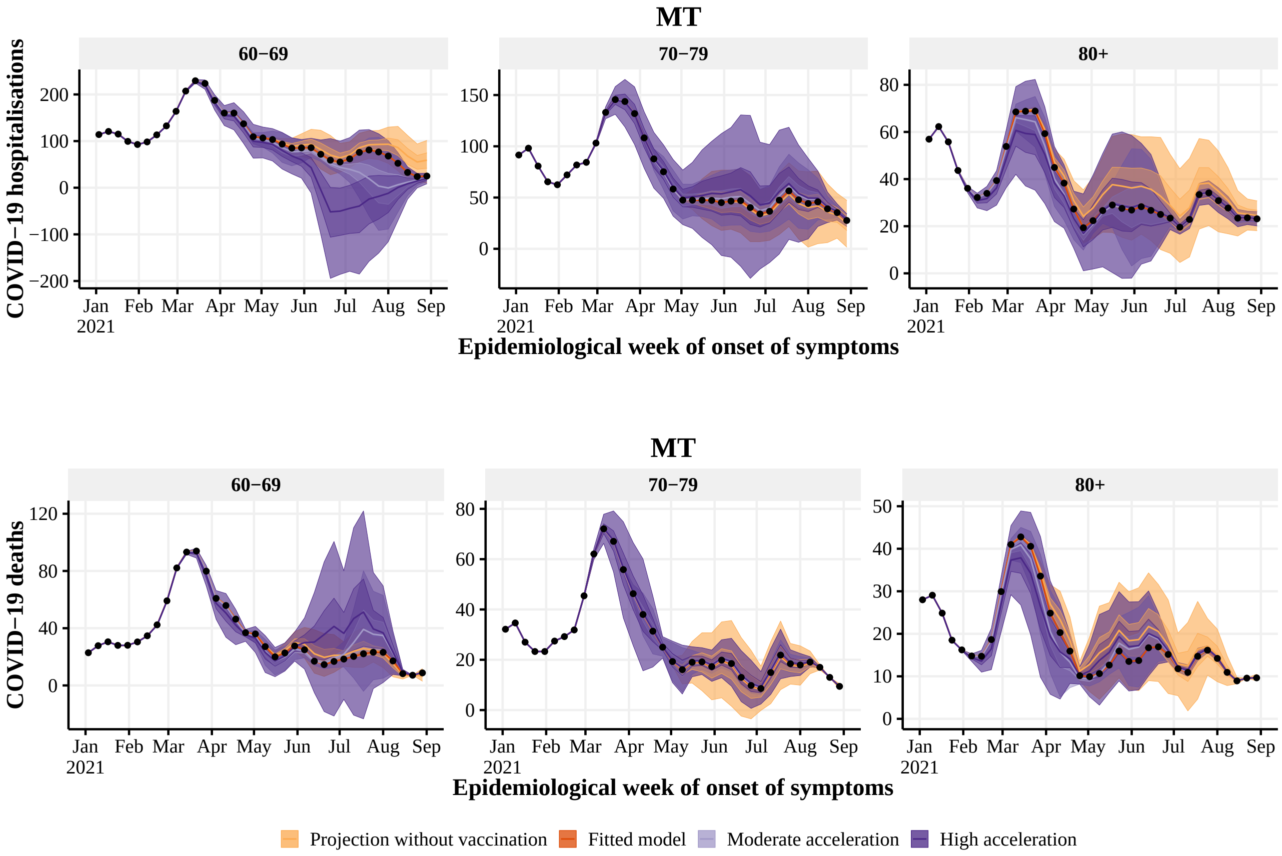


**Figure S58:** Estimated number of hospitalisations (top) and deaths (bottom) by epidemiological week with the realized (dark orange), no vaccination (light orange), 4 (light purple) and 8 (dark purple) weeks earlier vaccination rollout, by age group (panels), in MT state. The observed number of hospitalisations and deaths are given by the black points. With 50% and 95% Credible Intervals.

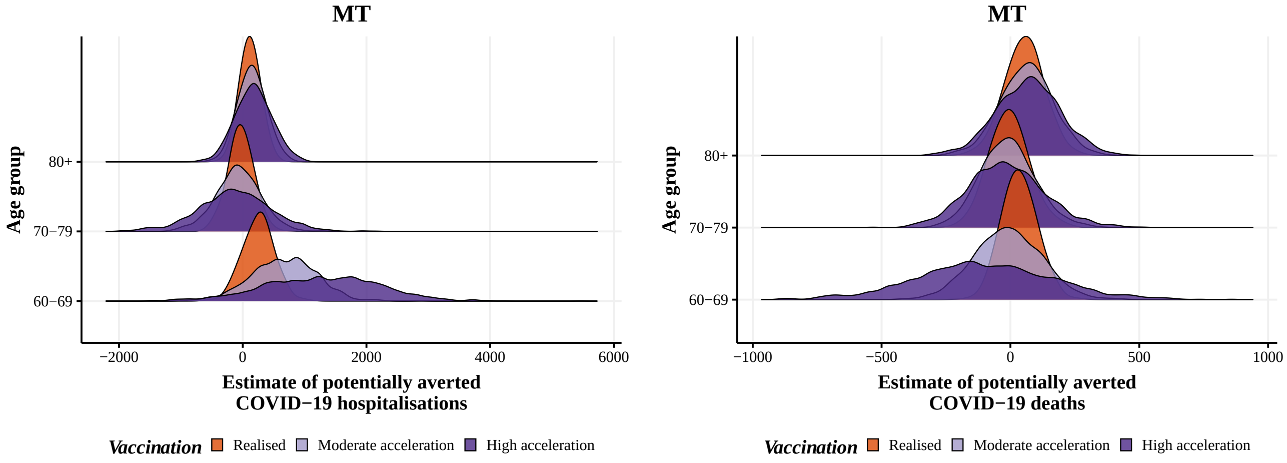


**Figure S59:** Posterior distribution of hospitalisations (left) and deaths (right) potentially averted by vaccination between 2021-01-01 and 2021-08-29 by age group, with the realized (orange), 4 (blue) and 8 weeks earlier (green) vaccination rollout in MT state.

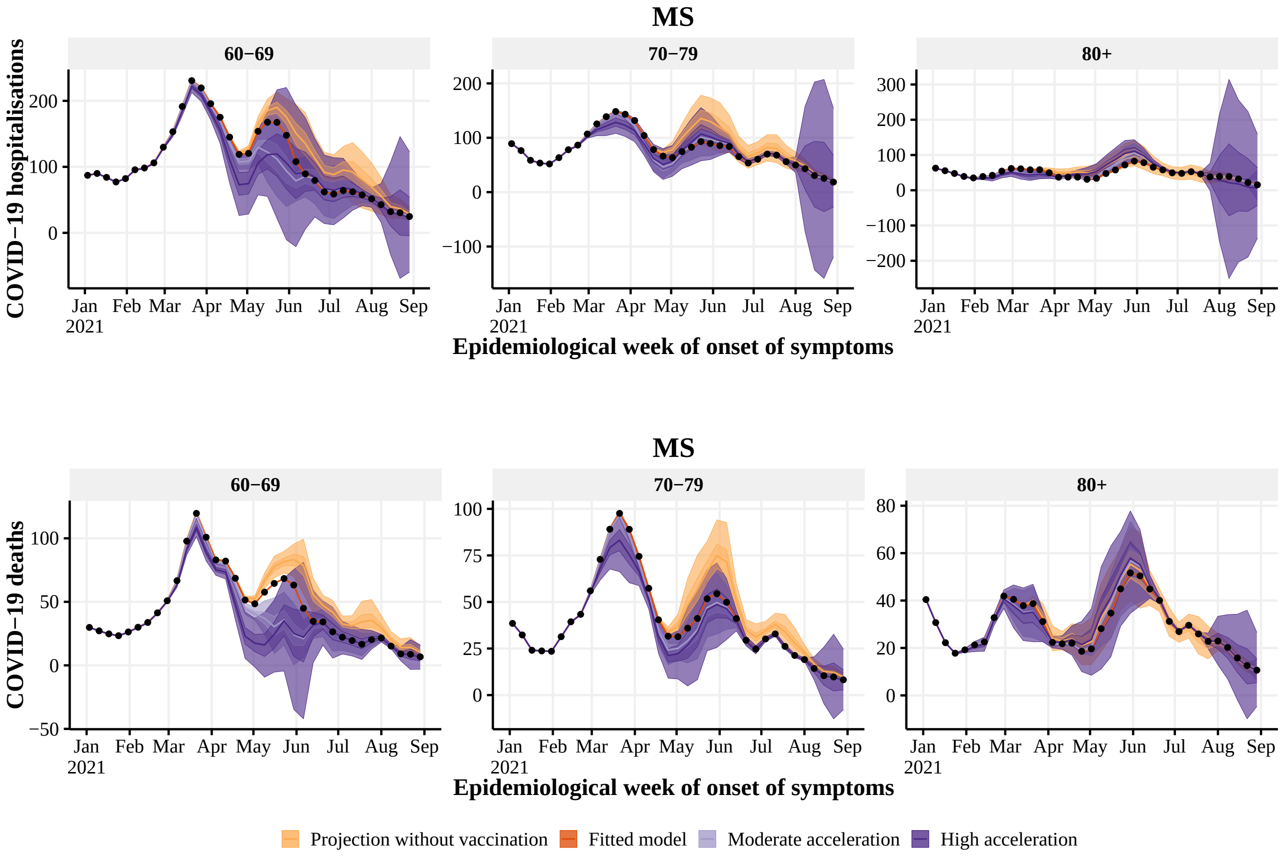


**Figure S60:** Estimated number of hospitalisations (top) and deaths (bottom) by epidemiological week with the realized (dark orange), no vaccination (light orange), 4 (light purple) and 8 (dark purple) weeks earlier vaccination rollout, by age group (panels), in MS state. The observed number of hospitalisations and deaths are given by the black points. With 50% and 95% Credible Intervals.

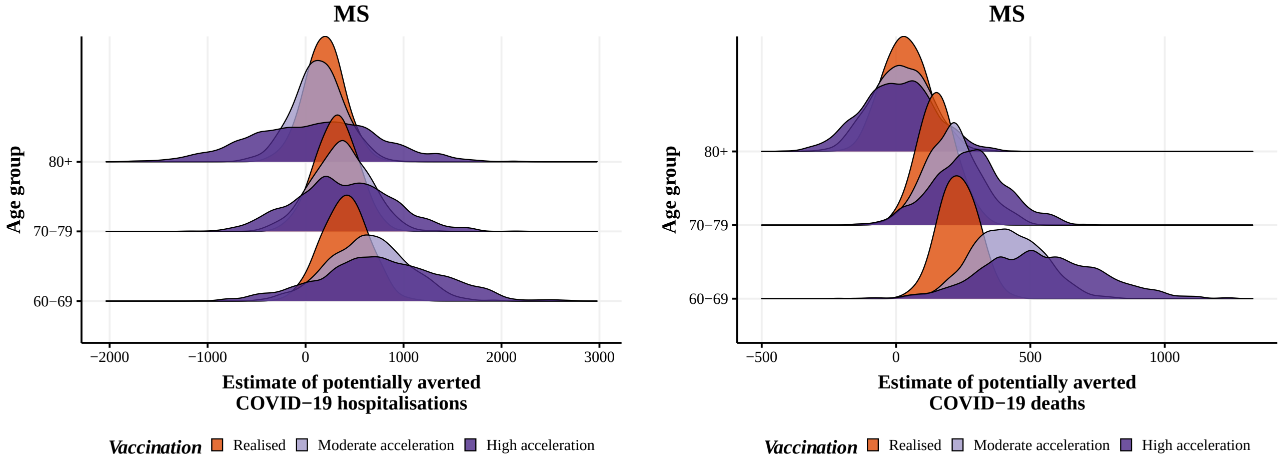


**Figure S61:** Posterior distribution of hospitalisations (left) and deaths (right) potentially averted by vaccination between 2021-01-01 and 2021-08-29 by age group, with the realized (orange), 4 (blue) and 8 weeks earlier (green) vaccination rollout in MS state.
